# Causal effect of C-reactive protein and vitamin D on human cerebral anatomy observed among genetically correlated biomarkers in blood

**DOI:** 10.1101/2021.09.11.21263146

**Authors:** Dylan J. Kiltschewskij, William R. Reay, Murray J. Cairns

## Abstract

Psychiatric disorders such as schizophrenia are commonly associated with structural brain alterations affecting the cortex, which frequently vary with clinically relevant factors including antipsychotic treatment, duration of illness and age of onset. While the underlying variables mediating these structural changes are poorly understood, recent genetic evidence suggests circulating metabolites and other biochemical traits play a causal role in a number of psychiatric disorders which could be mediated by changes in the cerebral cortex. In the current study, we leveraged publicly available genome-wide association study (GWAS) data to explore shared genetic architecture and evidence for causal relationships between a panel of 50 biochemical traits and measures of cortical thickness and surface area at both the global and regional levels. Linkage disequilibrium score regression identified a total of 20 significant and 156 suggestive genetically correlated biochemical-cortical trait pairings, of which six exhibited strong evidence for causality in a latent causal variable model. Interestingly, a negative causal relationship was identified between a unit increase in serum C-reactive protein levels and thickness of the lingual and lateral occipital regions that was also supported by Mendelian randomisation, while circulating vitamin D (25-hydroxyvitamin D) levels exhibited a positive causal effect on temporal pole thickness. Taken together, our findings suggest a subset of biochemical traits exhibit shared genetic architecture and potentially causal relationships with cortical thickness in functionally distinct regions, which may contribute to alteration of cortical structure in psychiatric disorders.

## INTRODUCTION

The pathogenesis of psychiatric disorders is underpinned by a complex interplay of genetic and environmental risk factors. Large-scale genetic studies have identified a strong genetic component of these disorders, characterised by a vast polygenic burden across the genome arising from variants ranging in frequency from common to ultra-rare, as well as large effect size structural variants [1, 2]. Cross-disorder analyses have additionally revealed substantial proportions of shared genetic architecture between psychiatric disorders [3-5], potentially contributing to similarities in clinical presentation and the frequency of observed psychiatric comorbidities. In conjunction with shared genetic risk, many psychiatric disorders are also associated with alteration of brain structure which may account for a range of psychiatric symptoms and offer utility in dissecting underlying pathophysiological mechanisms. Widespread dysregulation of cortical structures in particular has emerged as a consistent feature, with recent neuroanatomical meta-analyses observing grey matter reductions in overlapping cortical regions for schizophrenia, major depressive disorder, bipolar disorder and obsessive-compulsive disorder, amongst others, while structural alterations unique to each disorder have also been identified [6-8]. Although further investigation of these neuroanatomical changes may assist in the discovery of clinically actionable pathways and biomarkers, the fundamental genetic and environmental factors associated with alterations to brain structure remain poorly understood.

Dysregulation of circulating biochemical factors is one broad mechanism through which variations in cortical structure may arise in psychiatric and neurodegenerative disorders. During CNS development, alteration of these circulating factors could plausibly interfere with patterns of neuronal differentiation and migration important for cortical development, while dysregulation in the adult brain may disrupt neuronal cytoarchitecture and integrity. Systemic effects have been widely studied in psychiatry as many of these biochemical variables can be modulated through existing drugs or lifestyle intervention, potentially informing novel treatment interventions. For example, observational evidence suggests elevated C-reactive protein (CRP) in the serum of individuals with schizophrenia is associated with cortical thinning in the frontal, insula and temporal regions [9]. An array of studies has identified many other blood-based biochemical traits with potential diagnostic or prognostic utility [10, 11], however a major drawback of these observational studies is their inability to discriminate between correlations and causal relationships, thus any biological effects resulting in alteration of brain structure are difficult to detect. To this end, genome-wide association studies (GWAS) are proving increasingly valuable for examining and distinguishing genetic correlations and genetically informed causal relationships amongst traits of interest. In particular, germline genetic variants are largely fixed at birth which helps to mitigate effects due to reverse causation in observational studies [12]. Indeed, recent studies utilising GWAS-guided methods of causal inference such as Mendelian randomization have uncovered putative causal relationships between blood-based biochemical traits and psychiatric disorders, using genome-wide significant SNPs as genetic proxies for biochemical exposures [13, 14]. However, no studies to date have examined the fundamental relationship between biochemical traits and structure of the human cerebral cortex. To address this, we employed GWAS summary statistics for a diverse panel of 50 blood-based biochemical traits (UK biobank; http://www.nealelab.is/uk-biobank) to examine shared genetic architecture and putative causal relationships with cortical thickness and surface area measurements (ENIGMA [15]). Our analyses revealed a diverse range of genetic correlations and identified clusters of cortical regions and biochemical traits with similar correlation profiles. In addition, we uncovered evidence for a causal relationship for serum CRP and vitamin D levels on regional cortical thickness, suggesting subsets of circulating biochemical traits may exert biologically significant effects on cortical structure with direct relevance to psychiatric disorders.

## METHODS

### Genome-wide association study data

GWAS summary statistics for cortical surface area (SA) and thickness (TH) as measured via magnetic resonance imaging (MRI) were obtained from the ENIGMA consortium study [15]. These data were generated via meta-analysis of 33,992 individuals of European ancestry, sourced from 49 independent cohorts in the ENIGMA consortium (23,909 individuals) as well as the UK Biobank (UKBB; 10,083 individuals) [16]. We utilised the global measures of mean cortical TH and total SA, as well as regional measures for 34 distinct cortical areas as defined by the Desikan-Killiany atlas. All summary statistics were covaried for age, age^2^, sex, sex-by-age and age^2^ interactions, ancestry (first four MDS components), diagnostic status (for case vs control studies) and dummy variables correcting for the use of multiple scanners.

We additionally obtained summary statistics for a panel of 50 blood-based biochemical traits produced from a cohort of > 300,000 individuals in the UK biobank (http://www.nealelab.is/uk-biobank). These traits included blood cell counts, metabolites, enzymes, lipids and other biomarkers, all of which have high or medium confidence SNP heritability estimates significantly different from zero. See Table S1 for further details.

### Genetic correlation

Genetic correlations amongst cortical and biochemical traits were examined via linkage disequilibrium score regression (LDSR), as described in detail previously [17]. Briefly, LDSR estimates genetic covariance between two traits by regressing SNP-level χ^2^ values – the product of SNP marginal effect sizes from both traits (*Z*_1_*Z*_2_) – with respect to LD scores, which estimate the total LD associated with the SNP of interest. Genetic correlation (*r*_*g*_) is then determined by normalising the genetic covariance by their respective trait heritabilities, with standard errors estimated by jackknifing over 200 blocks of adjacent SNPs. All LDSR analyses were conducted using the *ldsc* python package [18]. We initially analysed biochemical traits with respect to total cortical SA and mean TH, after which regional genetic correlations were explored. Notably, LDSR is robust to sample overlap, thus enabling accurate *r*_*g*_ estimates in the current study despite the presence of UKBB samples in both the biochemical and cortical GWAS. Prior to LDSR analysis, all summary statistics were converted to a standardised “munged” format, wherein approximately one million HapMap 3 SNPs outside the major histocompatibility complex (MHC) with minor allele frequency > 0.05 were retained for further analysis. We utilised LD scores (“eur_w_ld_chr”) previously computed from the 1000 Genomes Project European reference panel available at https://alkesgroup.broadinstitute.org/LDSCORE/. At the global and regional levels, correlations with Bonferroni-corrected *P* < 0.05 (*P*_*Global*_ = 5×10^−4^, 100 comparisons; *PRegional* = 1.471×10^−5^, 3,400 comparisons) were considered statistically significant, while trait pairings with Benjamini-Hochberg false discovery rate (FDR) < 0.05 were considered suggestively significant. To identify biochemical and cortical traits with similar genetic correlation profiles, LDSR *Z*-scores were subjected to unsupervised clustering by finite Gaussian mixture modelling (GMM) using the mclust R package (v5.4.7) [19], with the number of components (clusters) selected using the largest Bayesian Information Criterion (BIC) value.

### Latent causal variable models

To identify genetically correlated trait pairings with evidence for a causal relationship, we employed a latent causal variable (LCV) model to estimate genetic causality as described previously [20]. Briefly, the LCV model assumes a latent variable, *L*, mediates the genetic correlation between two traits such that if trait 1 is strongly correlated with *L*, it is assumed to be partially genetically causal for trait 2. To test for partial genetic causality, the mixed fourth moments (cokurtosis) of SNP marginal effect size distributions for each trait are compared to determine whether SNPs affecting trait 1 have proportional effects on trait 2, but not *vice versa*. For interpretation, the LCV model reports a posterior mean genetic causality proportion 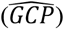 of trait 1 on trait 2, wherein 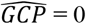 suggest no causal relationship, 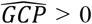 implies trait 1 is partially genetically causal for trait 2 and 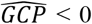 suggests trait 2 is partially genetically causal for trait 1. We consider significant 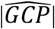 estimates > 0.6 as strong evidence for partial genetic causality as shown previously [20], noting that 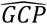 summarises the strength of evidence for a genetically causal relationship, rather than the magnitude of a causal effect. We note that, like LDSR, the LCV model is resistant to statistical inflation arising from sample overlap due to the incorporation of the LDSR intercept. All summary statistics subjected to LCV modelling were “munged” prior to analysis as recommended.

### Mendelian randomisation

Two-sample Mendelian randomization (MR) was employed to further explore and quantify the magnitude of causal relationships amongst trait pairings with evidence for partial genetic causality via LCV modelling. MR leverages genetic instrumental variables (IVs) rigorously associated with an exposure – most commonly independent genome-wide significant SNPs – to estimate the causal effect of the exposure on an outcome. We specifically tested the effect of C-reactive protein (CRP) and vitamin D (measured as 25-hydroxyvitamin D) exposures on regional cortical TH measurements utilizing LD-clumped, non-palindromic IVs sourced from well-powered non-UK Biobank GWAS [21, 22], as most two-sample MR methods are sensitive to sample overlap. For both GWAS, the IV *F*-statistic was > 10, indicating IVs for both exposures are sufficiently powered [23]. All exposure-outcome trait pairings were analysed via five MR models with differing underlying assumptions regarding the validity of using SNPs as IVs. Specifically, the inverse variance weighted model with multiplicative random effects (IVW_m_) was used as our principal model, which is generally considered the most-well powered model but has a zero percent breakdown point, and thus, assumes all IVs are valid [24]. We also incorporated an IVW model with fixed effects (IVW_f_), which is a less conservative iteration of the IVW estimator that accounts for inter-instrument heterogeneity in a more-general manner, and thus is more statistically valid at the risk of additional bias in the presence of heterogeneity. To account for potential violations of the “all-valid” assumption, we additionally utilised a weighted median model, which through the *plurality valid* assumption is said to be an unbiased estimator if at least 50% of the weight in the model is derived from valid IVs [25]. A weighted mode model was additionally employed, which also tests the *plurality valid* assumption and is more-robust to outlying IVs at the expense of power [26]. Finally, the MR Egger approach was also utilised which does not constrain the intercept as zero, with a significantly non-zero intercept considered to be evidence of potential unbalanced pleiotropy [27]. We also assessed whether there was further evidence of horizontal pleiotropy or other outlier effects via the following: Cochran’s *Q* to test heterogeneity in the individual IV exposure-outcome estimates [28], iteratively leaving out each IV and recalculating the IVW estimate (leave-one-out) [28], and the MR PRESSO test of global pleiotropy that is also related to heterogeneity [29]. Finally, the Steiger directionality test was implemented to evaluate evidence that the assumed causal direction (biochemical trait → cortical property) was correct through comparing the variance explained by the IVs in the exposure to their association with the cortical outcome GWAS [30]. All MR analyses were conducted using the *TwoSampleMR* (v.0.5.5) [31] and *MRPRESSO* (v.1.0) [29] *R* packages.

### Transcriptome-wide association studies

Transcriptome-wide association studies (TWAS) were conducted using the *FUSION* approach to identify genes associated with biochemical traits and cortical TH, as well as a predicted direction of expression [32]. TWAS utilizes *cis*-acting genetic variants to predict gene expression optimised through competing multivariate models (genetically regulated expression – GReX). Genes that have significantly heritable expression explained by *cis*-acting variants can be evaluated with this method. Specifically, the most predictive GReX model is utilised to correlate predicted expression with the GWAS trait of interest by integrating the SNP weights from the expression model with their corresponding effect estimated by the GWAS. As a result, the covariance between expression and the trait can be leveraged to interpret the direction of association positively correlated with the trait – for example, a gene with a positive TWAS test statistic (*Z* > 0) in the context of a continuous GWAS trait signifies that increased expression of the gene is correlated with an increase in the trait. We utilised SNP weights from both the blood and cortex from GTEx v7 [33]. In addition, since CRP has a well-documented function in the liver, we also included liver expression weights for this exposure. Tissue-specific, transcriptome-wide genetic correlations between traits were additionally examined using the *RHOGE R* package [34].

## RESULTS

### Extensive genetic correlation amongst biochemical traits and cortical properties

To identify blood-based biomarkers genetically correlated with global changes in cortical structure, we estimated correlations of a panel of 50 biochemical traits with respect to global mean cortical thickness (TH) and total surface area (SA) using LDSR. While no trait pairings survived multiple testing correction (*P*_*Bonferroni*_ < 0.05), mean cortical TH was positively correlated with haematocrit percentage (*r*_*g*_ = 0.125, *SE* = 0.037, *P* = 0.0008), haemoglobin (*r*_*g*_ = 0.12, *SE* = 0.037, *P* = 0.0011) and red blood cell (RBC) count (*r*_*g*_ = 0.117, *SE* = 0.035, *P* = 0.0009; Fig. 1a, Table 1, Table S2) after correction using the FDR (*P*_*BH*_ < 0.05). These specific correlations are broadly supportive of observational association between cerebral haemodynamics and cortical thinning [35], suggesting that global cortical SA and TH display some evidence of overlapping genetic architecture with this panel of biochemical traits.

**Figure 1.**
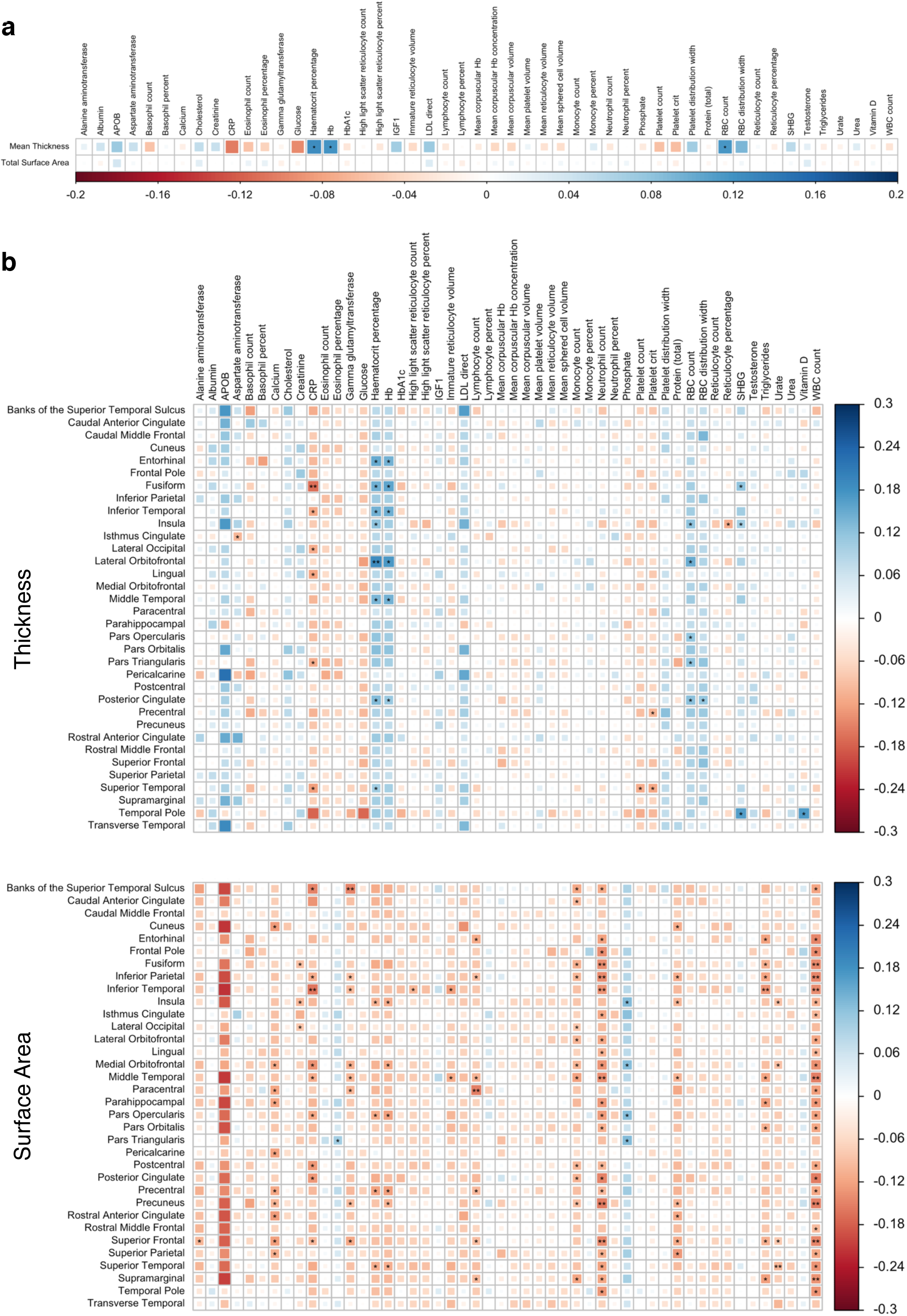
Genetic correlation amongst GWAS for biochemical markers and structural cortical traits. **(a)** Heatmap of LDSR genetic correlation coefficients (*r*_*g*_) between mean cortical TH and total cortical SA (averaged across both hemispheres) with respect to 50 biochemical traits. * = *P*_*Benjamini-Hochberg*_ < 0.05, ** = *P*_*Bonferroni*_ < 0.05. **(b)** As in **(a)**, except depicting genetic correlations between biochemical traits with respect to TH (top) and SA (bottom) of 34 cortical regions as defined by the Desikan-Killiany atlas.

**Table 1.** Top 10 genetic correlations for mean cortical TH and total SA.

| Cortical trait | Biochemical trait | $r_g^1$ | SE | P | $P_{BH}$ | $P_{Bonf}$ |
| --- | --- | --- | --- | --- | --- | --- |
| Mean thickness | Haematocrit % | 0.125 | 0.037 | $8 \times 10^{-4}$ | 0.037 | 0.08 |
| | RBC count | 0.117 | 0.035 | $9 \times 10^{-4}$ | 0.037 | 0.09 |
|  | Haemoglobin | 0.12 | 0.037 | 0.001 | 0.037 | 0.11 |
|  | C-reactive protein | -0.103 | 0.034 | 0.003 | 0.068 | 0.27 |
|  | Glucose | -0.096 | 0.036 | 0.009 | 0.172 | 0.86 |
|  | RBC distribution width | 0.091 | 0.038 | 0.018 | 0.29 | 1 |
|  | Platelet crit | -0.062 | 0.028 | 0.029 | 0.33 | 1 |
|  | Platelet count | -0.059 | 0.028 | 0.033 | 0.33 | 1 |
|  | Insulin-like growth factor 1 | 0.069 | 0.033 | 0.036 | 0.33 | 1 |
|  | Sex hormone binding globulin | 0.054 | 0.028 | 0.057 | 0.461 | 1 |
| Total surface area | Testosterone | 0.028 | 0.012 | 0.02 | 0.29 | 1 |
|  | Eosinophil % | 0.019 | 0.009 | 0.036 | 0.33 | 1 |
|  | Platelet crit | -0.018 | 0.009 | 0.061 | 0.461 | 1 |
|  | Cholesterol | 0.026 | 0.014 | 0.065 | 0.461 | 1 |
|  | Eosinophil count | 0.016 | 0.009 | 0.079 | 0.492 | 1 |
|  | Gamma glutamyltransferase | -0.016 | 0.009 | 0.095 | 0.522 | 1 |
|  | Lymphocyte % | -0.017 | 0.010 | 0.099 | 0.522 | 1 |
|  | LDL direct | 0.03 | 0.019 | 0.101 | 0.549 | 1 |
|  | Glycosylated haemoglobin | -0.014 | 0.009 | 0.131 | 0.626 | 1 |
|  | Mean corpuscular volume | 0.012 | 0.009 | 0.191 | 0.677 | 1 |

<sup>1</sup>Genetic correlation coefficient from LDSR.

We then explored site-specific genetic correlations throughout the cortex by analysing measures of SA and TH for 34 distinct regions. For regional SA, 18 traits pairings survived multiple testing correction while an additional 120 suggestive pairings were identified, involving a total of 18 biochemical traits and 32 cortical regions (Fig. 1b, Table 2, Table S3). Notably, 95.8% of these correlations were negative, with particularly strong representation from white blood cell (WBC) count (26), neutrophil count (23) and monocyte count (12), while the superior frontal (10), middle temporal (9), medial orbitofrontal (8) and inferior parietal (8) regions were most consistently affected. The extent of genetic correlation for regional TH was comparatively modest, with two significant and 33 suggestive trait pairings identified involving 11 biochemical traits and 15 cortical phenotypes (Fig. 1b, Table 2, Table S3). Although 72.7% of correlations were positive, many of these involved haematocrit percentage (8), haemoglobin (6) and red blood cell count (5), which were also suggestively correlated with global mean cortical TH (Fig. 1a). C-reactive protein, however, exhibited no response at the global level but was negatively correlated with the TH of six regions at a *P*_*BH*_ < 0.05, including the fusiform (*r*_*g*_ = –0.167, *SE* = 0.0367, *P* = 4.45×10^−6^, survived *P*_*Bonferroni*_ < 0.05), superior temporal (*r*_*g*_ = –0.126, *SE* = 0.041, *P* = 0.002), lingual (*r*_*g*_ = –0.118, *SE* = 0.038, *P* = 0.0018), inferior temporal (*r*_*g*_ =–0.115, *SE* = 0.034, *P* = 0.0008), lateral occipital (*r*_*g*_ = –0.111, *SE* = 0.034, *P* = 0.0013) and pars triangularis (*r*_*g*_ = –0.109, *SE* = 0.036, *P* = 0.0023). Strikingly, these data support the observational data that reports a negative correlation between CRP levels and cortical thinning in individuals with schizophrenia and healthy controls [9].

**Table 2.** Top 10 genetic correlations for regional cortical TH and SA.

| Metric | Cortical region | Biochemical trait | $r_g$ | $SE$ | $P$ | $P_{BH}$ | $P_{Bonf}$ |
| --- | --- | --- | --- | --- | --- | --- | --- |
| Thickness | Fusiform | C-reactive protein | -0.167 | 0.037 | $4.45 \times 10^{-6}$ | $1.53 \times 10^{-3}$ | 0.015 |
| | Lateral orbitofrontal | Haematocrit % | 0.189 | 0.043 | $1.02 \times 10^{-5}$ | $1.94 \times 10^{-3}$ | 0.035 |
| | Lateral orbitofrontal | Haemoglobin | 0.172 | 0.041 | $3.24 \times 10^{-5}$ | $4.4 \times 10^{-3}$ | 0.11 |
| | Middle temporal | Haemoglobin | 0.144 | 0.037 | $8.82 \times 10^{-5}$ | $9.19 \times 10^{-3}$ | 0.3 |
| | Superior temporal | Platelet crit | -0.114 | 0.03 | $1 \times 10^{-4}$ | $9.19 \times 10^{-3}$ | 0.34 |
| | Lateral orbitofrontal | RBC count | 0.147 | 0.039 | $1 \times 10^{-4}$ | $9.19 \times 10^{-3}$ | 0.34 |
| | Superior temporal | Platelet count | -0.109 | 0.029 | $2 \times 10^{-4}$ | 0.015 | 0.68 |
| | Posterior cingulate | RBC count | 0.126 | 0.034 | $2 \times 10^{-4}$ | 0.015 | 0.68 |
| | Middle temporal | Haematocrit % | 0.137 | 0.038 | $3 \times 10^{-4}$ | 0.017 | 1 |
| | Posterior cingulate | Haematocrit % | 0.132 | 0.036 | $3 \times 10^{-4}$ | 0.017 | 1 |
| Surface area | Precuneus | WBC count | -0.151 | 0.029 | $2.74 \times 10^{-7}$ | $7.5 \times 10^{-4}$ | $9.32 \times 10^{-4}$ |
| | Precuneus | Neutrophil count | -0.155 | 0.031 | $4.42 \times 10^{-7}$ | $7.51 \times 10^{-4}$ | $1.5 \times 10^{-3}$ |
| | Superior frontal | WBC count | -0.146 | 0.029 | $6.99 \times 10^{-7}$ | $7.92 \times 10^{-4}$ | $2.38 \times 10^{-3}$ |
| | Middle temporal | WBC count | -0.151 | 0.031 | $1.07 \times 10^{-6}$ | $9.07 \times 10^{-4}$ | $3.63 \times 10^{-3}$ |
| | Inferior temporal | WBC count | -0.139 | 0.029 | $1.75 \times 10^{-6}$ | $1.19 \times 10^{-3}$ | $5.94 \times 10^{-3}$ |
| | Fusiform | WBC count | -0.144 | 0.031 | $3.29 \times 10^{-6}$ | $1.46 \times 10^{-3}$ | 0.011 |
| | Inferior temporal | C-reactive protein | -0.16 | 0.034 | $3.39 \times 10^{-6}$ | $1.46 \times 10^{-3}$ | 0.02 |
| | Middle temporal | Neutrophil count | -0.14 | 0.030 | $3.43 \times 10^{-6}$ | $1.46 \times 10^{-3}$ | 0.012 |
| | Inferior parietal | WBC count | -0.146 | 0.032 | $4.49 \times 10^{-6}$ | $1.53 \times 10^{-3}$ | 0.015 |
| | BSTS<sup>1</sup> | GGT<sup>2</sup> | -0.149 | 0.033 | $5.69 \times 10^{-6}$ | $1.7 \times 10^{-3}$ | 0.019 |

<sup>1</sup>Banks of the superior temporal sulcus.
<sup>2</sup>Gamma glutamyltransferase.

### Clustering of genetic correlation profiles reveals cortical regions with similar biochemical relationships

To identify cortical regions with similar biochemical correlation profiles, we compared the computed LDSR *Z*-scores (*r*_g_/*SE*) amongst all cortical phenotypes. For both SA and TH, we observed a large number of positively correlated regions which loosely clustered by their spatial proximity after application of hierarchal clustering (Figs. 2a). For instance, analysis of cortical TH revealed a subset of seven adjacent regions in the frontal lobe (lateral and medial orbitofrontal, pars opercularis, pars triangularis, rostral and caudal middle frontal and superior frontal) with highly correlated biochemical *Z*-score profiles (Fig. 2a). We additionally compared correlation profiles amongst biochemical traits, wherein physiologically related biochemical markers (e.g. immune cell traits, reticulocyte traits) tended to cluster in both the SA and TH analyses (Fig. S1). In contrast to the cortical phenotypes, these traits exhibited a wider range of both positive and negative correlations.

**Figure 2.**
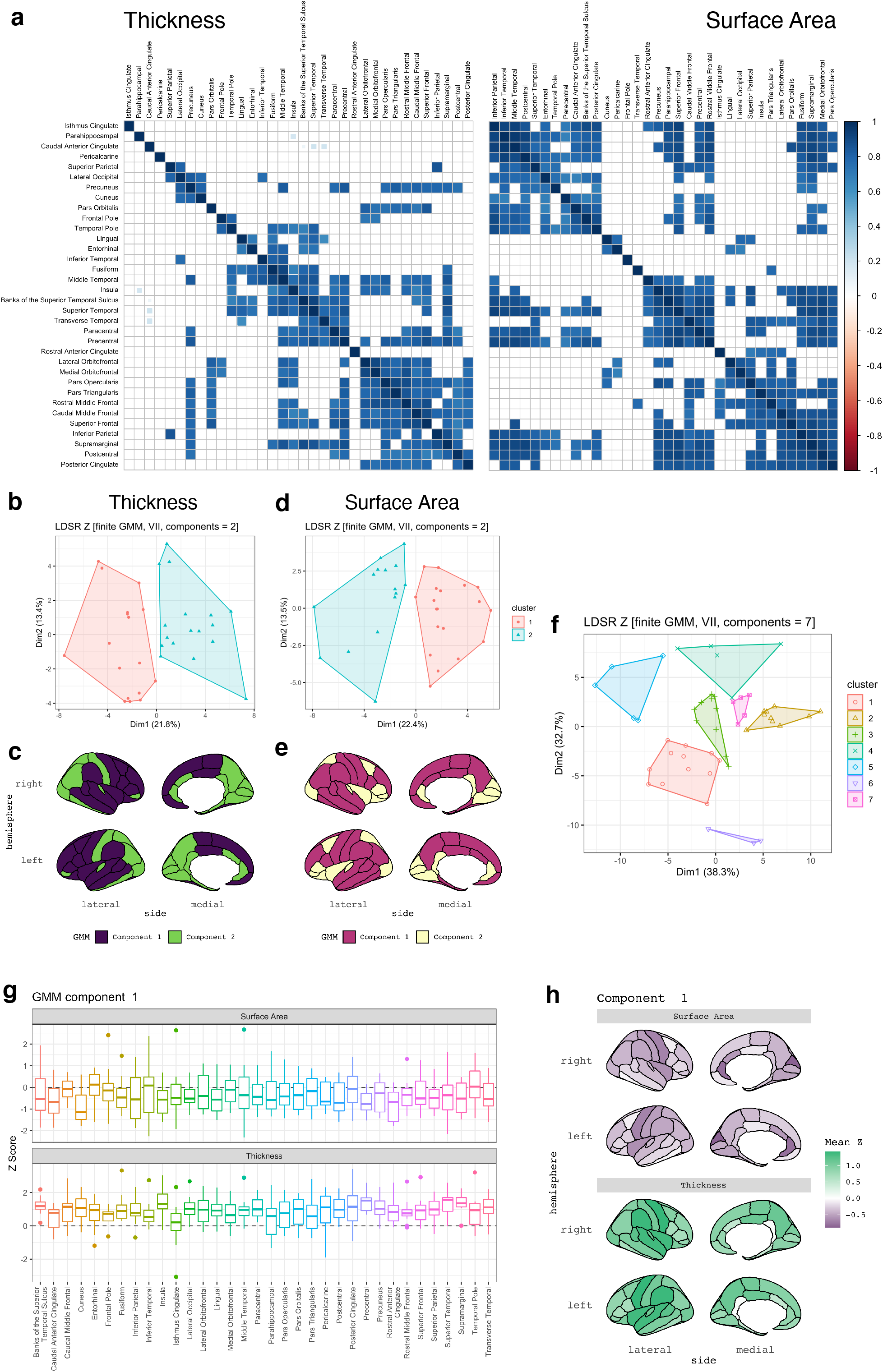
Clustering of cortical and biochemical correlation profiles. **(a)** Heatmaps depicting correlations between cortical regions after comparison of LDSR biochemical *Z-*score profiles. Cortical trait pairings with *P*_*Bonferroni*_ < 0.05 are shown, with hierarchal clustering via Ward’s D2 method. **(b)** Clustering of cortical TH measures via biochemical LDSR *Z*-score profiles using finite Gaussian mixture modelling (GMM). Optimal parameterisation of the covariance matrix was obtained using two clusters (components) of cortical regions with spherical distribution, variable volume and equal shape (VII). Components are plotted via their contribution to the first and second principal components of the LDSR *Z* matrix. **(c)** Spatial distribution of GMM components identified in panel **(b)**. Note that component one (purple) is largely confined to the frontal and temporal lobes, whereas component two predominantly consists of parietal and occipital regions. **(d & e)** As in **(b & c)**, except examining clusters of cortical regions subset by SA. **(f)** GMM clustering of biochemical traits by their TH and SA *Z-*score profiles, which revealed a total of seven biochemical clusters. **(g)** Boxplots of LDSR *Z*-scores for all biochemical traits in component one, including *APOB, albumin, aspartate aminotransferase, cholesterol, IGF1, LDL direct, monocyte percent, platelet distribution width, RBC distribution width, sex hormone binding globulin, testosterone* and *urea*. Note that *Z*-scores for these biochemical traits were generally negative with respect to cortical SA and positive with respect to cortical TH. **(h)** Spatial organisation of mean *Z-*scores for biochemical traits in component 1.

We subsequently employed finite Gaussian mixture modelling (GMM) to identify latent genetic relationships between cortical measures and biochemical traits. Two distinct components (clusters) of cortical regions with respect to their biochemical correlations were identified in the TH analysis, with cluster 1 (17 regions) largely localised to the frontal and temporal lobes, while cluster 2 (17 regions) was predominantly parietal and occipital (Fig. 2b & c, Table S4). Although two clusters were also observed in the SA analysis, the spatial distribution was less clear, as cluster 1 (19 regions) contained a series of contiguous regions spanning a number of cortical lobes, whereas cluster 2 (15 regions) consisted of regions with relatively diffuse spatial localisation (Fig. 2d & e, Table S5). These results therefore suggest that genetic correlations between regional cortical structure and biochemical traits exhibit a degree of spatial organisation.

GMM applied to cortical correlation profiles of the biochemical traits failed to identify discrete clusters of biomarkers for either global SA or TH, as the optimum BIC value yielded only a single component. Therefore, we conducted GMM for the biochemical traits after combining regional SA and TH correlation profiles to capture groups of biomarkers with similar effects on both cortical properties. Interestingly, seven clusters of biochemical traits were observed (Fig. 2f, Table S6). Clusters one and six were largely composed of traits with negative LDSR *Z-*scores for regional SA and positive scores for TH, including cholesterol, testosterone and RBC-related traits, amongst others (see Fig. 2g & h for representative cluster, see Fig. S2 for full data). Clusters two, three and seven were also negatively correlated with SA and exhibited discordant correlations with TH, with consistent representation of reticulocyte and immune cell traits. Notably, cluster four was negatively correlated with both measures and included CRP, glucose, glycated haemoglobin and platelet-related traits, while cluster five was positive for SA and negative for TH. Collectively, these findings indicate that groups of cortical regions tend to have similar genetic correlations with blood-based biomarkers, while subsets of biomarkers can be classified by their relationships with both SA and TH.

### Strong evidence for causal relationships between blood-based biomarkers and regional cortical thickness

We next examined whether any correlated trait pairings exhibited evidence for partial genetic causality using a latent causal variable (LCV) model, which assumes a latent variable, *L*, mediates genetic correlation between two traits. Under this model, trait 1 is deemed partially causal for trait 2 if *L* is more strongly correlated with trait 1 than trait 2, and *vice versa*. These correlations are quantified as a posterior mean *genetic causality proportion* 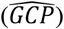, wherein 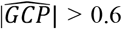 indicates evidence for partial causality between traits and the 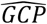 sign specifies directionality. Note that the magnitude of 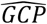 does not denote the magnitude of a causal effect, but rather implies the existence of a causal relationship. LCV models constructed for all correlated trait-pairs (*P*_*Bonferroni*_ and *P*_*BH*_ < 0.05) revealed a total of six trait pairings with strong evidence for partial genetic causality, all of which involved a biochemical trait with estimates of genetic causality on regional TH, rather than the reverse. These included: CRP on fusiform TH 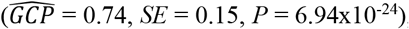, lateral occipital TH 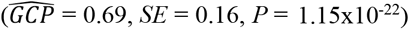 and lingual TH 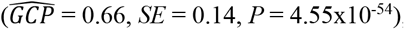, RBC count on insula TH 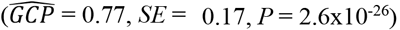, haematocrit percentage on insula TH 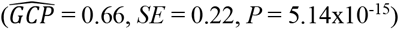 and vitamin D on temporal pole TH (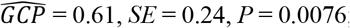 ; Fig. 3, Table 3, Table S7). To ensure these results were not affected by global measures, we repeated this analysis using GWAS summary statistics covaried for mean global cortical TH. Strong evidence for partial genetic causality remained after this adjustment for CRP on lingual TH 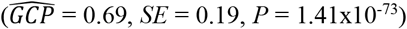 and lateral occipital TH 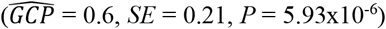 and vitamin D on temporal pole TH (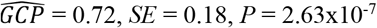; Table 3, Table S7). Considering the sign of genetic correlations between these traits, it is therefore likely that increased CRP levels are associated with decreased lingual (*r*_*g*_ = –0.118) and lateral occipital (*r*_*g*_ = –0.111) TH, whereas increased vitamin D is associated with increased temporal pole TH (*r*_*g*_ = 0.169). The evidence implicating the remaining three pairings were ablated after correction for global TH: CRP on fusiform TH 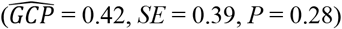, RBC count on insula 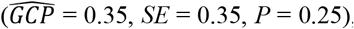, and haematocrit percentage on insula TH (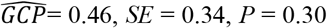 ; Table 3, Table S7). Modest 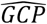 estimates were also obtained for these biochemical markers with respect to global cortical TH: CRP 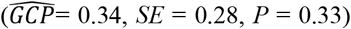, RBC count 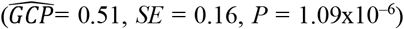, and haematocrit percentage(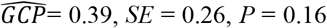 ; Table S8). We thus suspect the strong 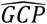 estimates obtained for these biochemical traits with respect to uncorrected regional TH GWAS quantified the combined effect on regional and global TH.

**Figure 3.**
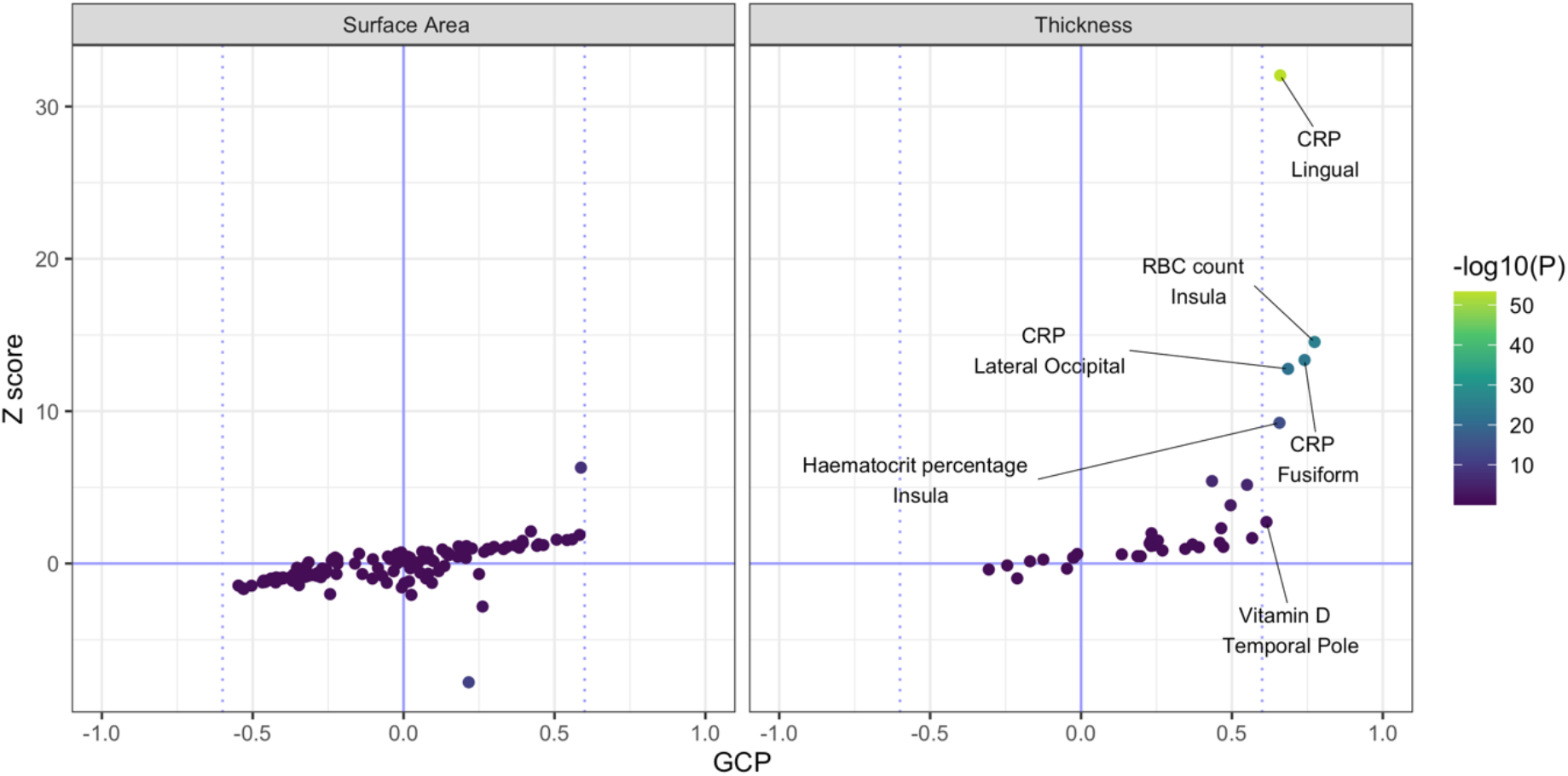
Genetically inferred causal relationships amongst biochemical and cortical traits. Biochemical-cortical trait pairings with suggestive (*P*_*BH*_ < 0.05) or significant (*P*_*Bonferroni*_ < 0.05) genetic correlation were subjected to latent causal variable (LCV) analysis. Of 35 tested trait pairings associated with regional cortical TH, six exhibited strong evidence 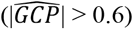 for a causal relationship, with all involving a biochemical trait acting on the cortical trait. In contrast, no trait pairings associated with cortical SA exhibited strong evidence for causality.

**Table 3.** Latent casual variable analyses for trait pairings with significant evidence for a causal relationship.

| Biochemical trait | Cortical trait | Without global TH correction |  |  |  | With global TH correction<sup>1</sup> |  |  |  |
| --- | --- | --- | --- | --- | --- | --- | --- | --- | --- |
| | | $\widehat{GCP}^2$ | $P^3$ | $SE$ | $r_g^4$ | $\widehat{GCP}$ | $P$ | $SE$ | $r_g$ |
| C-reactive protein | Lateral occipital TH | 0.69 | $1.15 \times 10^{-22}$ | 0.16 | -0.11 | 0.60 | $5.93 \times 10^{-6}$ | 0.21 | -0.05 |
| C-reactive protein | Lingual TH | 0.66 | $4.55 \times 10^{-54}$ | 0.14 | -0.12 | 0.69 | $1.41 \times 10^{-73}$ | 0.19 | -0.07 |
| C-reactive protein | Fusiform TH | 0.74 | $6.94 \times 10^{-24}$ | 0.15 | -0.17 | 0.42 | 0.28 | -0.39 | -0.15 |
| Haematocrit % | Insula TH | 0.66 | $5.14 \times 10^{-15}$ | 0.22 | 0.12 | 0.46 | 0.30 | 0.34 | 0.05 |
| RBC count | Insula TH | 0.77 | $2.60 \times 10^{-26}$ | 0.17 | 0.12 | 0.35 | 0.25 | 0.35 | 0.06 |
| Vitamin D | Temporal pole TH | 0.61 | $7.61 \times 10^{-03}$ | 0.24 | 0.17 | 0.72 | $2.63 \times 10^{-7}$ | 0.18 | 0.22 |

<sup>1</sup>Denotes LCV analyses conducted using GWAS summary statistics produced with mean global cortical TH used as a covariate.
<sup>2</sup>The posterior mean genetic causality proportion.
<sup>3</sup> $P$ value which tests whether the $\widehat{GCP}$ estimate is significantly different from zero.
<sup>4</sup>Genetic correlation coefficient.

### C-reactive protein and vitamin D are associated with regional cortical thickness

The total effect of CRP levels (natural log transformed mg/L) on lingual and lateral occipital TH (in mm) was next quantified via two-sample Mendelian randomisation (MR). Due to the inclusion of UKBB samples in the ENIGMA metanalysis, we obtained instrumental variables (IVs) for CRP utilising summary statistics sourced from a non-UKBB cohort [21]. Furthermore, we employed an inverse variance weighted estimator with multiplicative random effects (IVW_m_) as our primary test, followed by four other methods with different underlying assumptions regarding IV validity. Across all analyses, a total of 48 CRP-associated IVs were identified after harmonisation and removal of palindromic SNPs. Interestingly, we identified evidence suggesting each natural log transformed mg/L increase in CRP was associated with a significant reduction in lingual (*β*_*IVWm*_= –0.009, *SE*_*IVWm*_ = 0.003, *P*_*IVWm*_ = 0.004) and lateral occipital (*β*_*IVWm*_ = –0.008, *SE*_*IVWm*_ = 0.003, *P*_*IVWm*_ = 0.025) TH, consistent with LDSR and LCV results (Fig. 4a & c, Table 4, Table S9). Using the less conservative IVW estimator with fixed effects decreased the standard error of those estimates, in line with expectation (*P*_*lingual*_ = 1.39×10^−4^, *P*_*lateral occipital*_ = 0.004). However, the weighted median, weighted mode and MR Egger models, whilst directionally consistent, generally did not achieve statistical significance, with the exception of the weighted mode model for CRP on lingual TH (*β* = –0.007, *SE* = 0.003, *P* = 0.04; Fig. 4a & c, Table 4, Table S9). We additionally examined these relationships after covarying for global cortical TH and found the causal estimate for CRP on lingual TH remained significant (*β*_*IVWm*_ = –0.007, *SE*_*IVWm*_ = 0.002, *P*_*IVWm*_ = 0.003), whereas the effect on lateral occipital TH was marginally attenuated (*β*_*IWVm*_ = –0.004, *SE*_*IVWm*_ = 0.002, *P*_*IVWm*_ = 0.09; (Fig. 4b & d, Table 4, Table S9). Across all analyses, we identified no substantial evidence for outlier SNPs using MR-PRESSO and leave-one-out analyses, although the IV exposure-outcome effects exhibited evidence for SNP heterogeneity in both instances (Cochran’s *Q* ≥ 69.86, *P* ≤ 0.01; Tables S10 & S11). This heterogeneity, however, is not unexpected given the underlying biological complexity of these traits. Moreover, the Egger intercepts were not significantly different than zero (*P* ≥ 0.26), therefore providing statistical evidence of no unbalanced pleiotropy (Table S12).

**Figure 4.**
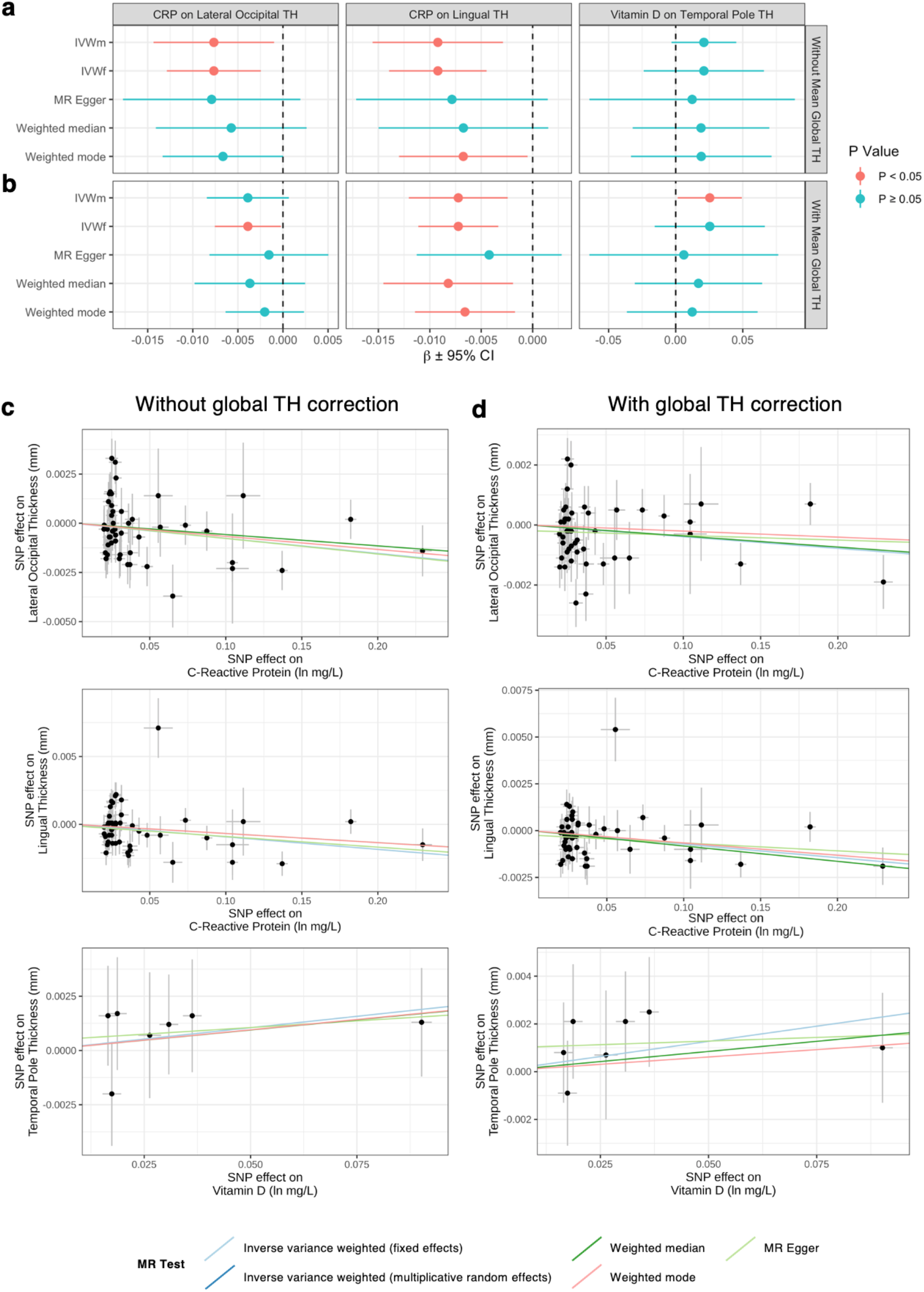
Estimated effect of C-reactive protein and vitamin D exposures on regional cortical thickness measures. **(a)** Effect size estimates (± 95% CI) of C-reactive protein and vitamin D exposures on cortical TH GWAS. **(b)** As in **(a)**, except using cortical GWAS with global mean cortical TH included as a covariate. **(c)** Scatter plots comparing IV effect sizes in exposure and outcome GWAS. Each trendline corresponds to one of five MR methods utilized for sensitivity analysis. **(d)** As in **(c)**, except using cortical GWAS corrected for global TH.

**Table 4.**
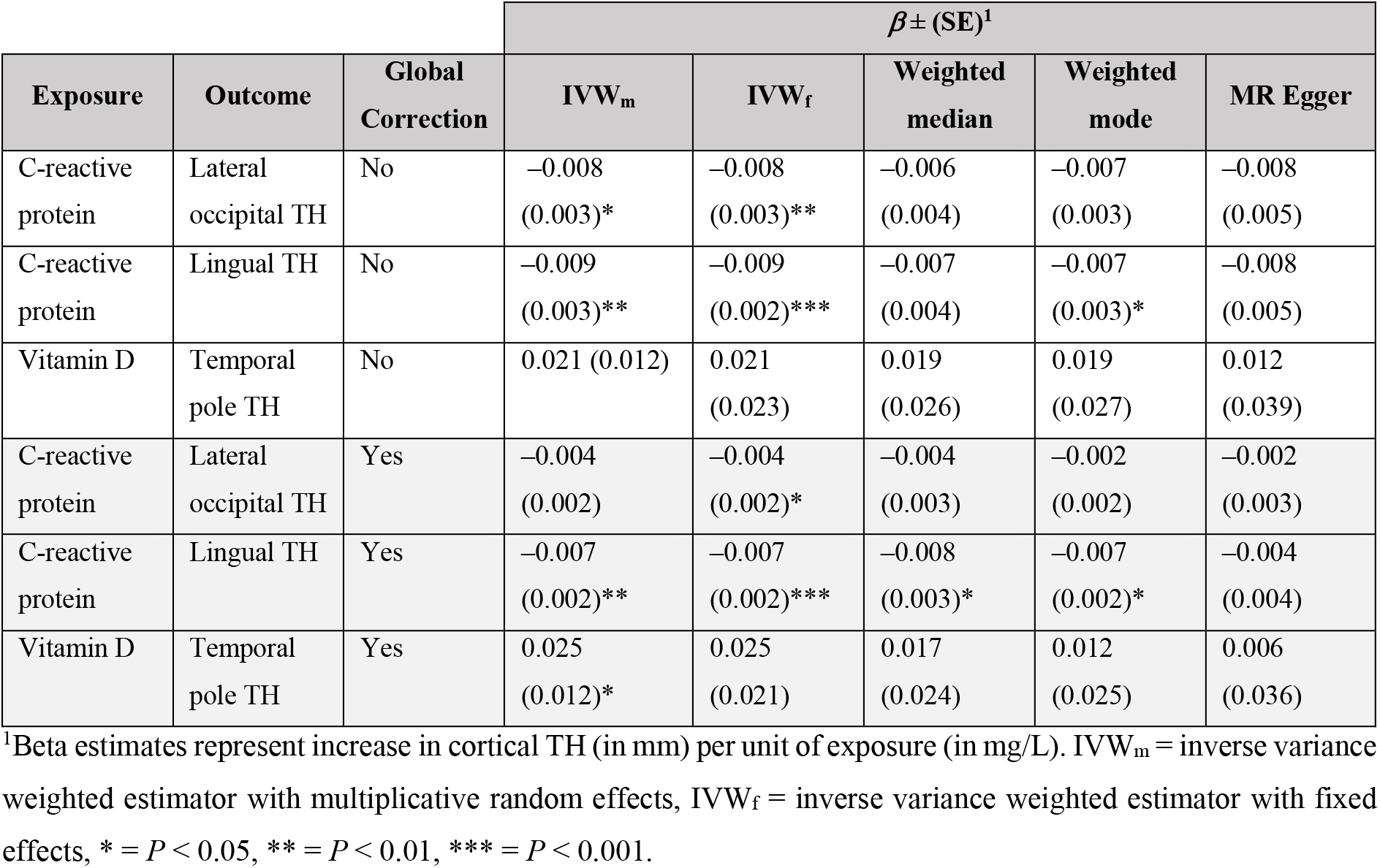
Causal relationships between biochemical traits and regional cortical TH estimated via two-sample Mendelian randomisation.

| Exposure | Outcome | Global Correction | $\beta \pm (SE)^1$ | | | | |
| --- | --- | --- | --- | --- | --- | --- | --- |
|  |  |  | IVW<sub>m</sub> | IVW<sub>f</sub> | Weighted median | Weighted mode | MR Egger |
| C-reactive protein | Lateral occipital TH | No | -0.008<br>(0.003)* | -0.008<br>(0.003)** | -0.006<br>(0.004) | -0.007<br>(0.003) | -0.008<br>(0.005) |
| C-reactive protein | Lingual TH | No | -0.009<br>(0.003)** | -0.009<br>(0.002)*** | -0.007<br>(0.004) | -0.007<br>(0.003)* | -0.008<br>(0.005) |
| Vitamin D | Temporal pole TH | No | 0.021 (0.012) | 0.021<br>(0.023) | 0.019<br>(0.026) | 0.019<br>(0.027) | 0.012<br>(0.039) |
| C-reactive protein | Lateral occipital TH | Yes | -0.004<br>(0.002) | -0.004<br>(0.002)* | -0.004<br>(0.003) | -0.002<br>(0.002) | -0.002<br>(0.003) |
| C-reactive protein | Lingual TH | Yes | -0.007<br>(0.002)** | -0.007<br>(0.002)*** | -0.008<br>(0.003)* | -0.007<br>(0.002)* | -0.004<br>(0.004) |
| Vitamin D | Temporal pole TH | Yes | 0.025<br>(0.012)* | 0.025<br>(0.021) | 0.017<br>(0.024) | 0.012<br>(0.025) | 0.006<br>(0.036) |

<sup>1</sup>Beta estimates represent increase in cortical TH (in mm) per unit of exposure (in mg/L). IVW<sub>m</sub> = inverse variance weighted estimator with multiplicative random effects, IVW<sub>f</sub> = inverse variance weighted estimator with fixed effects, \* = $P < 0.05$ , \*\* = $P < 0.01$ , \*\*\* = $P < 0.001$ .

We also analysed the effect of natural log transformed vitamin D (as 25-hydroxyvitamin D, nmol/L) concentrations on temporal pole TH using GWAS summary statistics from the SUNLIGHT consortium [22], from which seven harmonised, non-palindromic IVs were identified. Interestingly, while serum vitamin D initially exhibited a positive trend with respect to temporal pole TH (*β*_*IVWm*_ = 0.021, *SE*_*IVWm*_ = 0.012, *P*_*IVWm*_ = 0.088), this relationship actually attained statistical significance after covarying for global TH (*β*_*IVWm*_ = 0.025, *SE*_*IVWm*_ = 0.012, *P*_*IVWm*_ = 0.038; Fig 4, Table 4, Table S9). For both analyses, we observed no evidence for outlying SNPs, confounding pleiotropy (*P*_*EI*_ ≥ 0.54) nor heterogeneity (Cochran’s *Q* ≤ 2.04, *P* ≥ 0.89; Tables S10–S12). We would posit that the attenuated significance in the relationship between vitamin D and temporal pole TH using an MR construct relative to the LCV model is not directly comparable, given that MR exploits genome-wide SNP effects as IVs rather than modelling genome-wide trait effects.

Finally, trait pairings which did not survive global correction in the LCV analysis (i.e. CRP → fusiform TH, RBC count → insula TH and haematocrit percentage → insula TH) were examined to determine whether any causal relationships could be resolved through IVs. For RBC count and haematocrit percentage, 26 and 11 IVs were respectively identified from GWAS sourced from the INTERVAL consortium [36], ensuring UKBB samples were excluded. Limited evidence for causal relationships was identified across all pairings, with only RBC count exhibiting evidence for a causal effect on insula TH without correction for global TH (*β*_*IVWm*_ = 0.01, *SE*_*IVWm*_ = 0.005, *P*_*IVWm*_ = 0.025), thus generally supporting the LCV findings (Table S9).

### Limited transcriptome-wide overlap amongst causally linked trait pairings

Transcriptome-wide association studies were performed for CRP, vitamin D and their respective causally associated cortical phenotypes to compare and contrast predicted gene expression profiles. Briefly, this approach utilises tissue-specific *cis*-eQTL expression weights to impute gene expression profiles associated with the phenotype of interest. We specifically employed pre-computed SNP-expression weights for cortical tissue and whole blood, while liver expression weights were additionally included as CRP is hepatically synthesised (see Table S13 for full results). At a *P*_*Bonferroni*_ < 0.05 cut-off, the average number of associated genes per trait across all tissues was as follows: CRP = 157, vitamin D = 37, lateral occipital TH = 1, lingual TH = 4 and temporal pole TH = 2 (see Fig. 5a for representative Miami plots, see Table S14 for full results). Given the sparse identification of associated genes for the cortical traits, no overlapping genes were identified (Table 5, Table S15). However, after correcting for global TH, *RP11-148021*.*6*, was found to overlap between CRP (*Z*_*TWAS*_ = 6.03) and lateral occipital TH (*Z*_*TWAS*_ = –4.46) using cortical expression weights (Table 5, Table S14 & S15). Utilising a more-liberal Benjamini-Hochberg significance cut-off, genes overlapping CRP and lingual TH were identified as follows: cortical weights: *KRT18P34, PPFIA1, RP11-555M1*.*3*, whole blood weights: *KRT18P34, PPFIA1, ZNF660* and liver weights: *PPFIA1* (Table 5, Table S14 & S15). Interestingly, *PPFIA1*, which encodes the synaptic scaffolding protein Liprin-α-1 [37], was represented across all three tissues with consistently negative *Z-*scores in the CRP TWAS and positive *Z-*scores in the lingual TH TWAS. To more-broadly explore transcriptomic overlap amongst trait pairings with evidence for causality, we utilised *RHOGE* to estimate transcriptome-wide correlation (*ρ*_*GE*_). For all pairings and tissues, no significant correlations were identified after multiple testing correction (Fig. 5b, Table S16), however, at a nominal *P* < 0.05 cut-off vitamin D and temporal pole TH were positively correlated after correction for global measures using cortical weights (*ρ*_*GE*_ = 0.45, *P* = 0.023, *SE* = 0.187). Collectively, these findings suggest there is minimal predicted transcriptomic overlap between causal biochemical-cortical trait pairings. However, we also identified only a limited number of transcriptome-wide associated signals for the cortical properties suggesting that greater sample sizes are required, as well as more tissue and cell-type specific expression weights.

**Figure 5.**
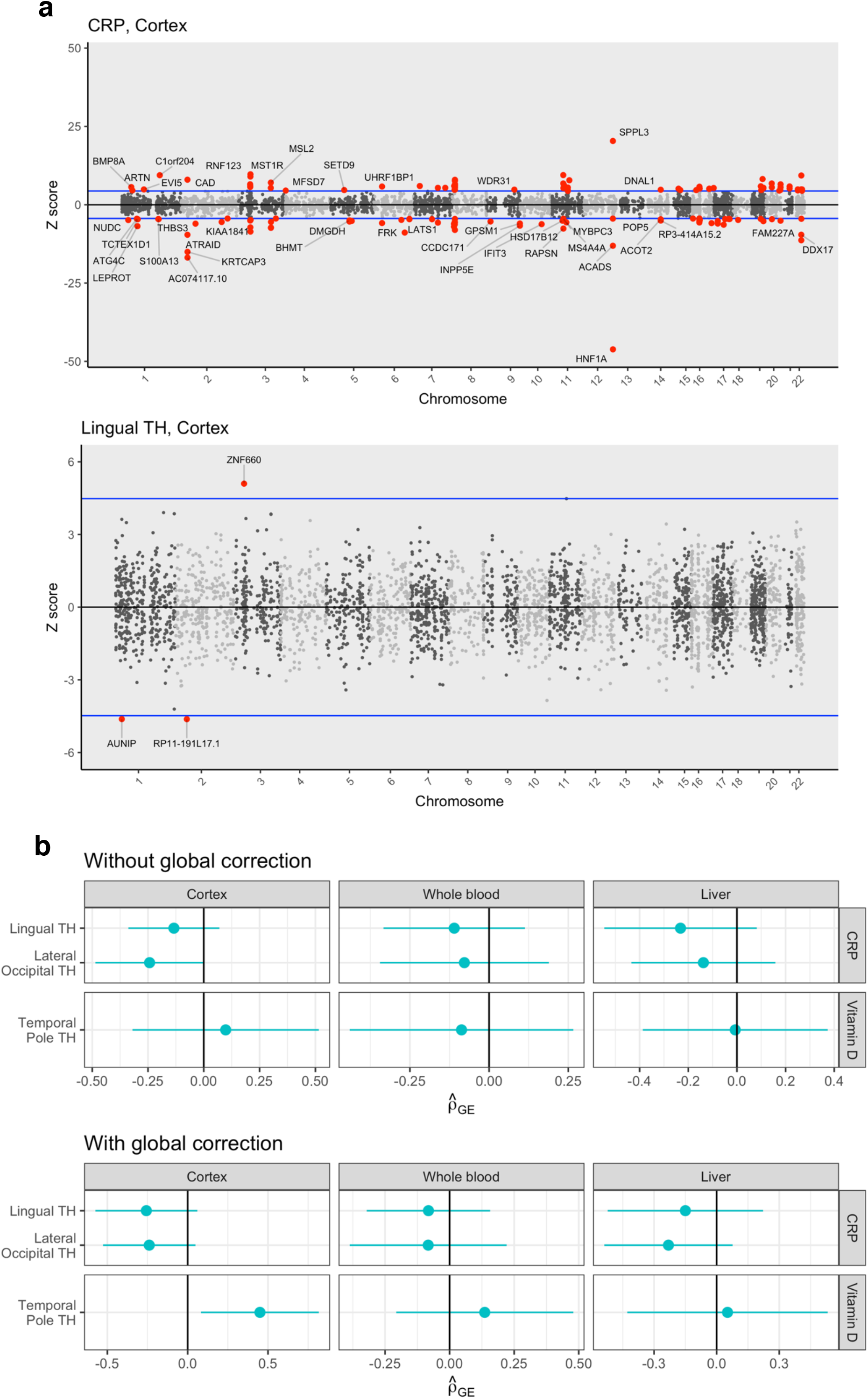
Transcriptome-wide correlation of TWAS *Z-*score profiles between biochemical and cortical traits. **(a)** Miami plots depicting TWAS results for two representative traits – CRP and lingual TH (without correction for global TH) – using cortical expression weights. Horizontal blue lines correspond to *P*_*Bonferroni*_ < 0.05. **(b)** Correlation of TWAS *Z-*score profiles amongst all trait pairings with evidence for a causal relationship using RHOGE. Across all tissues, no trait pairings were significantly correlated after multiple testing correction, however the direction of *ρ*_*GE*_ was generally consistent with genetic correlations. Note that temporal pole TH and vitamin D expression profiles were nominally correlated (*ρ*_*GE*_ = 0.45, *P* = 0.023, *SE* = 0.187) after correction for global TH and using cortical weights.

**Table 5.**
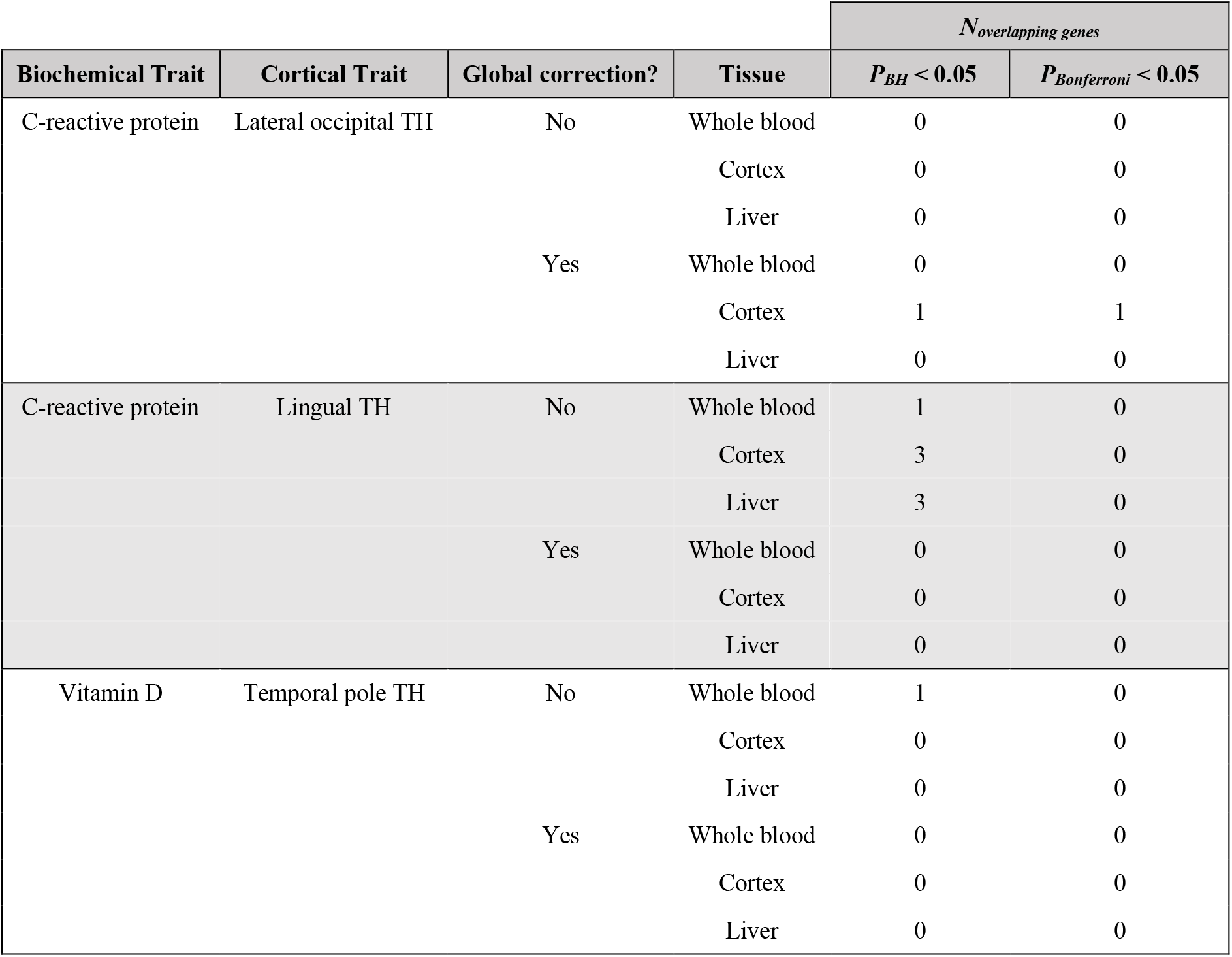
Overlapping genes between biochemical and cortical TWAS.

| Biochemical Trait | Cortical Trait | Global correction? | Tissue | *N* overlapping genes |  |
| --- | --- | --- | --- | --- | --- |
| | | | | $P_{BH} < 0.05$ | $P_{Bonferroni} < 0.05$ |
| C-reactive protein | Lateral occipital TH | No | Whole blood | 0 | 0 |
|  |  |  | Cortex | 0 | 0 |
|  |  |  | Liver | 0 | 0 |
|  |  | Yes | Whole blood | 0 | 0 |
|  |  |  | Cortex | 1 | 1 |
|  |  |  | Liver | 0 | 0 |
| C-reactive protein | Lingual TH | No | Whole blood | 1 | 0 |
|  |  |  | Cortex | 3 | 0 |
|  |  |  | Liver | 3 | 0 |
|  |  | Yes | Whole blood | 0 | 0 |
|  |  |  | Cortex | 0 | 0 |
|  |  |  | Liver | 0 | 0 |
| Vitamin D | Temporal pole TH | No | Whole blood | 1 | 0 |
|  |  |  | Cortex | 0 | 0 |
|  |  |  | Liver | 0 | 0 |
|  |  | Yes | Whole blood | 0 | 0 |
|  |  |  | Cortex | 0 | 0 |
|  |  |  | Liver | 0 | 0 |

## DISCUSSION

Circulating biochemical factors play a critical role in the development and homeostasis of the central nervous system, however it is unclear whether individual biochemical exposures can modulate structural characteristics of specific brain regions. In the present study, we investigated shared genetic architecture and causal relationships between a panel of blood-based biochemical exposures and cortical structural properties with direct relevance to psychiatric disorders. Our analyses revealed marked genetic overlap between biochemical exposures and structural measures for discrete cortical regions which loosely clustered by their spatial proximity, suggesting subsets of genetic variants associated with biochemical traits are also associated with regional variations cortical structure. In addition, minimal genetic correlation was identified for global cortical SA and TH, further supporting region-specific organisation of cortical genetic architecture.

Interestingly, we observed clear divergence in the direction of correlations for regional SA (95.8% negative) and TH (72.7% positive), which was particularly evident after clustering of biochemical traits by their cortical LDSR *Z-*score profiles. Of seven identified biochemical clusters, only cluster four (CRP, glucose, glycosylated haemoglobin, platelet count and platelet crit) exhibited SA and TH *Z-*scores of the same sign and similar magnitude, whereas discordant correlation profiles were observed for the remaining biochemical exposures. Genetic comparison of cortical SA and TH has previously revealed these characteristics are genetically independent [38] and exhibit discordant profiles of genetic correlation with respect to neurobehavioral traits and psychiatric disorders [15], implying these structural properties are regulated via independent biological mechanisms. This is consistent with neuroanatomical studies reporting a range of concordant and discordant relationships between region-specific SA and TH with age [39-41] and in the context of psychiatric disorders [42, 43]. Future studies should thus focus on characterisation of genetic variants associated with biochemical traits with extensive genetic correlations (for example, white blood cell count, red blood cell count and haemoglobin) and their impact on cortical SA and TH to further reconcile biochemical exposures with their predicted associations with cortical structure. Although many biochemical traits genetically correlated with cortical structure failed to exhibit strong evidence for a causal relationship, we suspect the combined effect of these exposures may mediate biologically significant cortical alterations, given subsets (such as triglycerides [44] and haemoglobin [35]) have been previously associated with cortical integrity.

By utilising genetically informed methods of causal inference, evidence for a putative causal relationship was identified for CRP on lingual and lateral occipital thinning. CRP is widely characterised as an acute inflammatory protein synthesised by hepatocytes as a soluble homopentamer (pCRP) in response to IL-6 and IL-1β signalling [45]. After biosynthesis, pCRP enters systemic circulation and dissociates into proinflammatory monomers (mCRP) at the site of insult, which increase the abundance of M1 macrophages and Th1 T-cells, thereby activating a robust immune response with potentially deleterious effects on host tissues [46]. Persistent elevation of CRP is therefore strongly associated with chronic, low-level inflammation thought to negatively impact the structural integrity of brain regions. For instance, elevated CRP is positively correlated with cortical thinning [9, 47] and decreased grey matter volume [48, 49], while deposition of mCRP has been identified at sites of neurodegeneration [50]. Although CRP-mediated thinning of the lingual and lateral occipital regions has not been explicitly reported in previous studies, recent work has shown elevated CRP is associated with lower regional cerebral blood flow to the lingual gyrus, suggesting alteration of cerebral vasculature may contribute to spatially-restricted cortical thinning [51]. Despite this extensive observational evidence, the exact mechanisms through which CRP alters cortical structure remains unclear due to difficulties in dissecting casual relationships via observational studies. Whilst we utilised TWAS to identify common genes amongst CRP, lingual TH and lateral occipital TH to explore shared pathways with potential biological significance, few genes were significantly associated with the cortical measures across all tested tissues, limiting our capacity to genetically probe the mechanistic basis through which CRP modulates cortical integrity in a region-specific manner. This could likely be improved in future studies by boosting the discovery GWAS sample size and specificity of brain regions available, particularly as the FUSION approach is limited to genes with a significantly heritable *cis*-acting component. Moreover, the genetic overlap between these cortical and biochemical traits may not be heavily mediated by *cis*-acting influences on mRNA expression, and thus, integration of other functional data, such as chromatin and methylome related annotations, could yield more genes associated with these traits. However, we did identify consistent representation of *PPFIA1* across all three tissues, with negative *Z-*scores in the CRP TWAS and positive *Z-*scores in the lingual TH TWAS. This gene encodes Liprin-α-1, a synaptic scaffolding molecule associated with axon targeting, synaptic morphogenesis synaptic vesicle transport and recruitment of AMPA receptors [37], thus genetically mediated regulation of this gene may contribute to the interrelationship between CRP and lingual TH. We additionally note that reverse causality is unlikely for these putative relationships given the strong directional evidence identified via LCV.

Mounting observational evidence suggests psychiatric disorders including schizophrenia, major depressive disorder and bipolar disorder are associated with elevated CRP levels [9, 52, 53]. In the case of schizophrenia, high peripheral CRP is correlated with negative symptoms [54] as well as cortical thinning in frontal, insula and temporal regions [9]. However, no studies to date have specifically reported CRP-associated thinning of the lingual and lateral occipital regions, nor CRP-associated alteration of their accompanying cognitive functions, such as visual memory (lingual), visual imagery (lingual) and object recognition (lateral occipital) [55, 56]. Thus, it is unclear whether the putative causal relationships identified in the current study bear direct functional significance for psychiatric disorders. More-generally, conflicting results from MR and LCV analyses have recently reported a protective effect for CRP in schizophrenia, obsessive compulsive disorder and anorexia nervosa, and a causal effect in major depressive disorder [13, 57], further obscuring the underlying role of this protein in psychiatric illness. While these discoveries seemingly conflict with our results and previous observational evidence, complex dimensions of CRP function may underpin these divergent findings. Specifically, recent evidence suggests pCRP can dampen inflammation by skewing macrophages and T-cells towards anti-inflammatory M2 and Th2 phenotypes [46], respectively, suggesting that while high serum pCRP may be indicative of a pervasive acute phase inflammatory response, moderately elevated pCRP may indeed exert a protective effect against underlying chronic inflammation. This is an issue for observational inferences in particular, which employ tests such as the mean difference in CRP between cases and controls, as the magnitude of CRP elevation can often be relatively small and more indicative of this moderate state. Regardless of the underlying mechanism, these findings collectively advocate further investigation of the direct and indirect effects of CRP on neuronal integrity to further reconcile the impact of CRP levels on cortical structure and psychiatric disorders.

We additionally identified a positive causal relationship for vitamin D on temporal pole TH after correcting for global TH. A nominally significant transcriptome-wide correlation between these traits was also observed after correcting for global TH. Together, these findings suggest any effect of vitamin D was highly specific to the temporal pole, broadly consistent with a recent cross-sectional study wherein total vitamin D intake and vitamin D supplementation were specifically associated with enhanced temporal lobe TH in cognitively normal, older adults (>65 years) [58]. However, similar studies have previously reported increased prefrontal and cingulate TH in individuals with higher serum vitamin D concentrations [59, 60], suggesting any protective effect of vitamin D may not be spatially restricted to structures such as the temporal pole. Indeed, vitamin D receptors and other vitamin D associated genes (such as *CYP27B1*) are expressed throughout the brain [61], while functional studies indicate vitamin D promotes axon outgrowth [62], enhances expression of neurotrophic factors (such as NGF) [62], and exhibits neuroprotective effects with respect to excessive calcium, reactive oxygen species and corticosterone [63], indicating a particularly important role throughout the brain. While only one significant relationship was identified for vitamin D in the present study, we consequently suspect protective effects of vitamin D on cortical regions other than the temporal pole may be mediated through complex multivariate interactions not readily detectable in our analyses.

The proposed relationship between vitamin D and temporal pole TH is nonetheless significant for general cognition and psychiatric illness given the diverse range of functions attributed to this region, including: visual cognition, face recognition, visual memory, visual discrimination, social cognition, language and semantic processing and autobiographic memory, amongst others [64, 65]. Subsets of these cognitive functions are indeed disrupted in schizophrenia and other psychiatric disorders [66-68], and furthermore, structural alterations affecting the temporal pole have also been reported in this setting [65]. In schizophrenia, temporal pole thinning is correlated with clinically relevant factors such as normalised medication dose, symptom severity and duration of illness, and negatively correlated with age of onset [69]. Reduced temporal pole volume has additionally been reported in as obsessive compulsive disorder [70], bipolar disorder [71], major depressive disorder [72] and attention deficit hyperactive disorder [73], indicating consistent dysregulation amongst a range of psychiatric disorders. In conjunction with these findings, low neonatal and adult vitamin D levels have been previously identified as a risk factor for schizophrenia [74, 75], however it is unclear whether low vitamin D directly contributes to schizophrenia-associated alterations to cortical structures such as the temporal pole. Furthermore, any causal impacts of vitamin D alterations in psychiatric disorders are inherently difficult to observe, given that affected individuals often have poorer general health and diets, while results from randomised controlled trials assessing the effect of vitamin D supplementation in schizophrenia have reported conflicting results [76]. Whilst promising large-scale randomised controlled trials [77] are expected to further elucidate a potential role for therapeutic vitamin D supplementation in schizophrenia, ongoing studies examining the role of neonatal vitamin D deficiency on brain development and schizophrenia risk [76] will likely prove critical in mapping the impact of vitamin D in development of key cortical regions such as the temporal pole.

In summary, our findings suggest subsets of biochemical exposures and cortical structural properties share genetic architecture and, in some cases, exhibit evidence for causal relationships. We acknowledge a number of limitations and caveats with respect to interpretation of the presented data. Firstly, all analyses in the current study are inherently subject to limitations and biases potentially associated with the utilised summary statistics, such as population stratification [78] and selection bias [79]. The UKBB cohort in particular is predominantly composed individuals over the age of 40, thus age-related modulation of variants may affect genetic correlations and causal estimates. Secondly, while many of the reported genetic correlations likely reflect shared genetic architecture, horizontal pleiotropy may mediate associations between some trait pairings, however we note these are still useful in guiding identification of genes that impact both biochemical traits and cortical structure. Finally, we caution that all reported causal relationships require validation in randomised controlled trials to confirm the putative causal effects. Despite the limitations of these analyses, genetically informed causal inference represents an exciting opportunity to screen and prioritise biochemical traits *en masse* to guide future investigation of these exposures in the context of neuronal function, brain cytoarchitecture and psychiatric illness.

## Data Availability

All GWAS summary statistics are freely available without restriction, with the source of each GWAS fully described in the manuscript.

## ACKNOWLEDGEMENTS

This work was supported by a National Health and Medical Research Council (NHMRC) grants (1147644, 1188493). M.J.C. is supported by an NHMRC Senior Research Fellowship (1121474), URL: https://www.nhmrc.gov.au/ and a University of Newcastle College of Health Medicine and Wellbeing, Gladys M Brawn Senior Fellowship, URL: https://www.nhmrc.gov.au/. The funders had no role in study design, data collection and analysis, decision to publish, or preparation of the paper. W.R.R was supported by an Australian Government Research Training Program Stipend.

## AUTHOR CONTRIBUTIONS

D.J.K. conceived and designed the study with input from W.R.R and M.J.C. D.J.K. conducted the primary analyses and composed the manuscript. W.R.R provided methodological insight, interpretation of the results and assisted with drafting and editing of the manuscript. M.J.C. provided funding, contributed to interpretation of the results and edited the manuscript.

## DATA AND CODE AVAILABILITY

GWAS summary statistics for the ENIGMA cortical measures are available upon request at http://enigma.ini.usc.edu/research/download-enigma-gwas-results/. GWAS summary statistics for the UKBB biochemical traits (round 2) are available for download at http://www.nealelab.is/uk-biobank. Non UKBB GWAS for CRP (ieu-b-35) and vitamin D (ebi-a-GCST005367) were sourced from https://gwas.mrcieu.ac.uk, while RBC count (GCST004601) and haematocrit percentage (GCST004604) were downloaded from GWAS Catalog (https://www.ebi.ac.uk/). Code utilized in this study is available upon request.

