## Supplementary Figures for "Causal effect of C-reactive protein and vitamin D on human cerebral anatomy observed among genetically correlated biomarkers in blood"

Contents

Figure S1. Correlation of biochemical traits via comparison of LDSR cortical *Z* score profiles.	2.

Figure S2. Cortical *Z* score distributions across all biochemical trait clusters.	3–5.

Figure S3. Miami plots of TWAS results.	6–15.

Figure S4. Forest plots of all RHOGE results.	16–17.

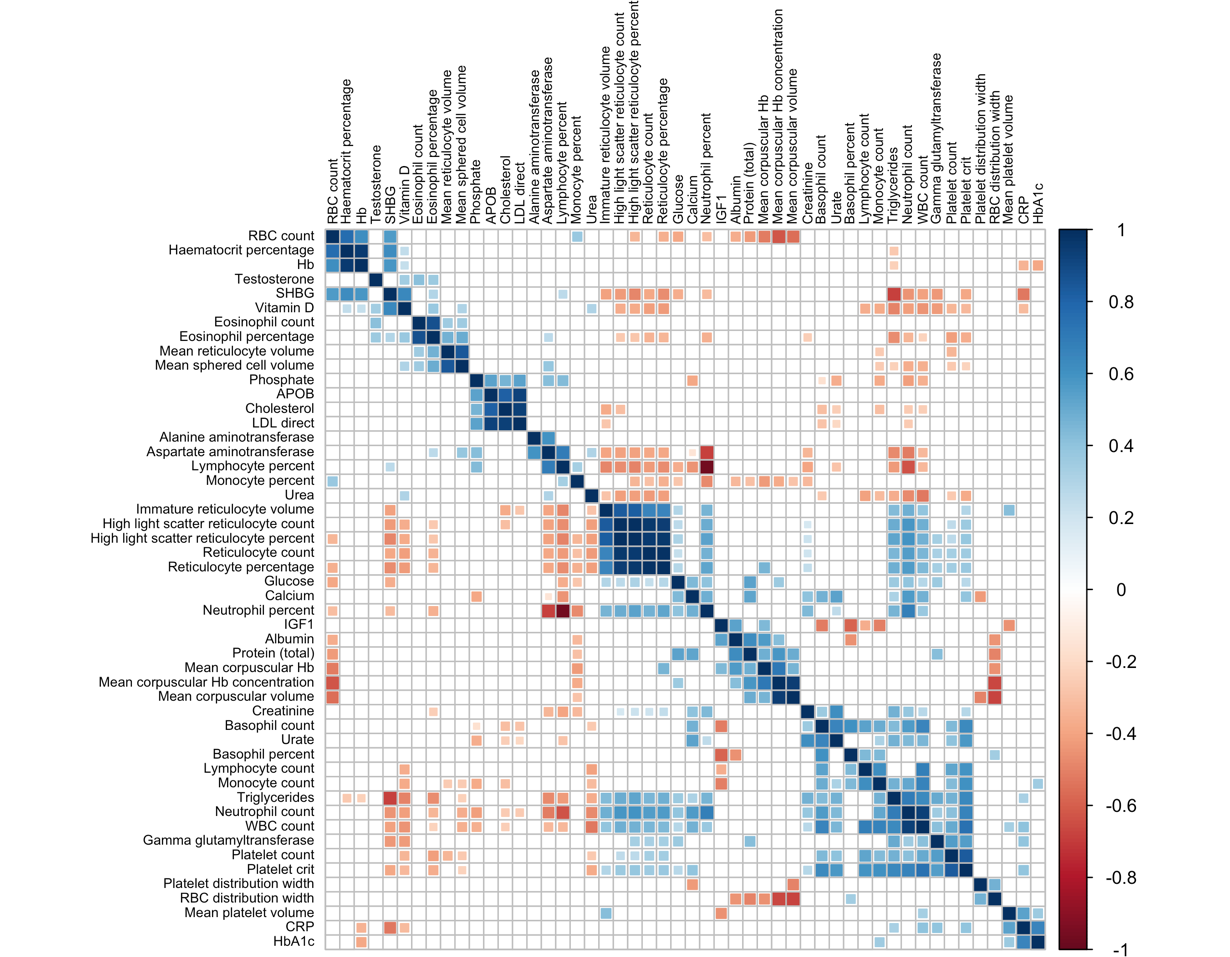

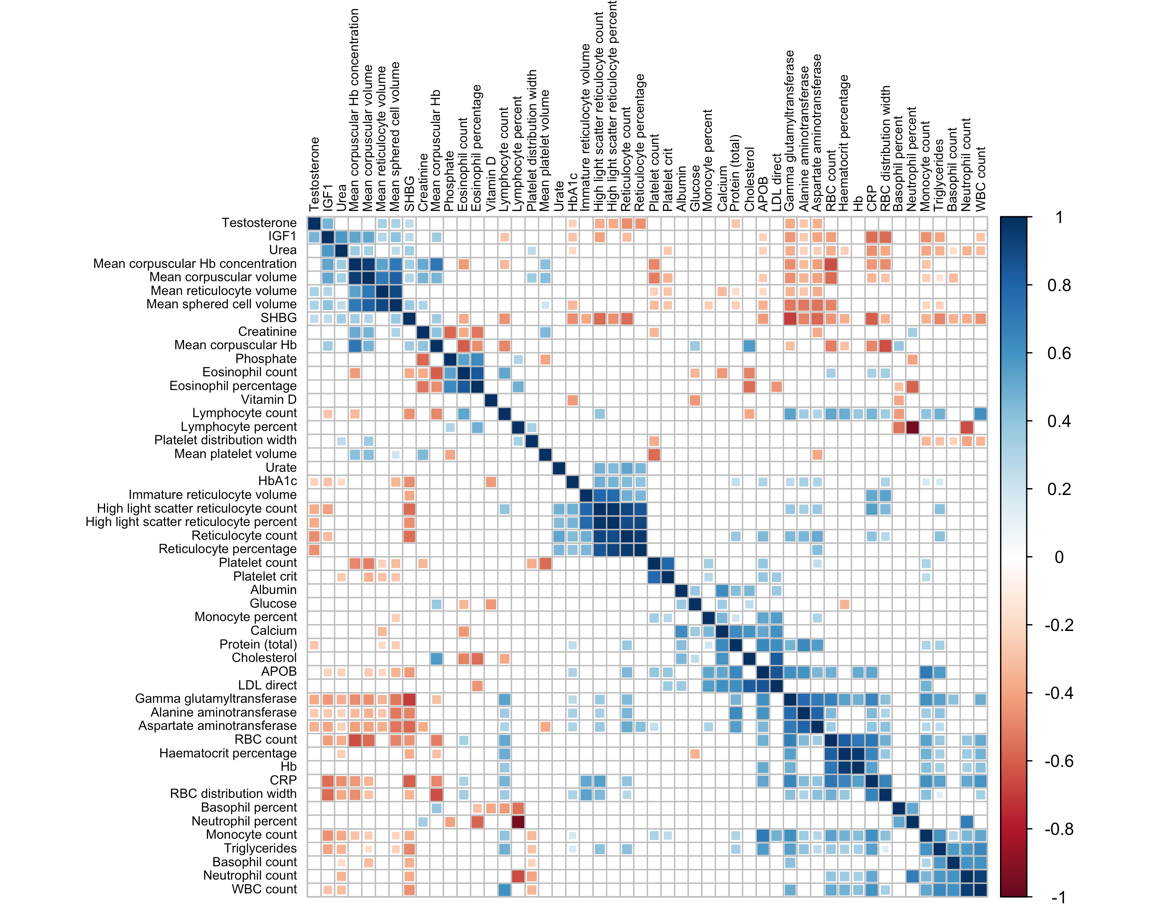

**Surface Area**

**Thickness**

**Figure S1. Correlation of biochemical traits via comparison of LDSR cortical *Z-*score profiles.** Heatmaps depicting correlations between biochemical traits after comparison of LDSR cortical *Z-*scores. Biochemical trait pairings with *P_Bonferroni_* < 0.05 are shown, with hierarchal clustering via Ward’s D2 method.

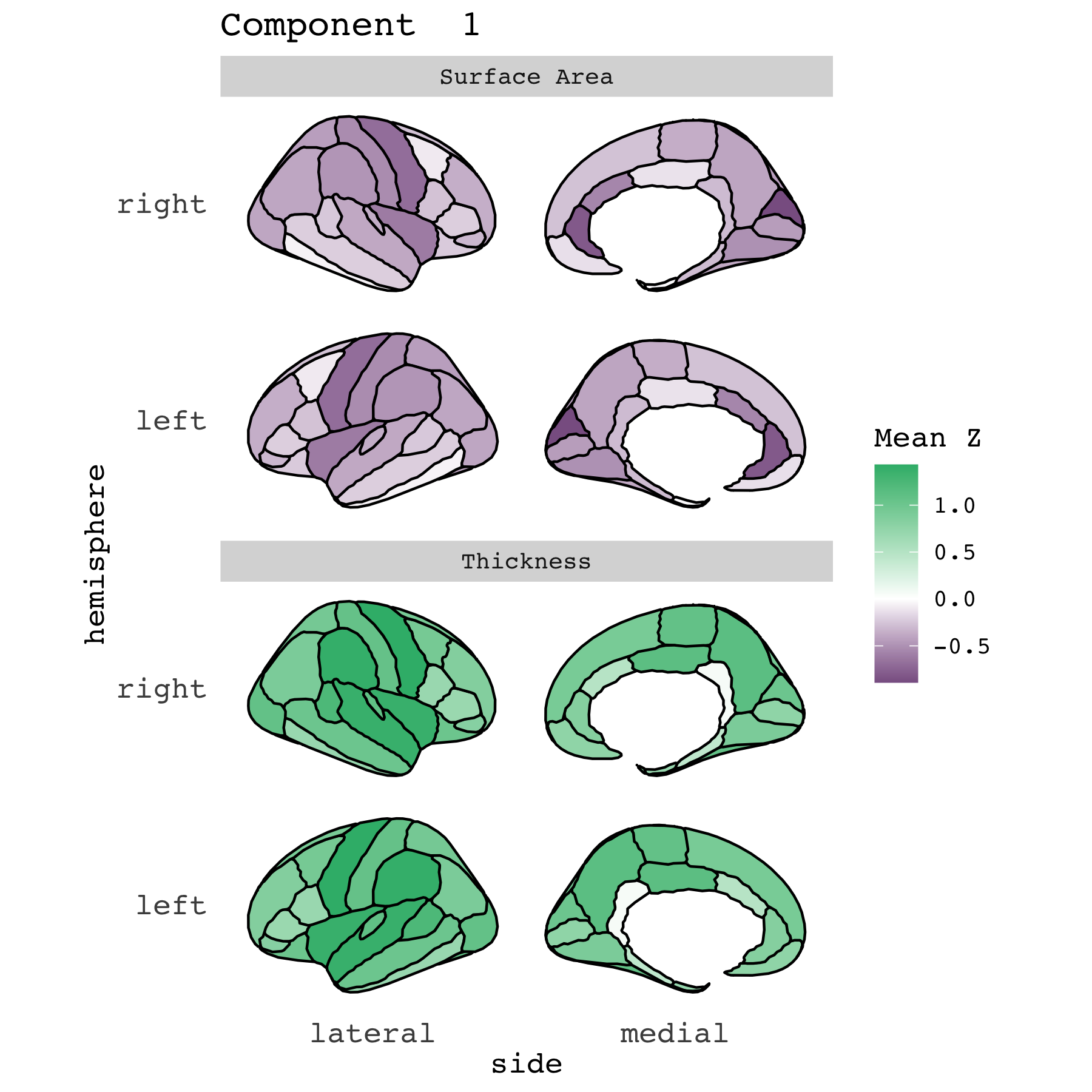

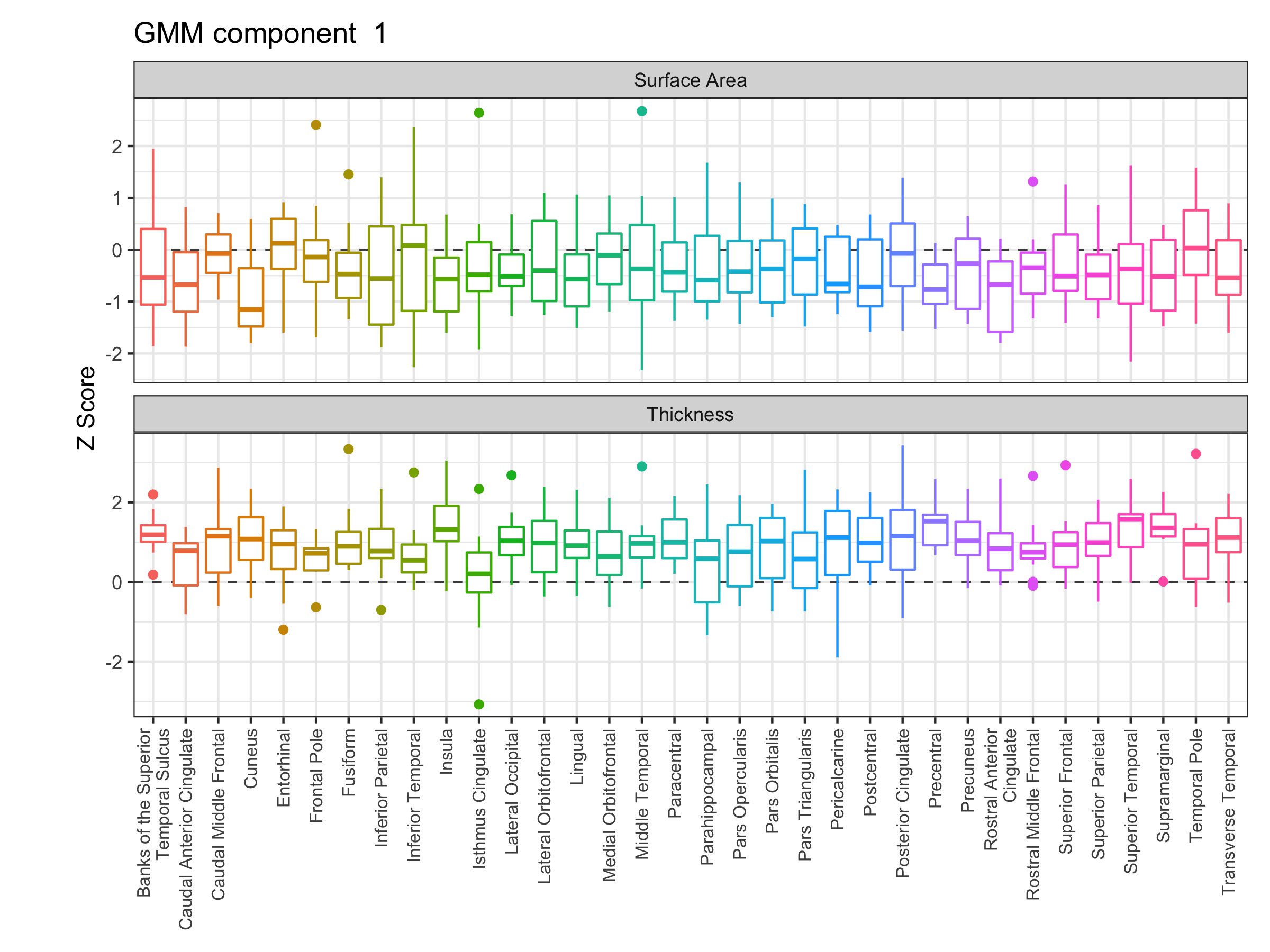

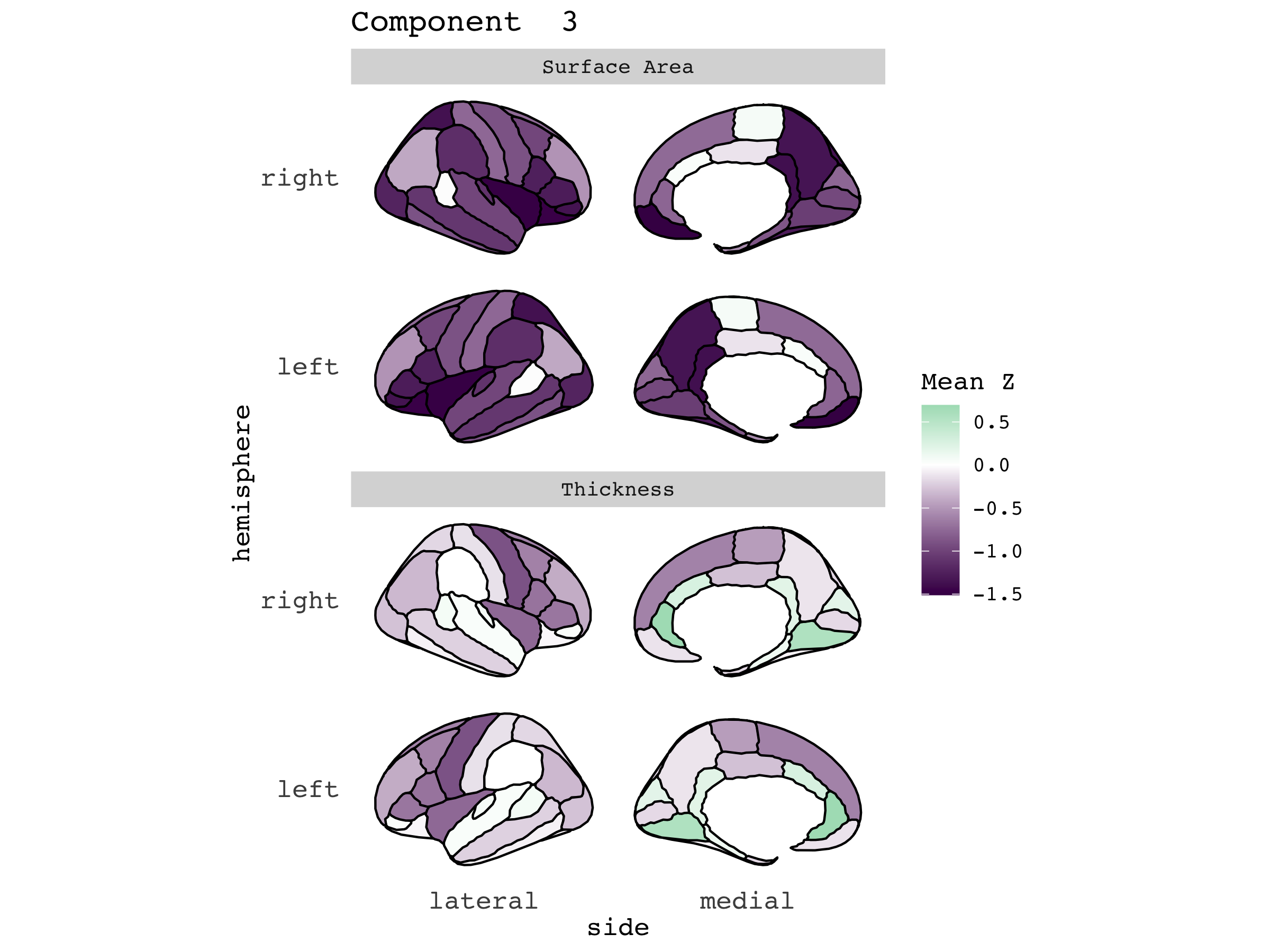

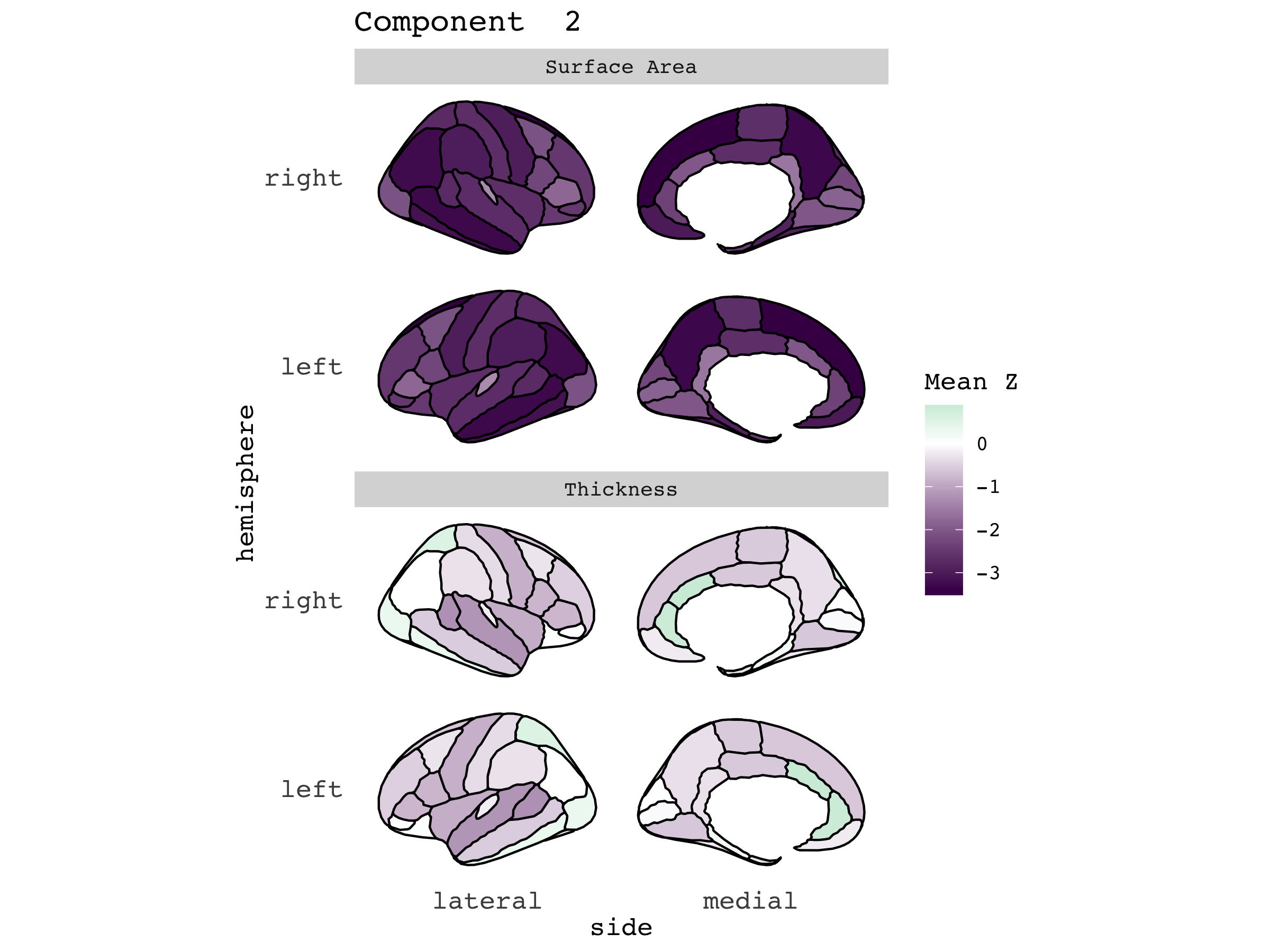

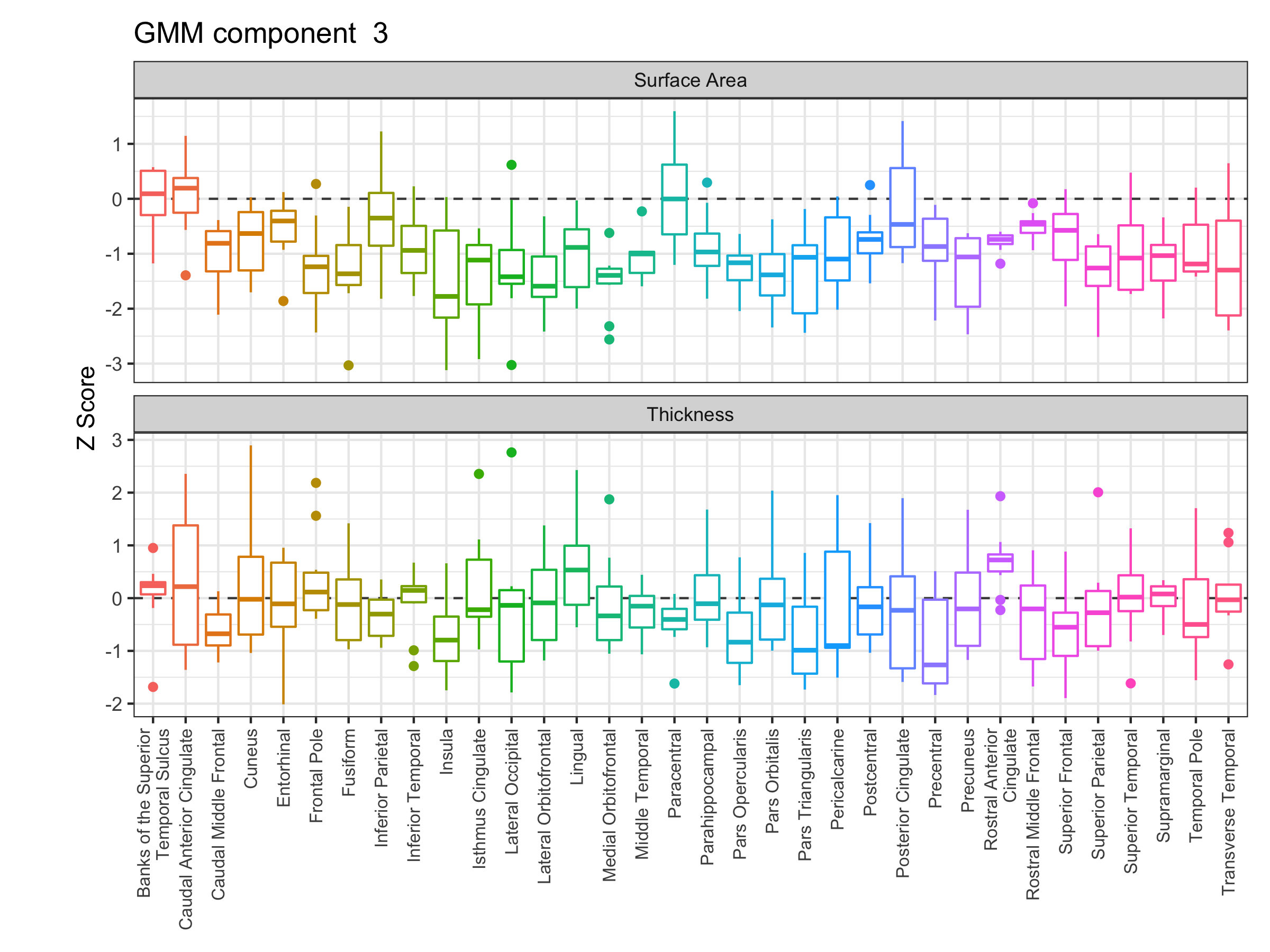

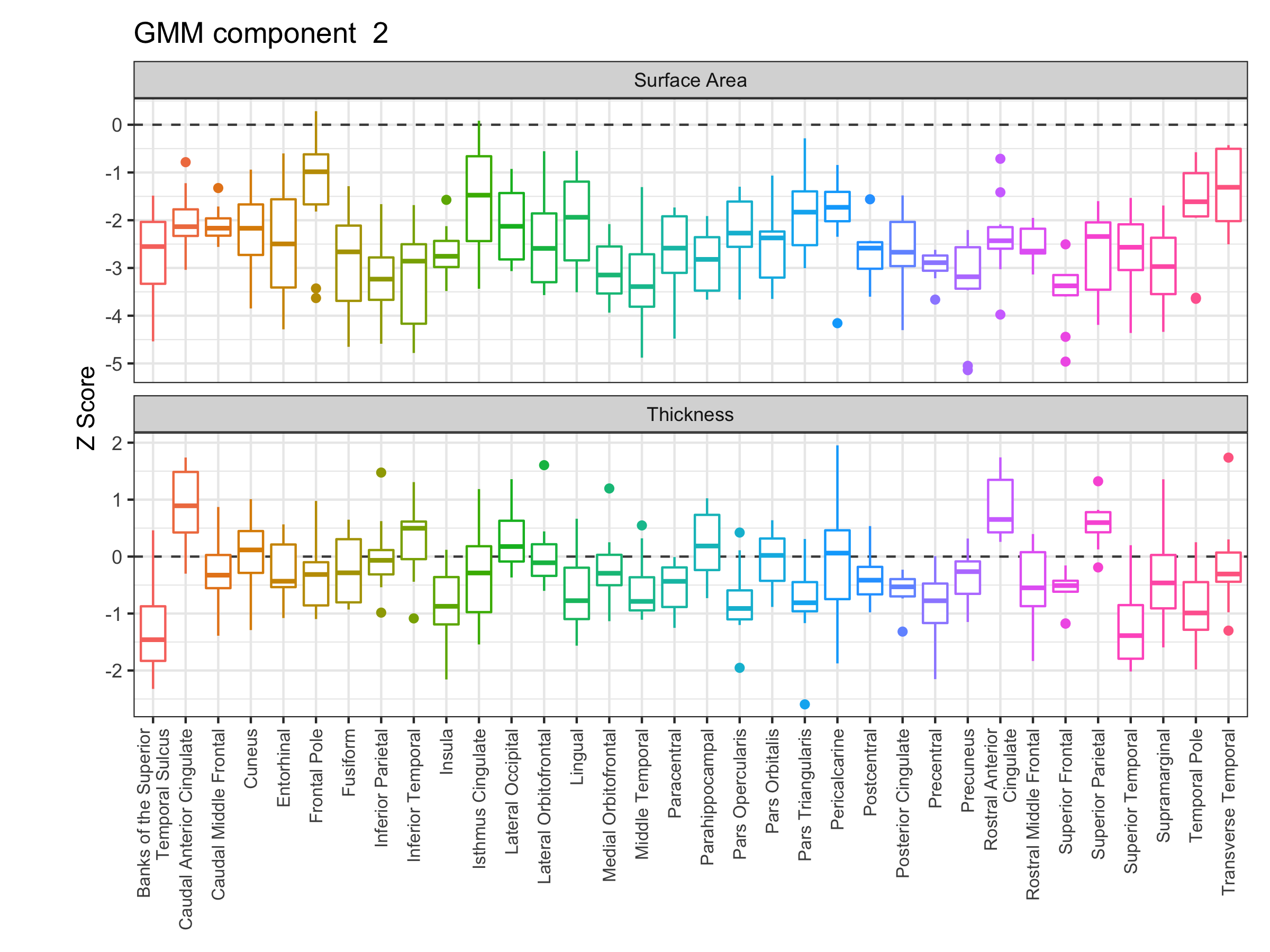

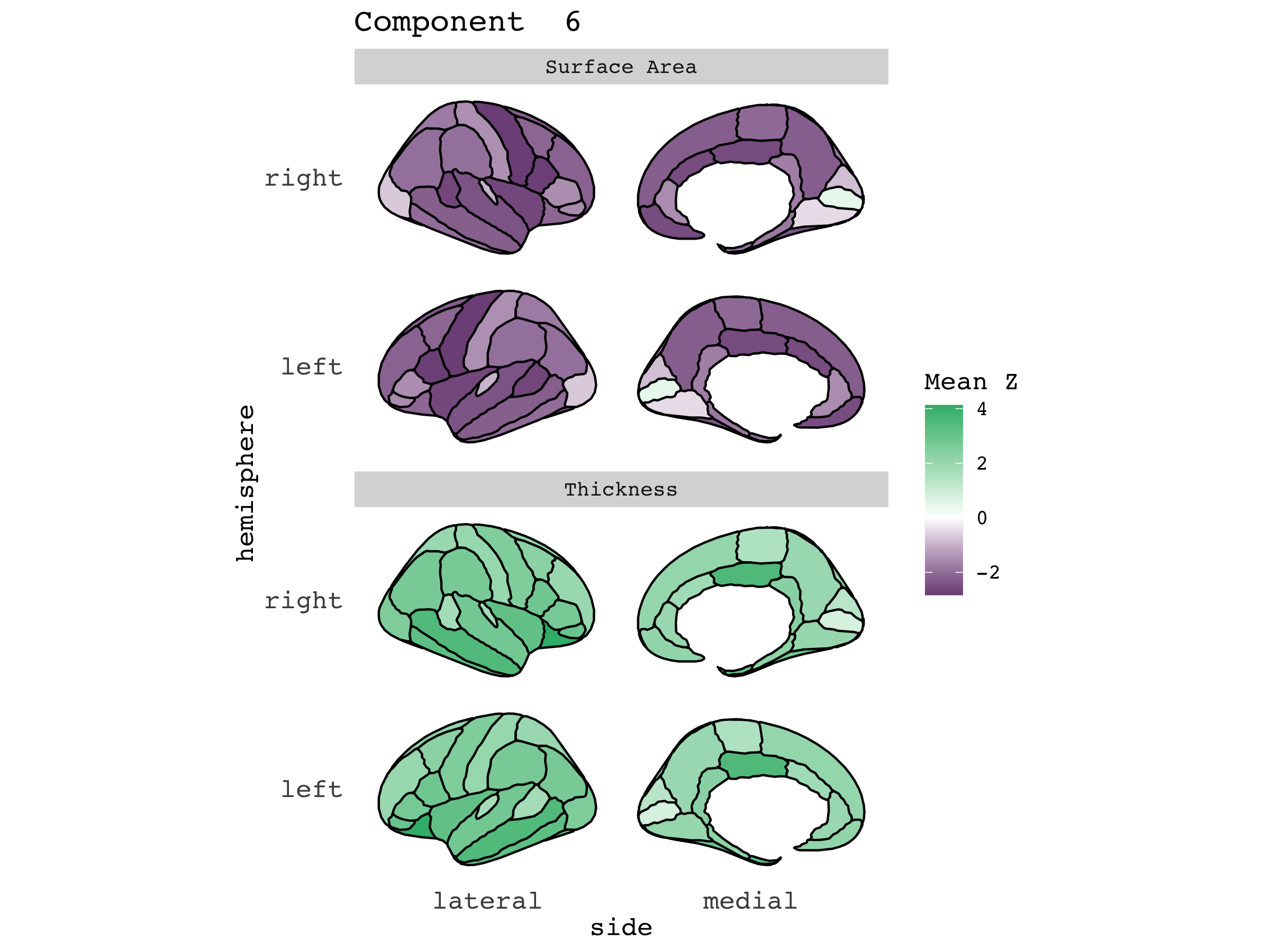

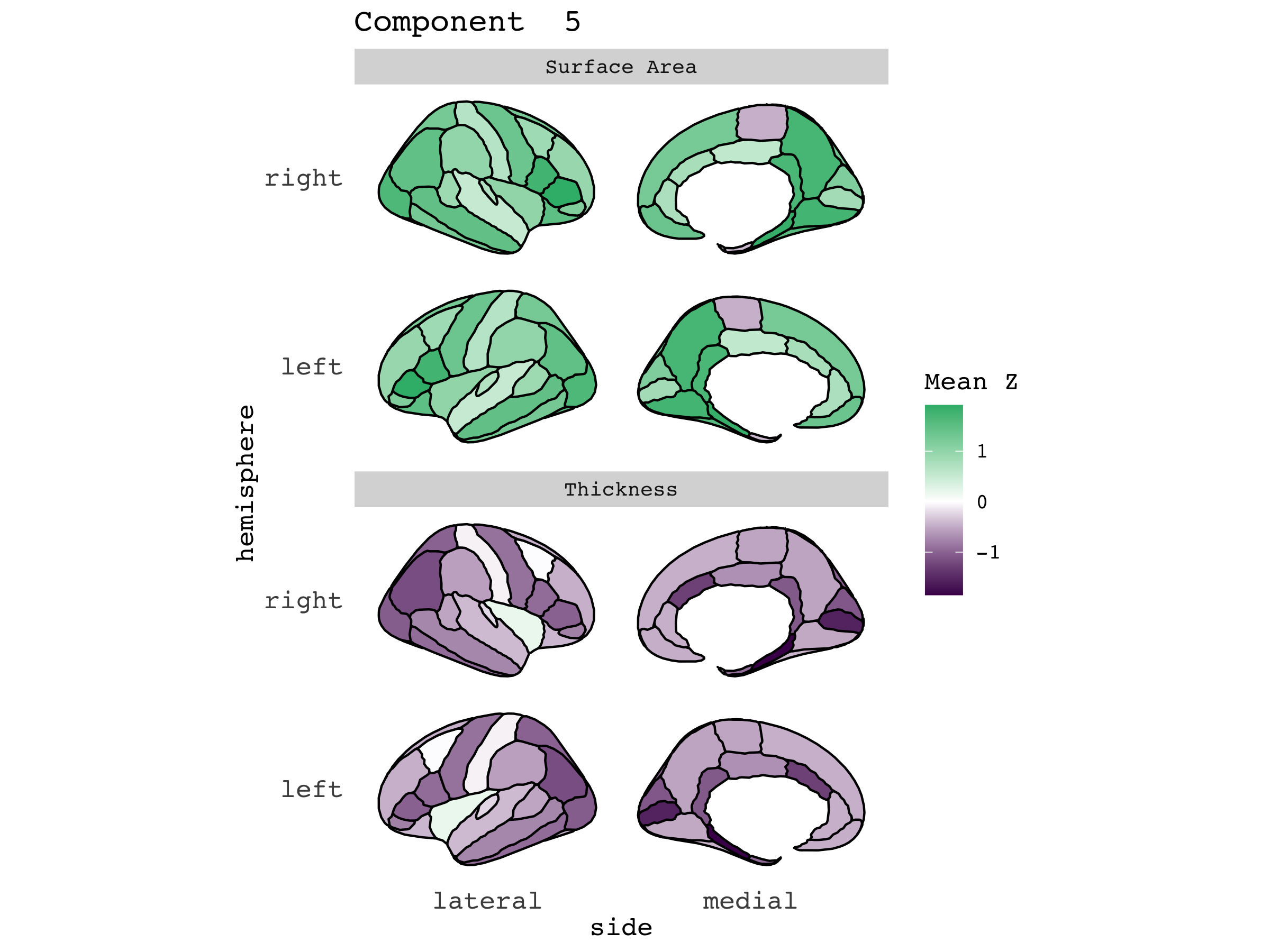

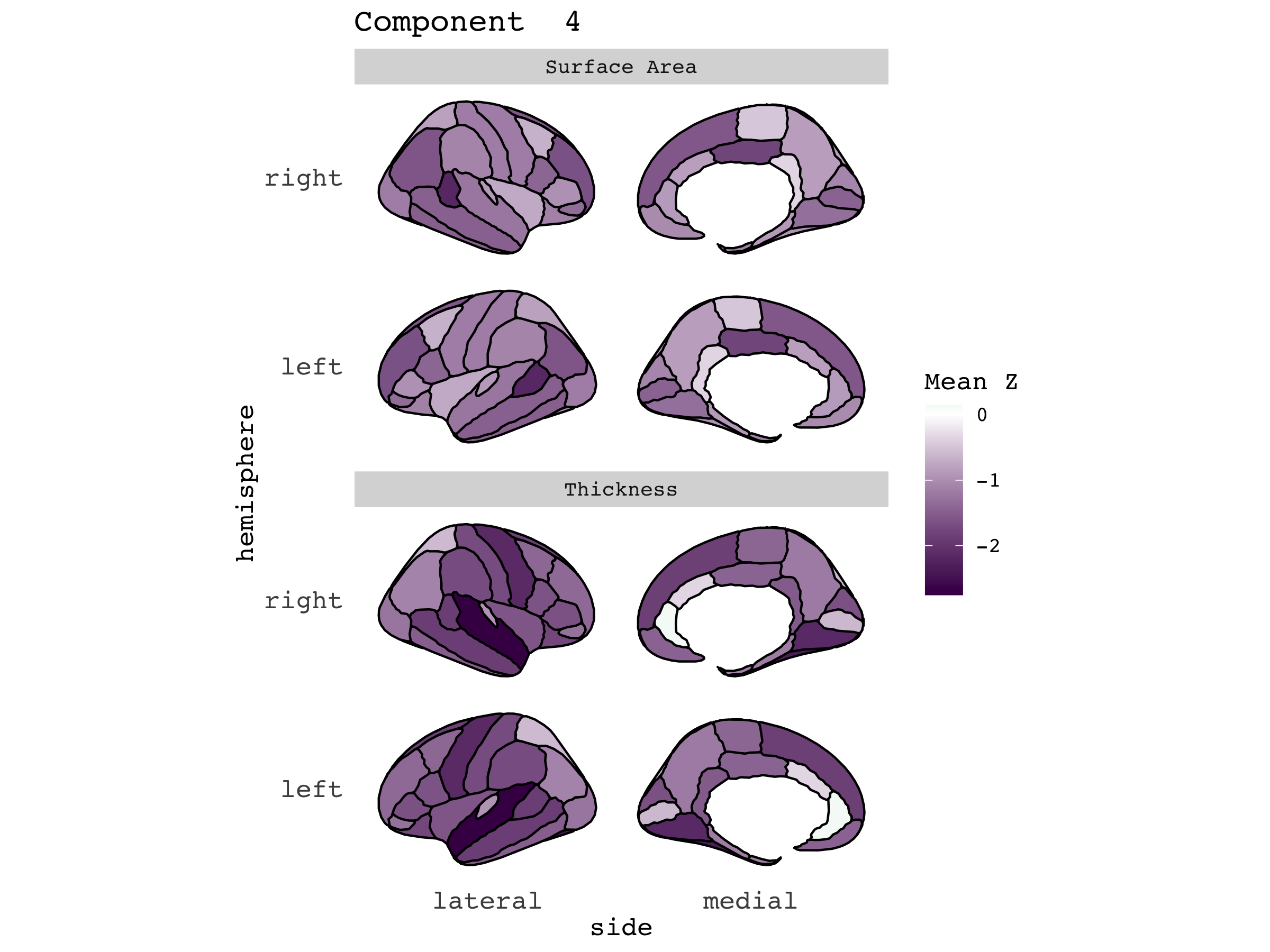

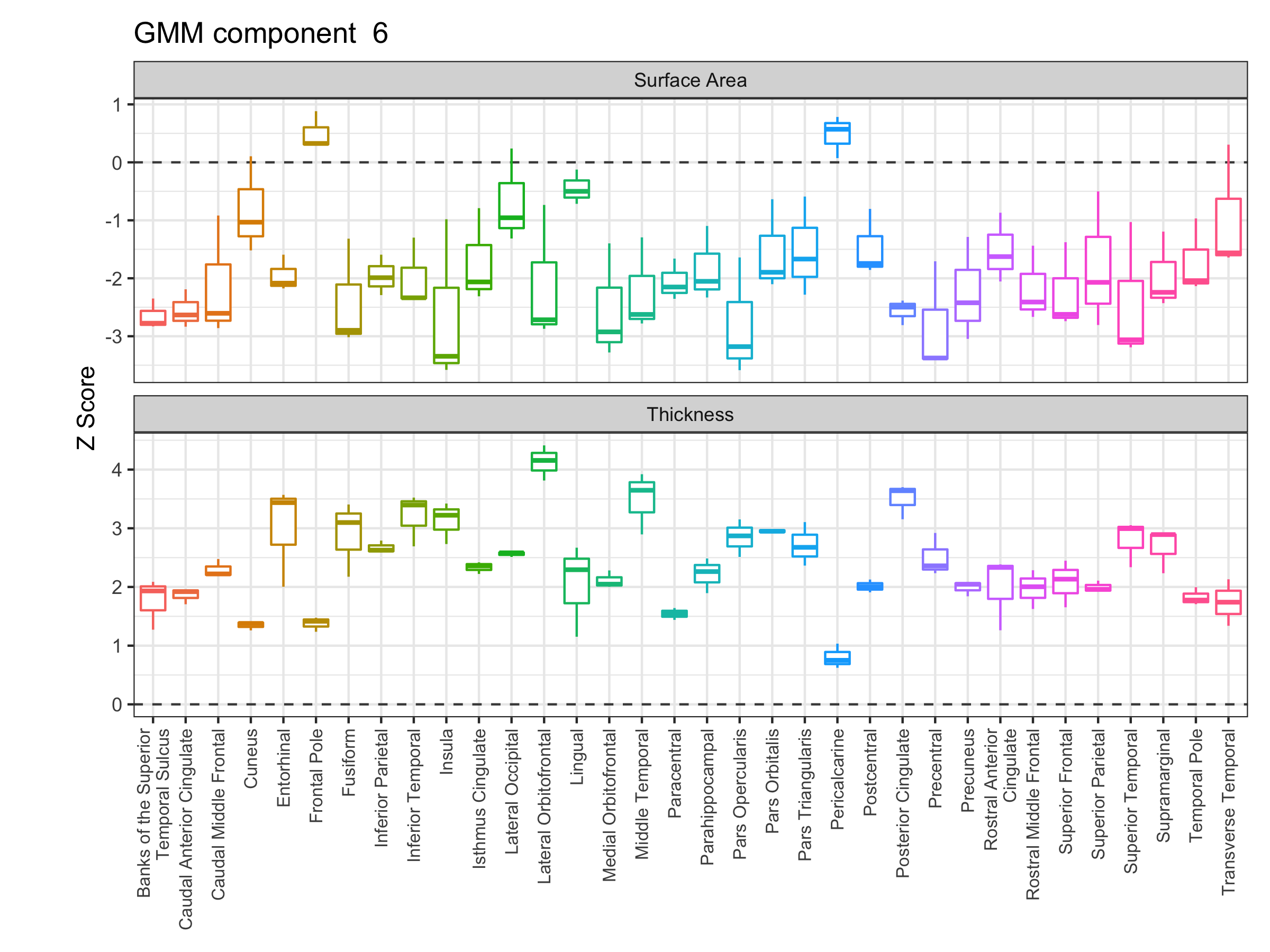

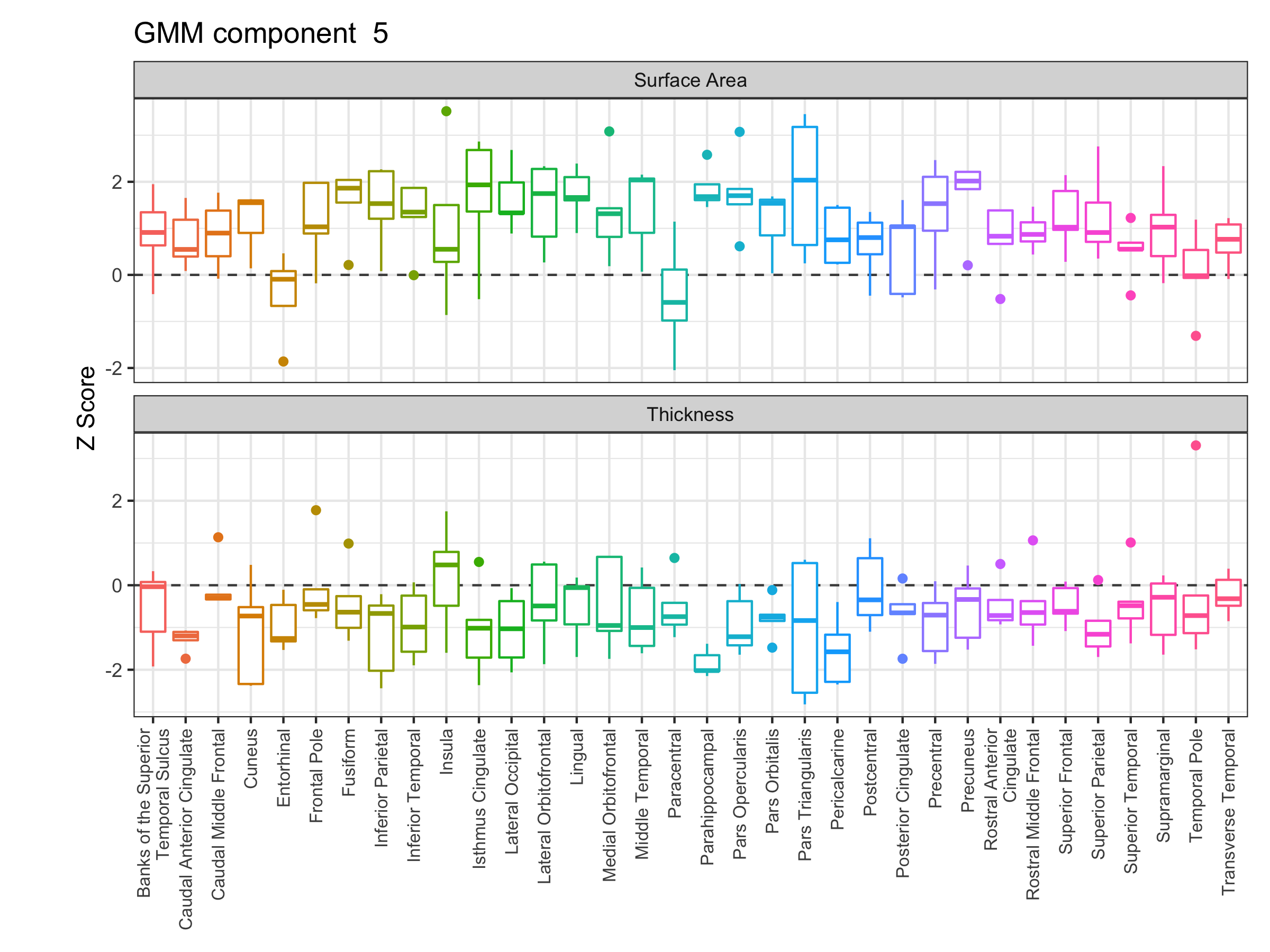

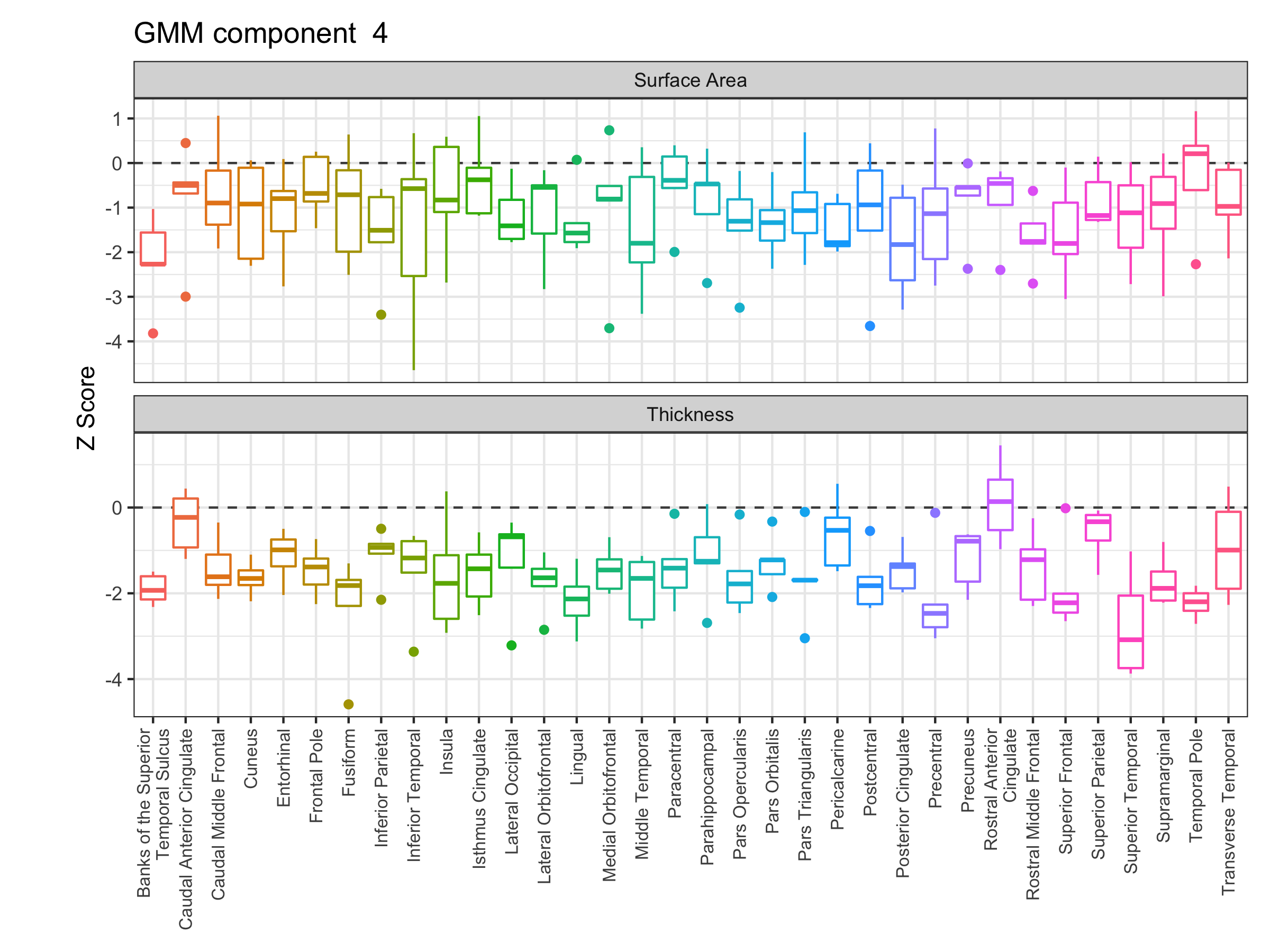

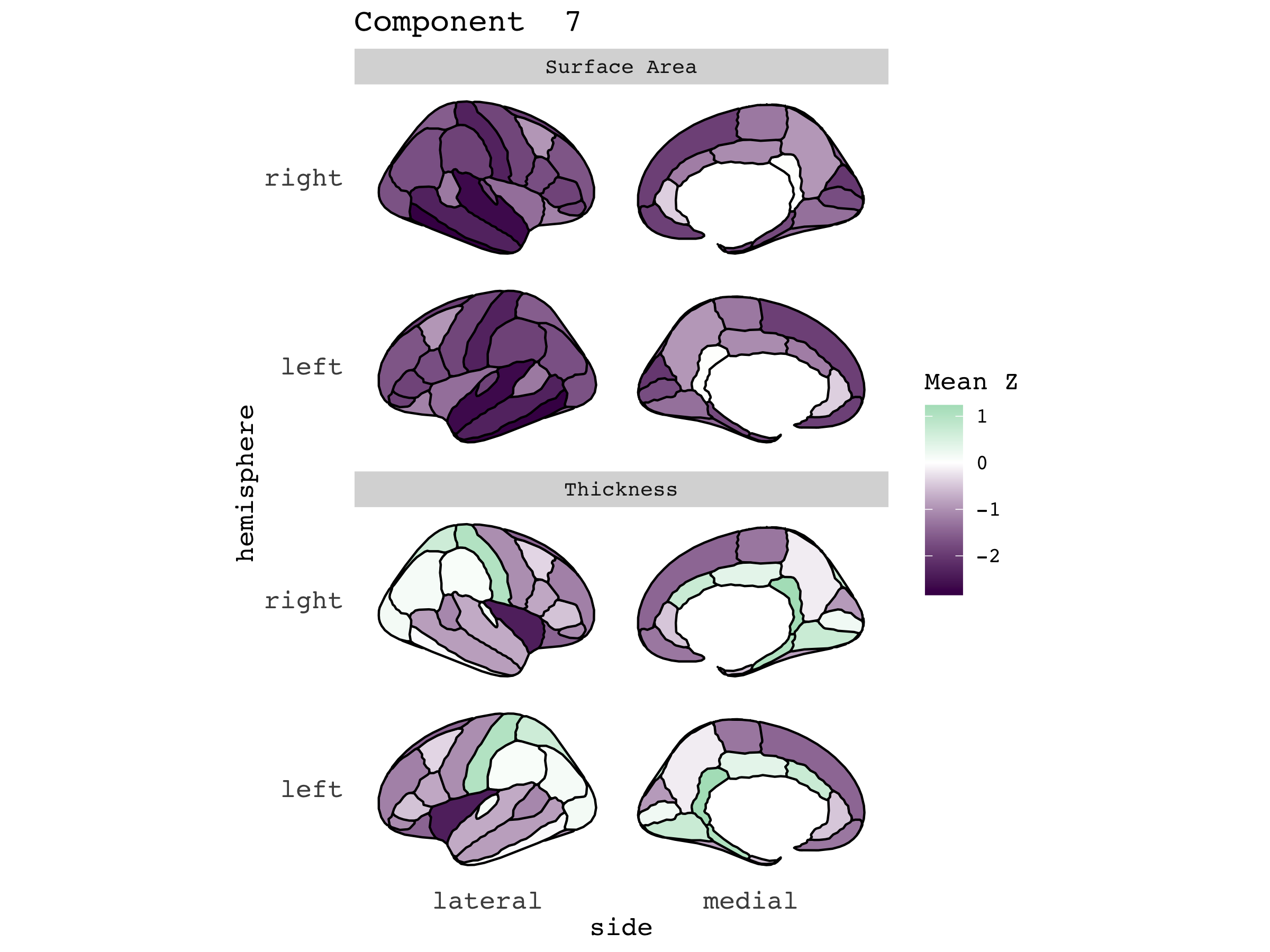

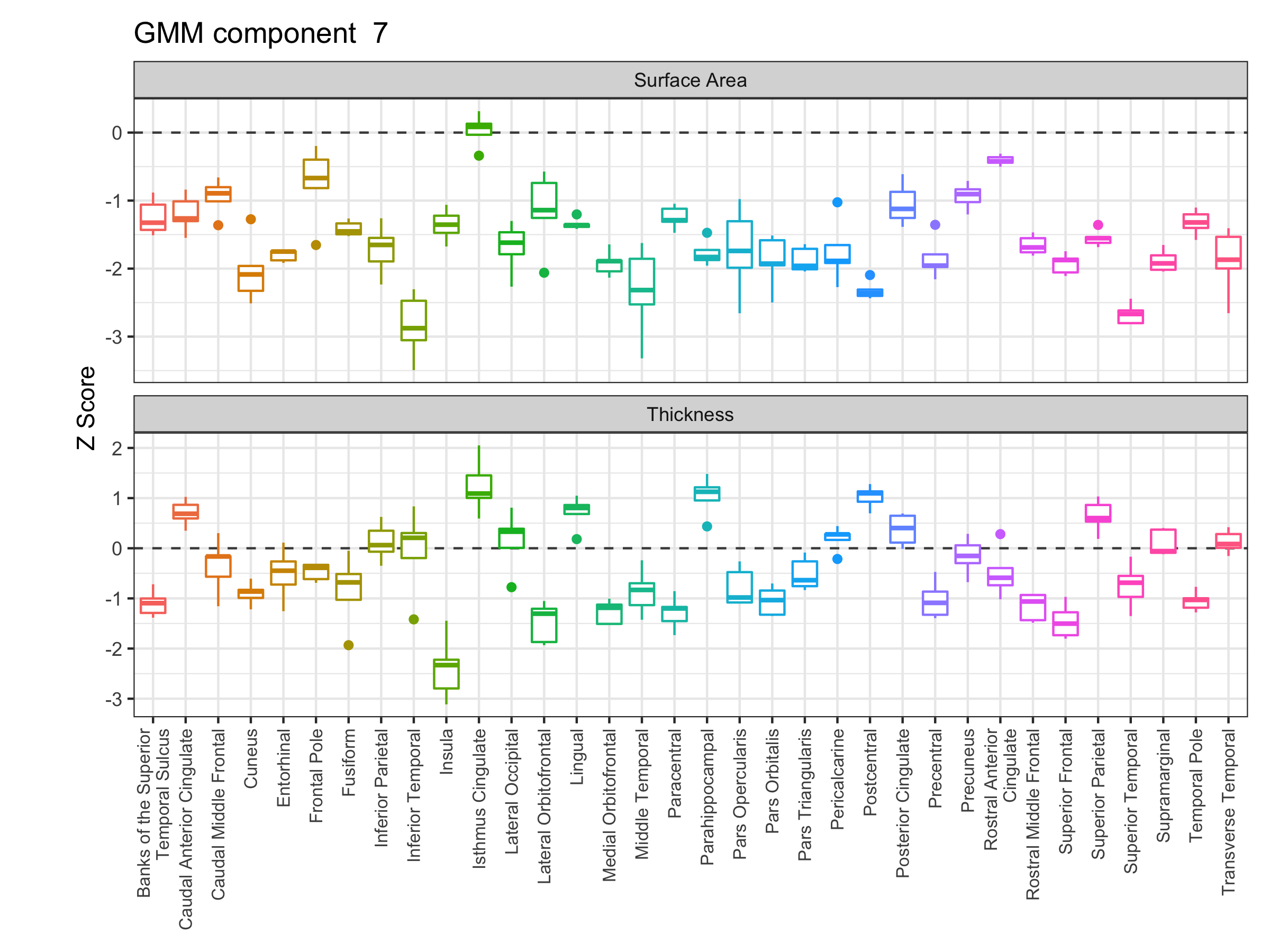

**Figure S2. Cortical *Z-*score distributions across all biochemical trait clusters.** (left) Box plots depicting biochemical LDSR *Z*-score distributions for each cortical measure across all seven clusters of biochemical traits identified via finite Gaussian mixture modelling. (right) Spatial distribution of mean biochemical *Z-*scores for all cortical measures across the seven biochemical clusters.

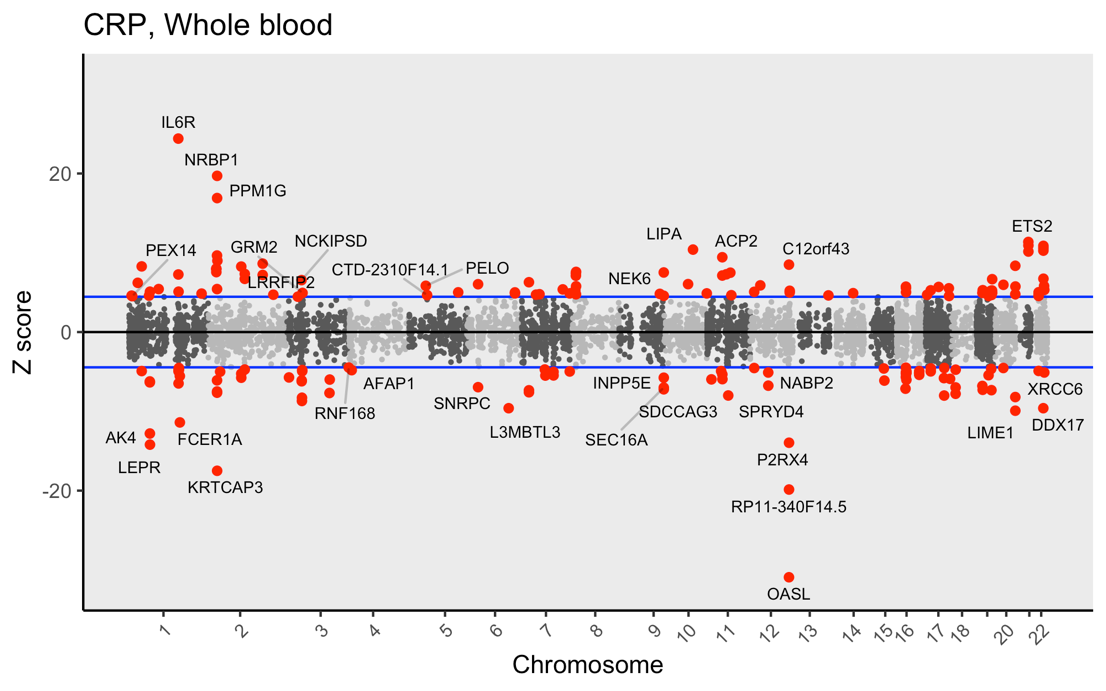

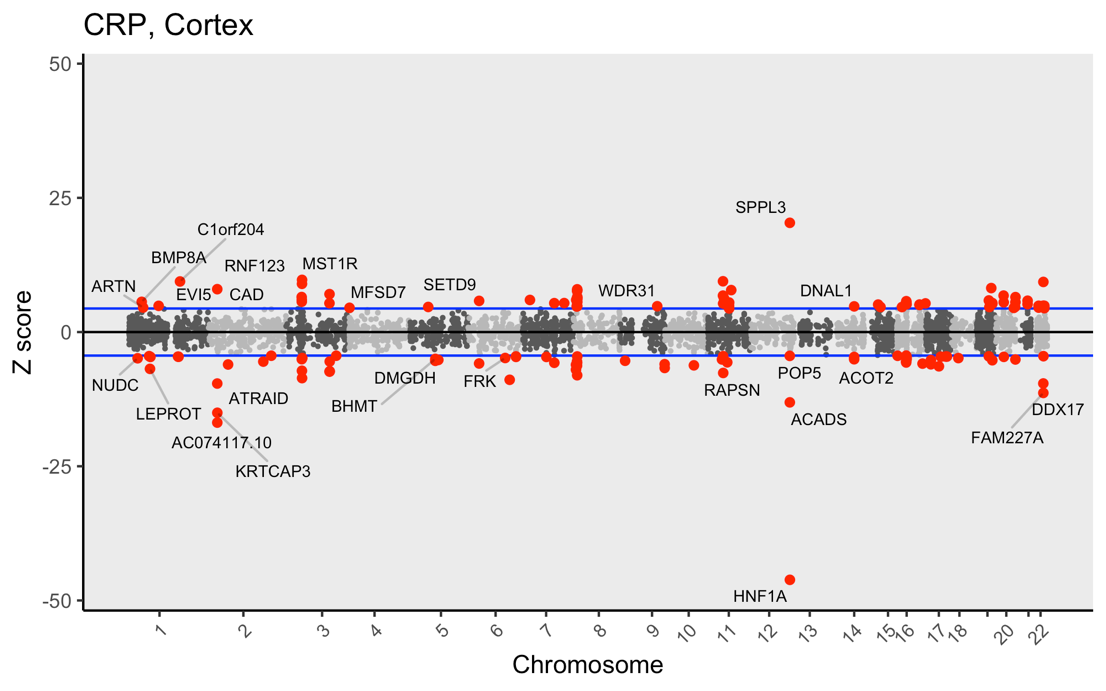

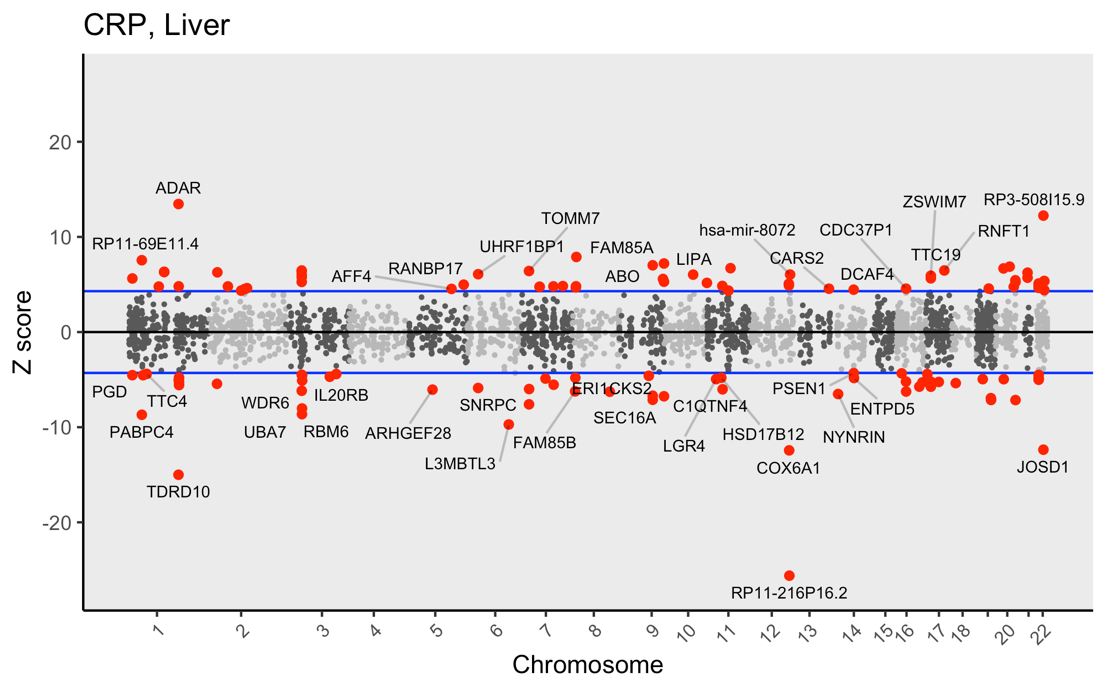

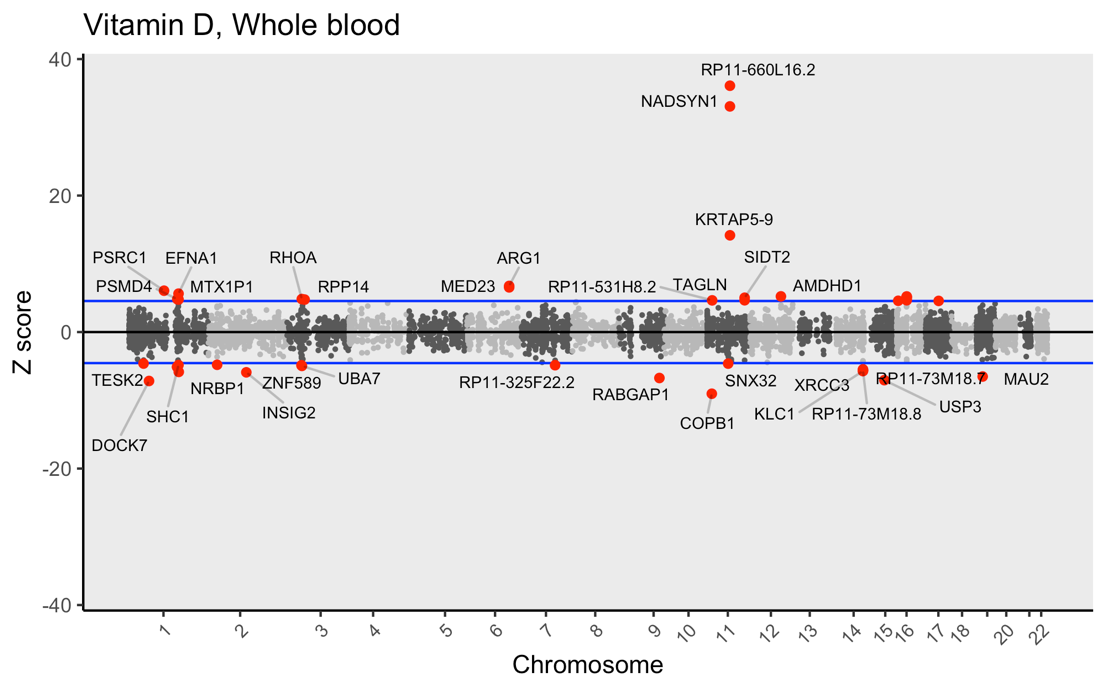

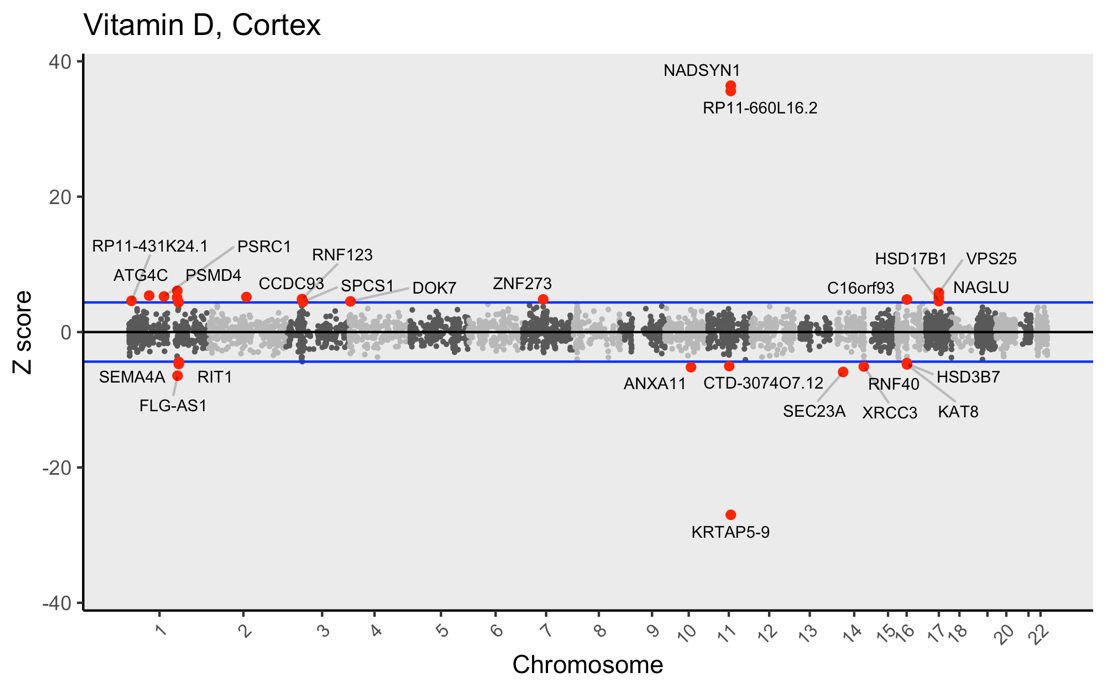

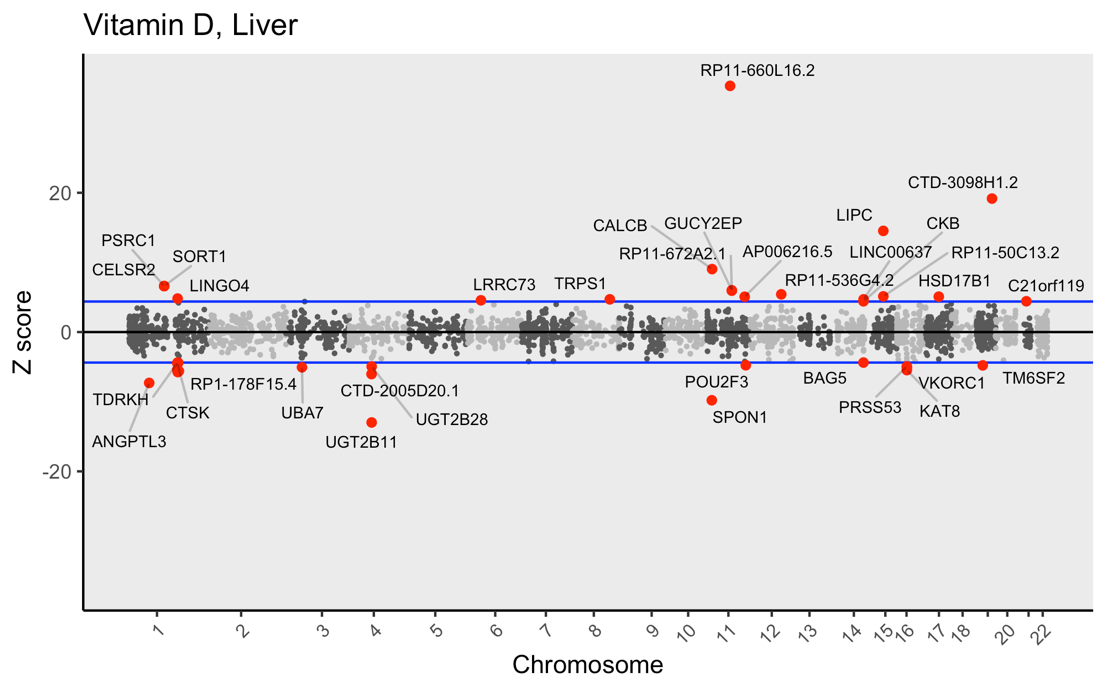

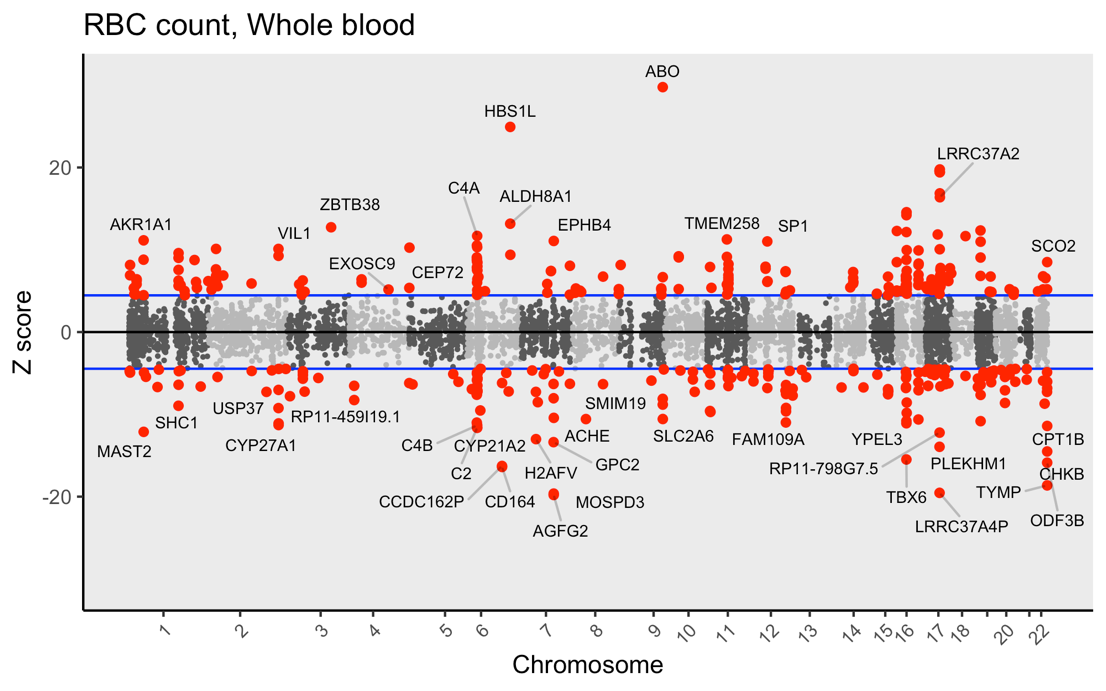

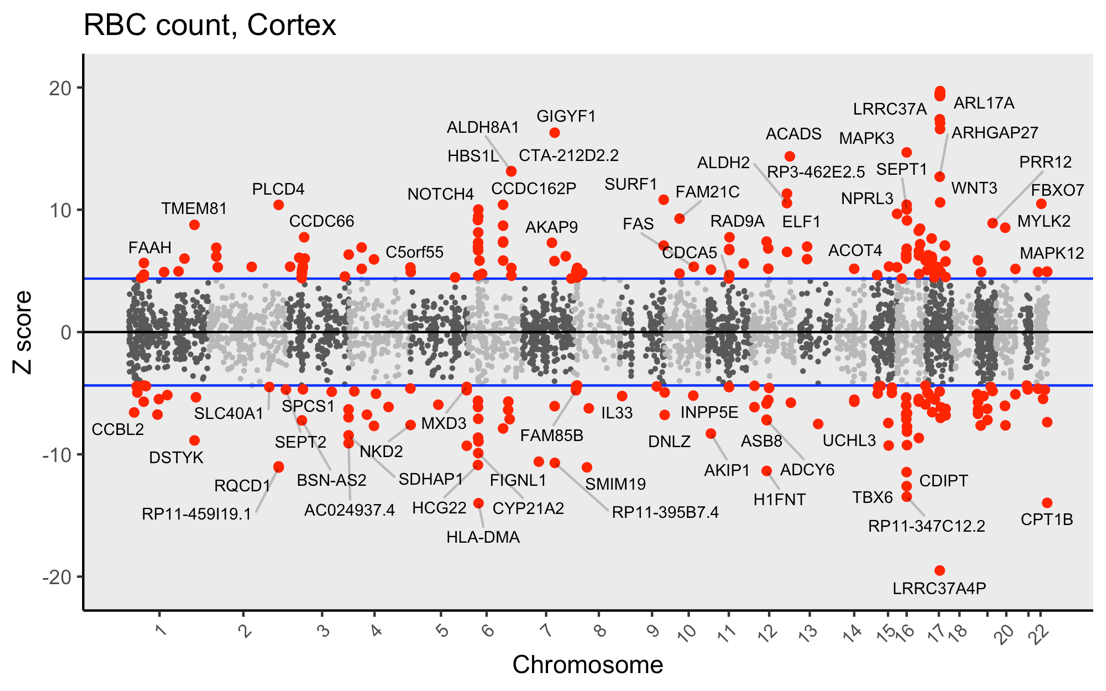

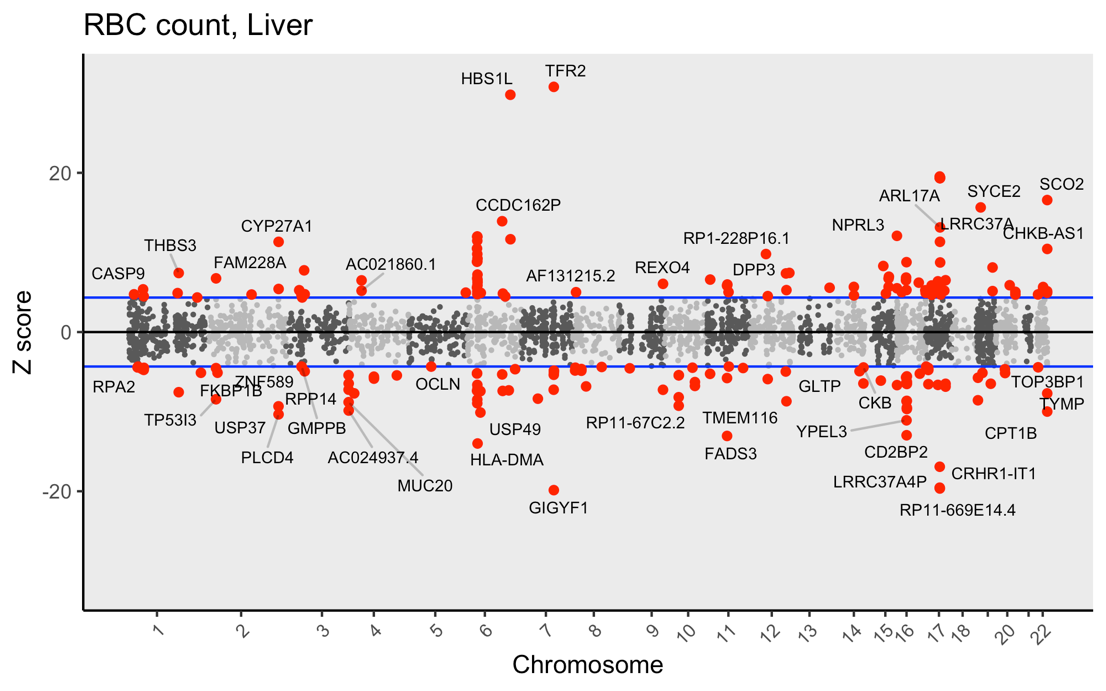

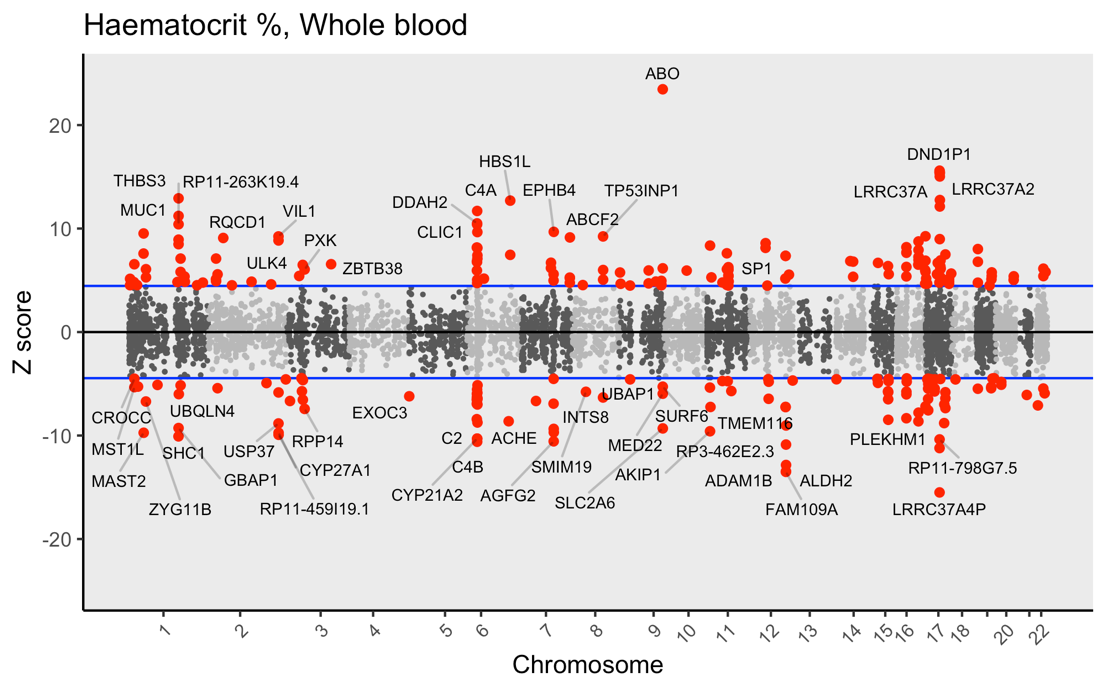

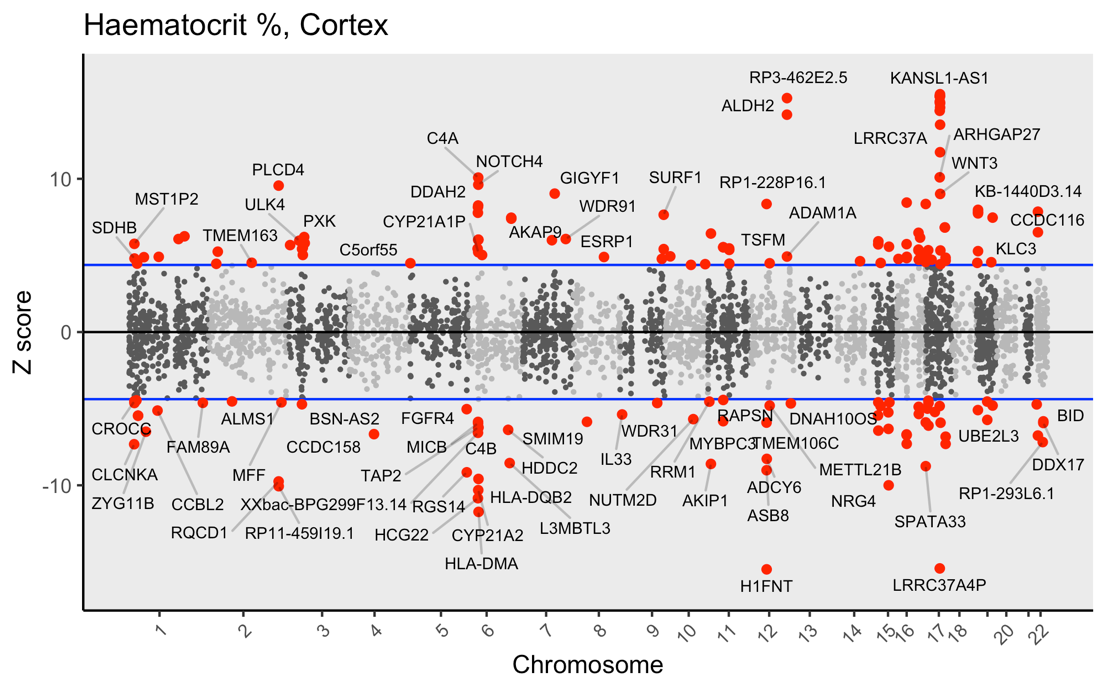

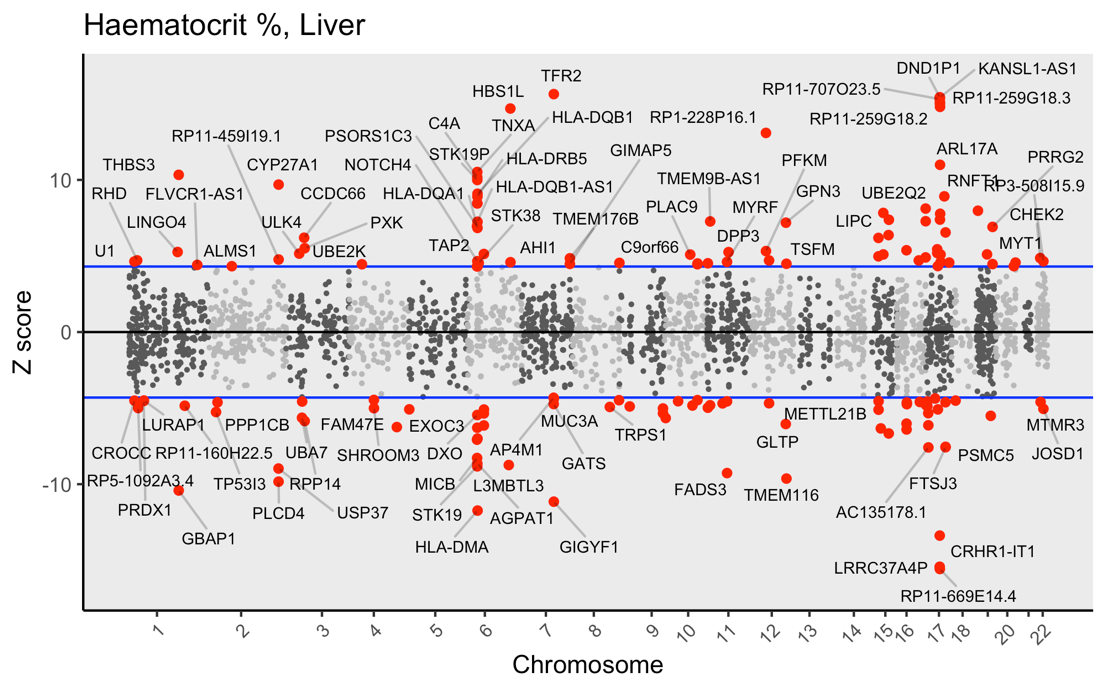

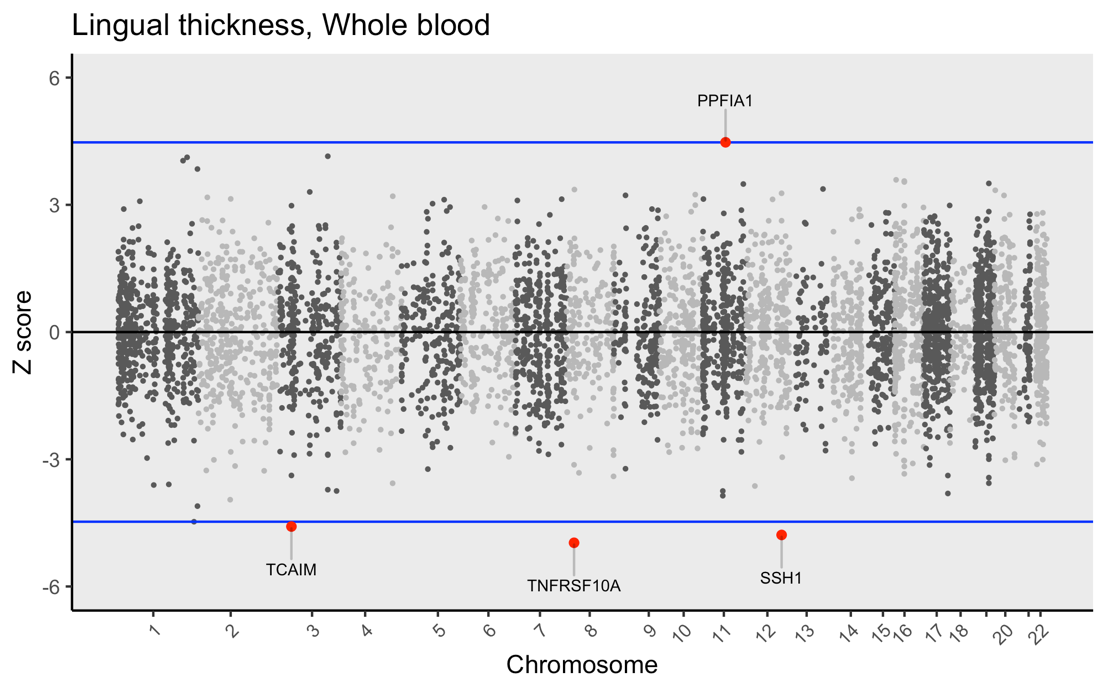

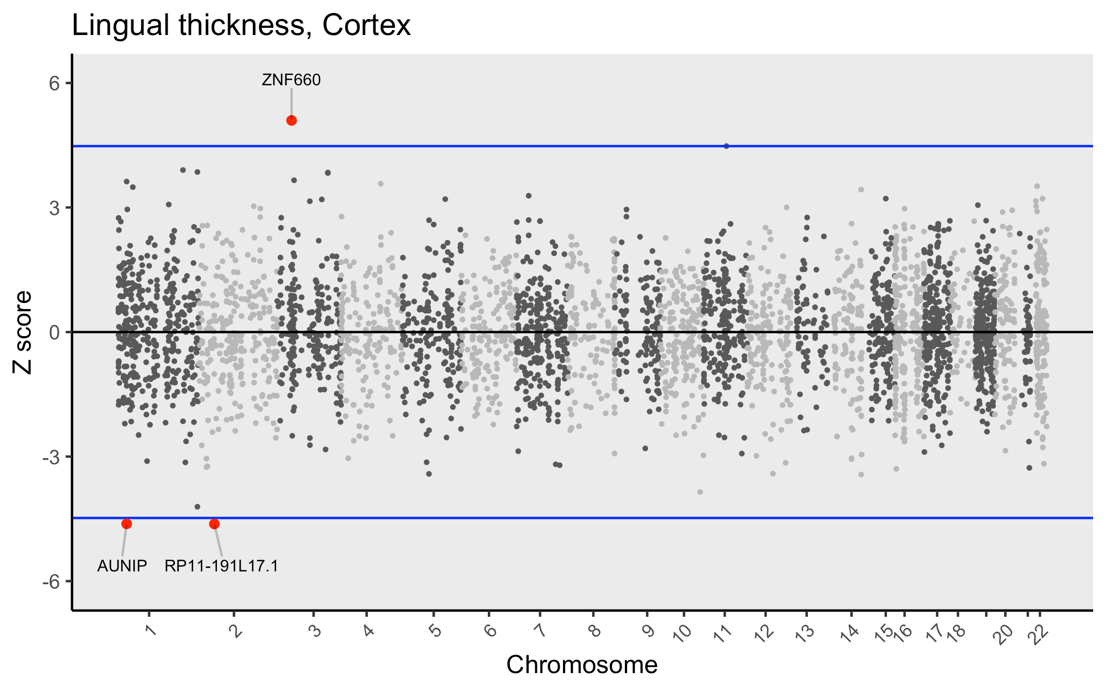

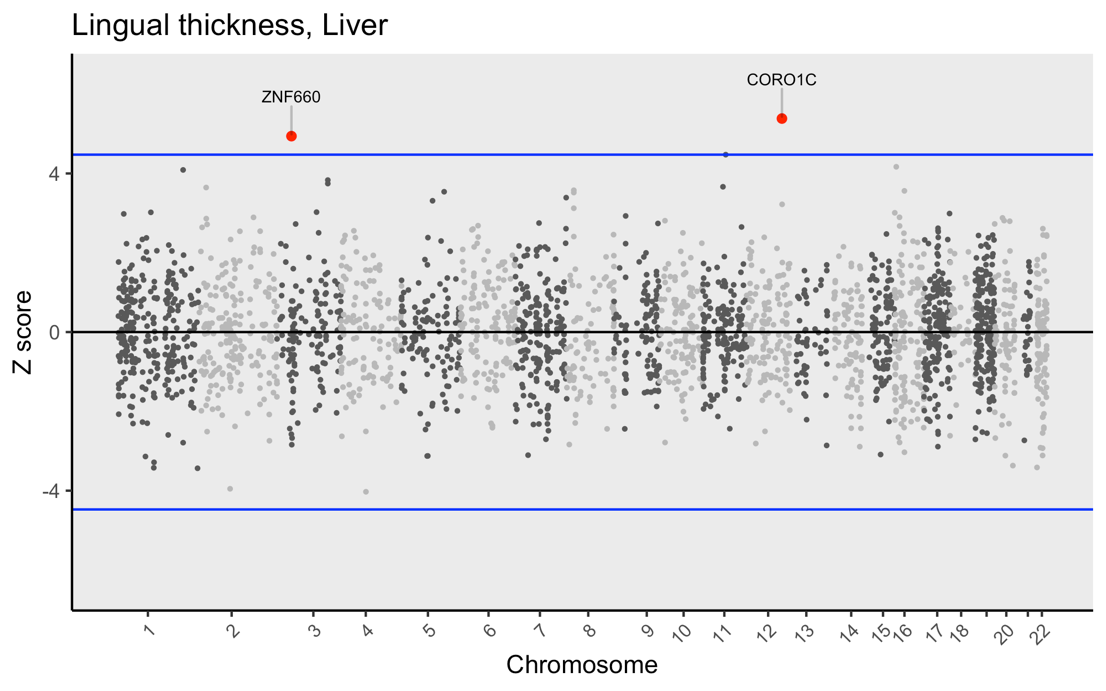

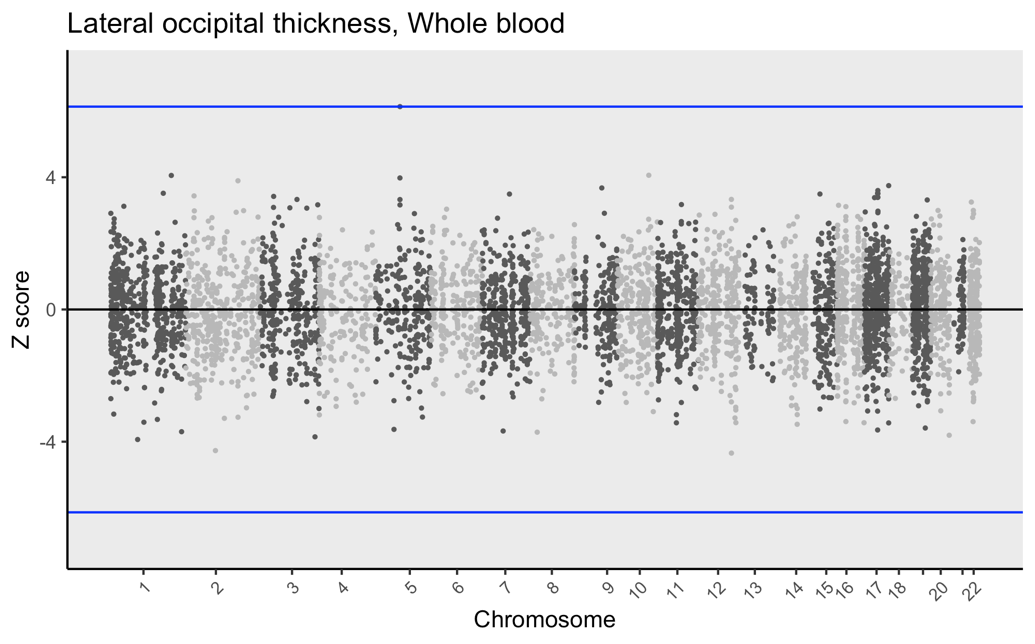

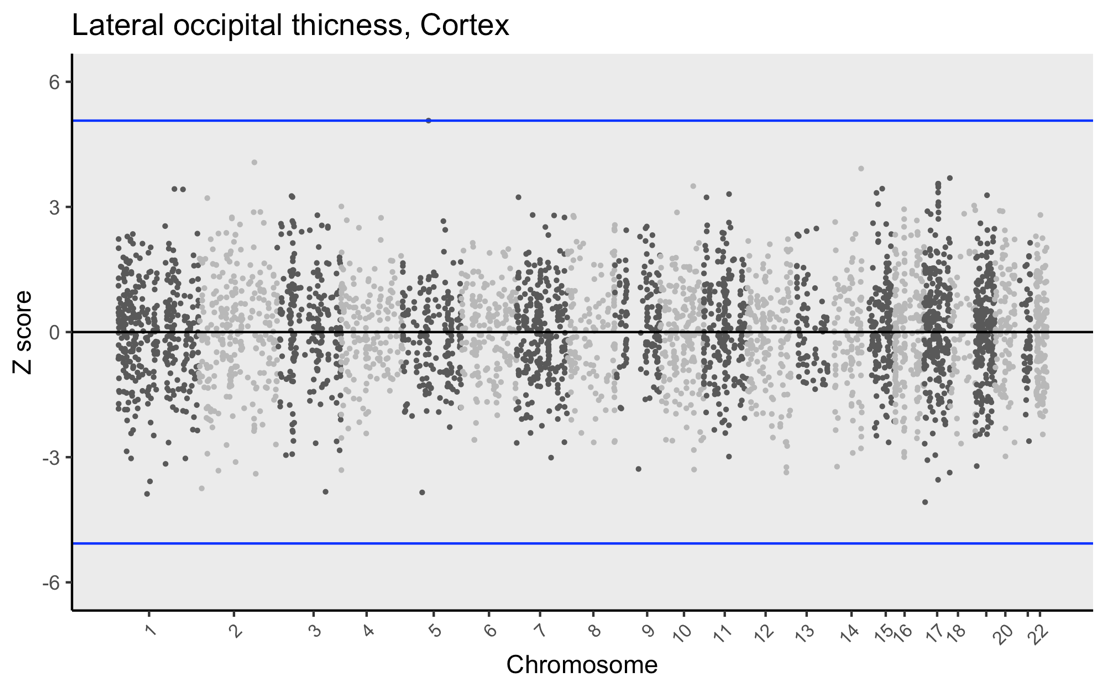

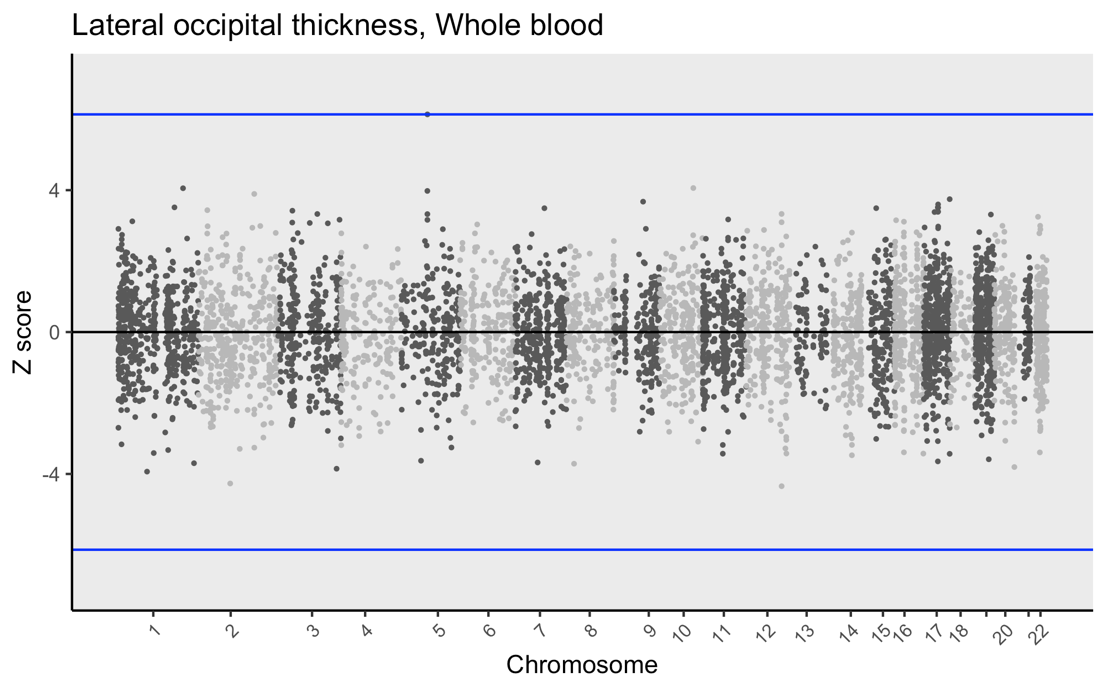

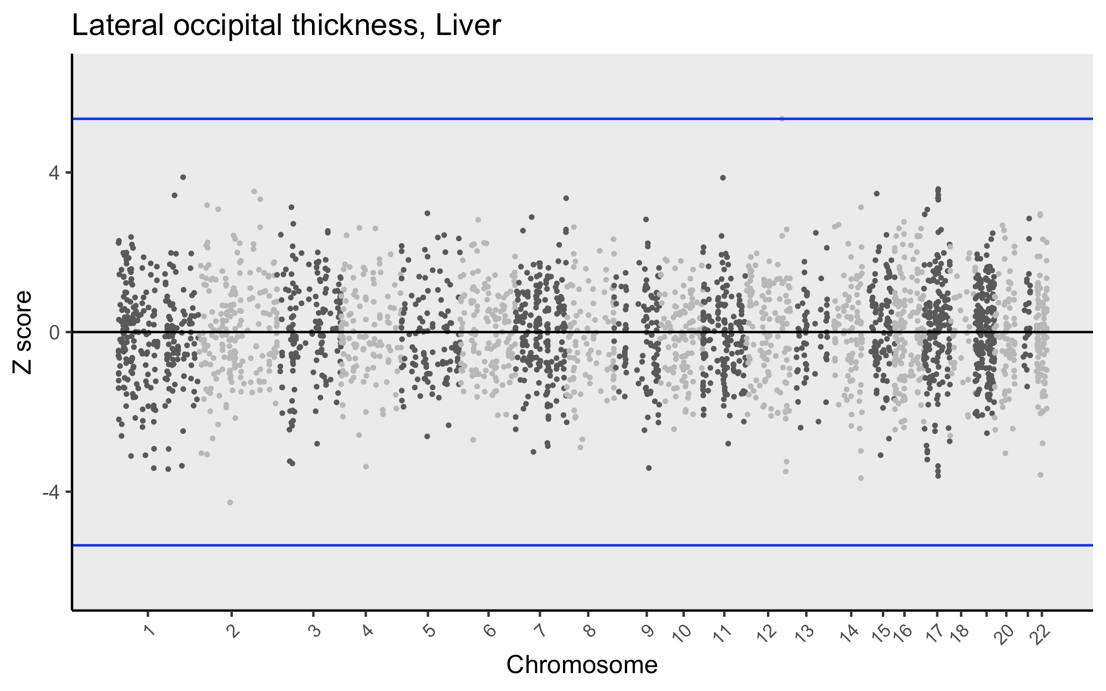

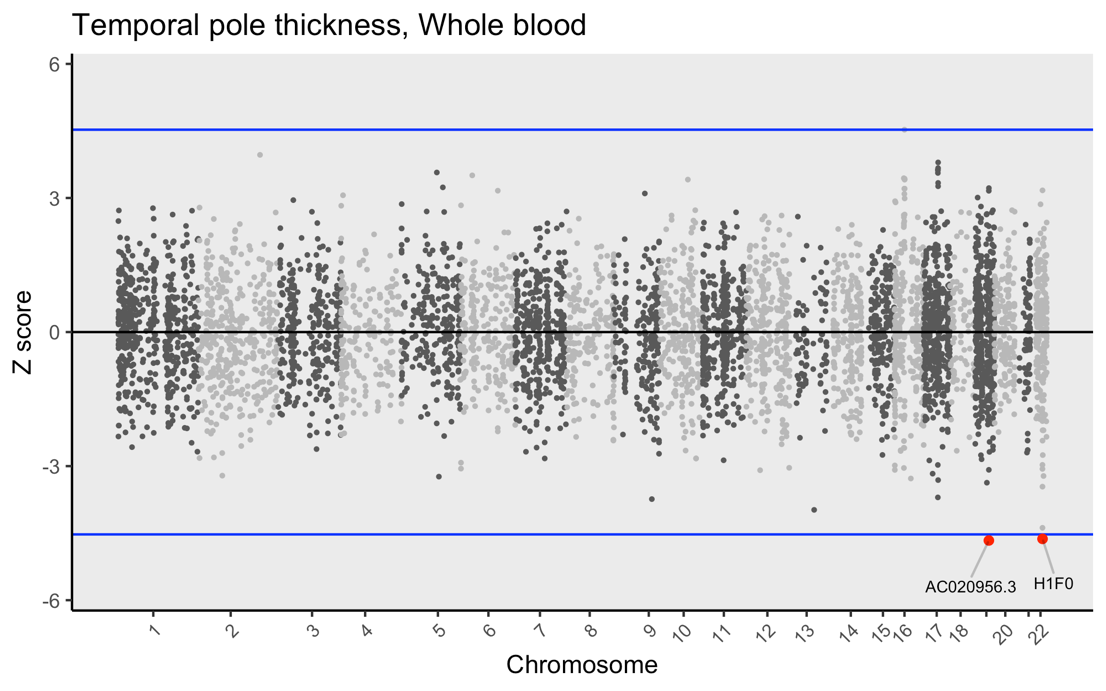

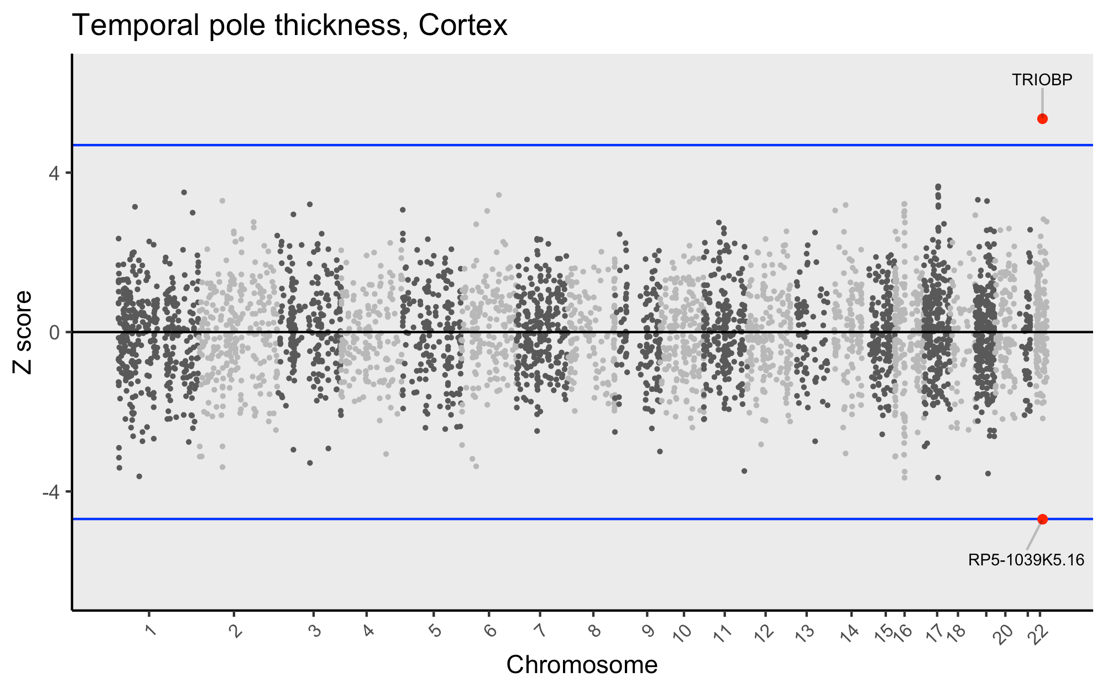

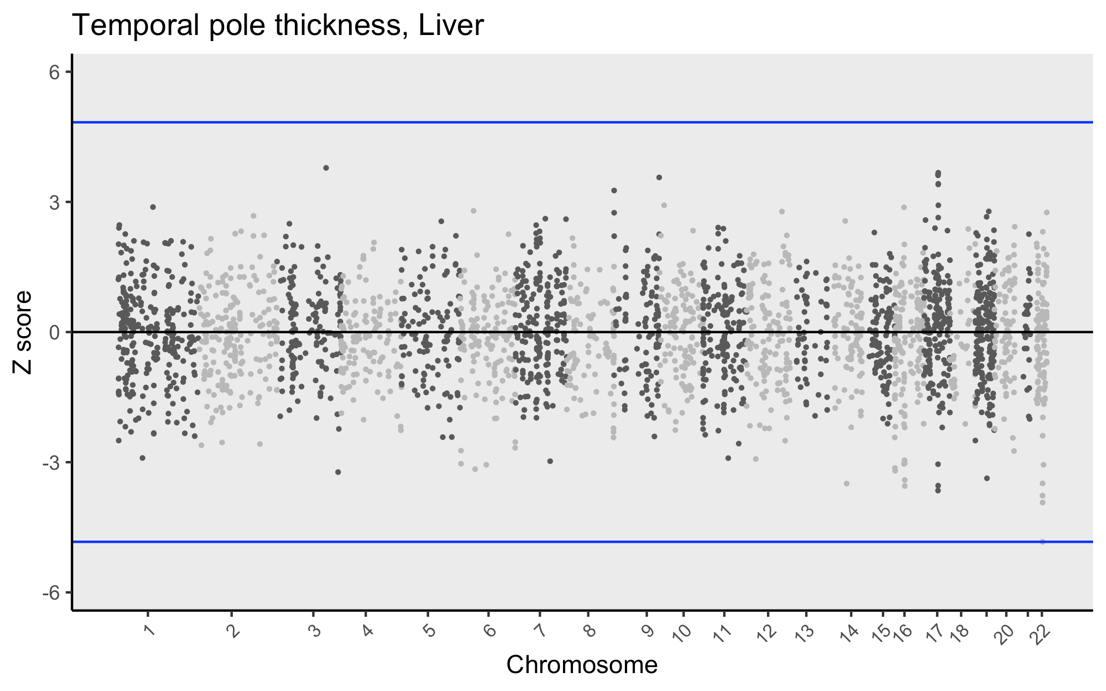

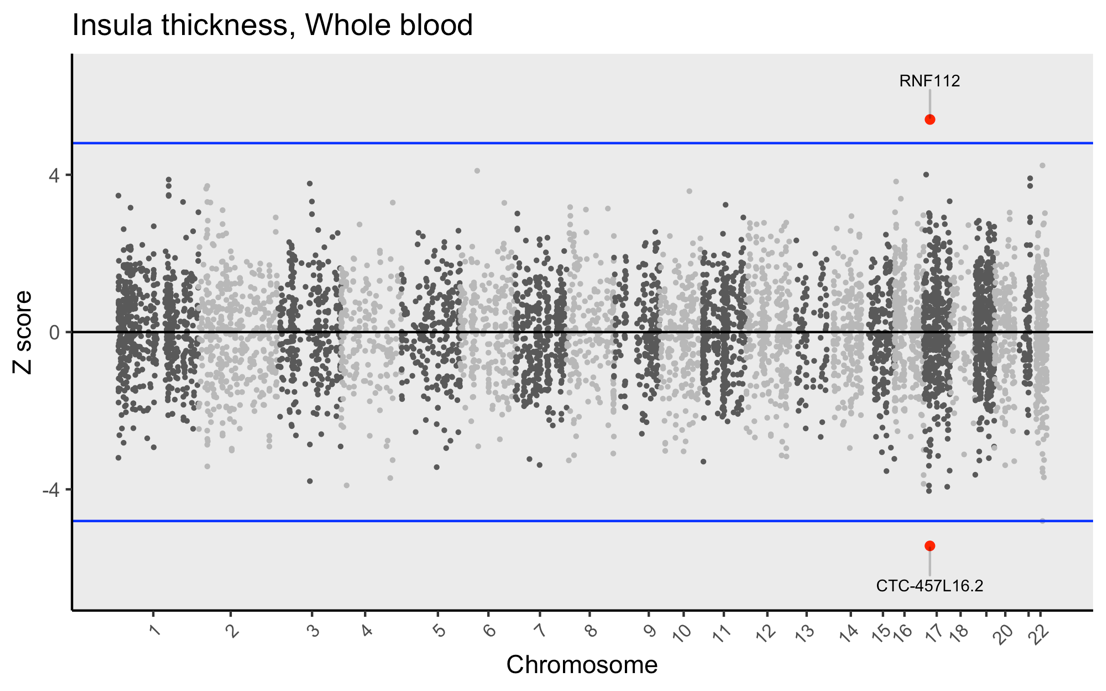

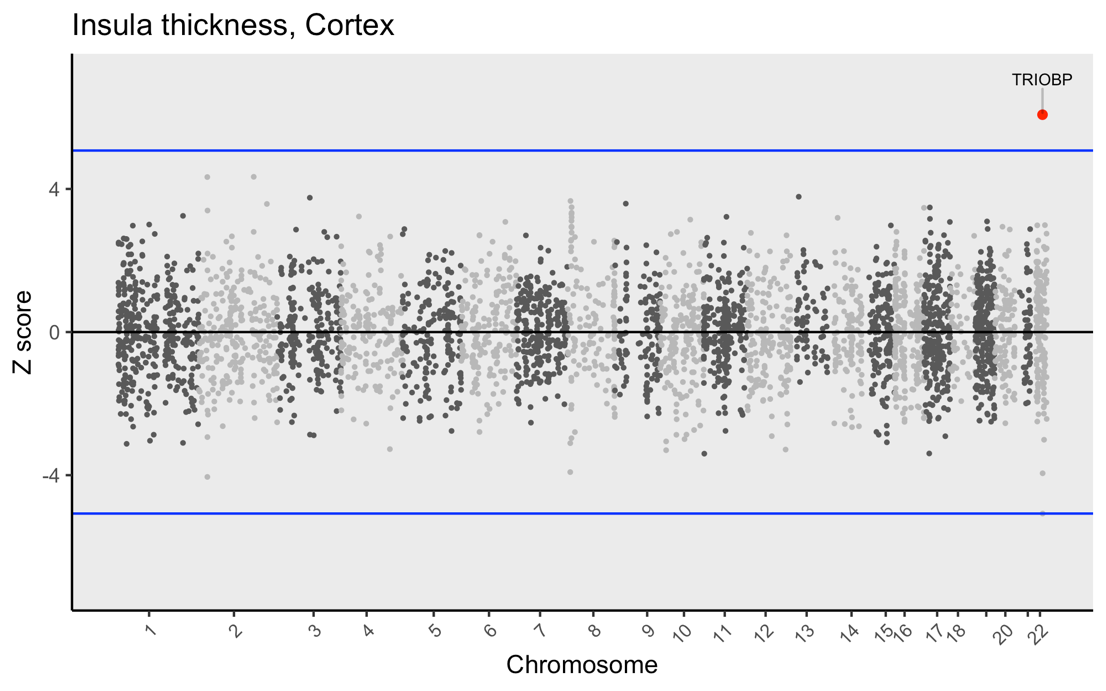

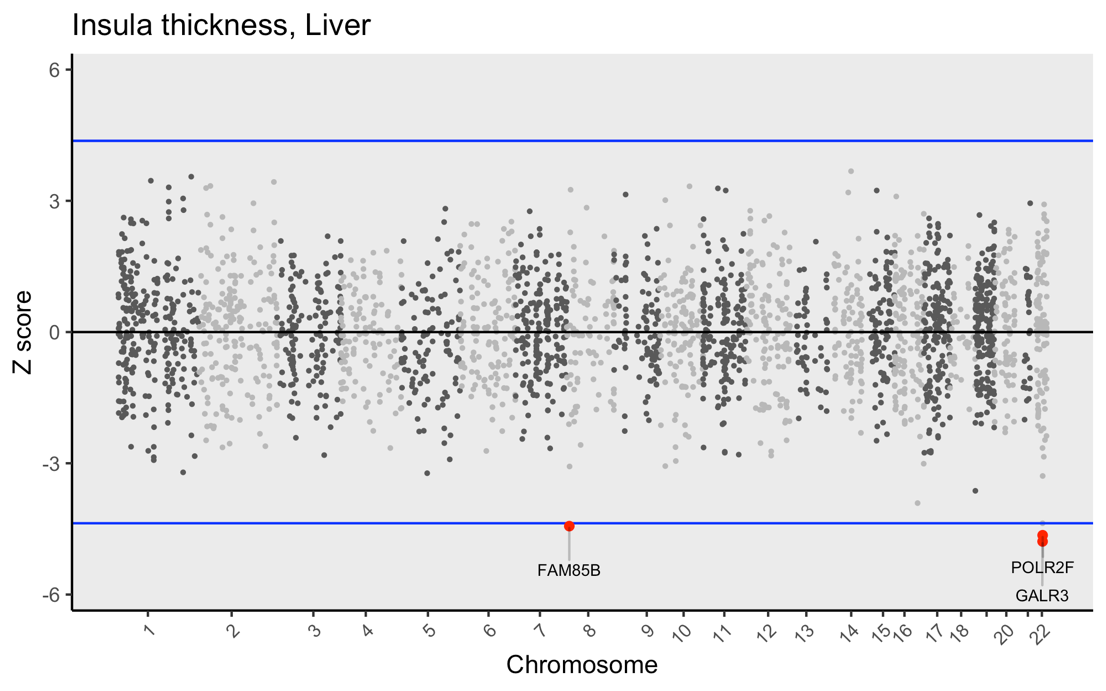

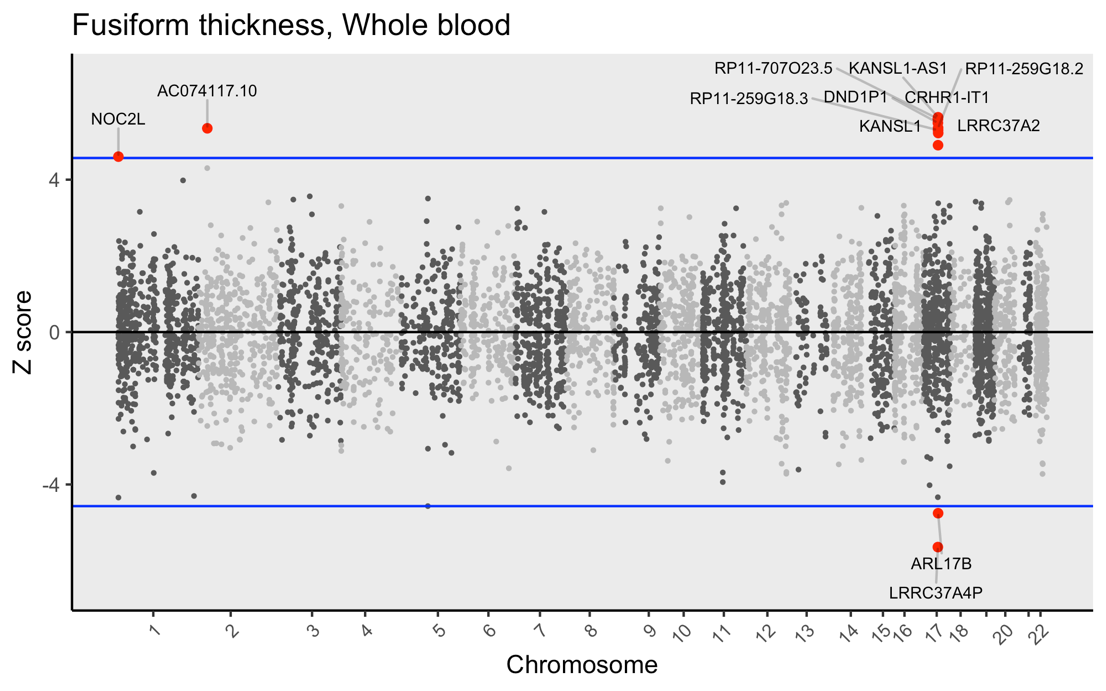

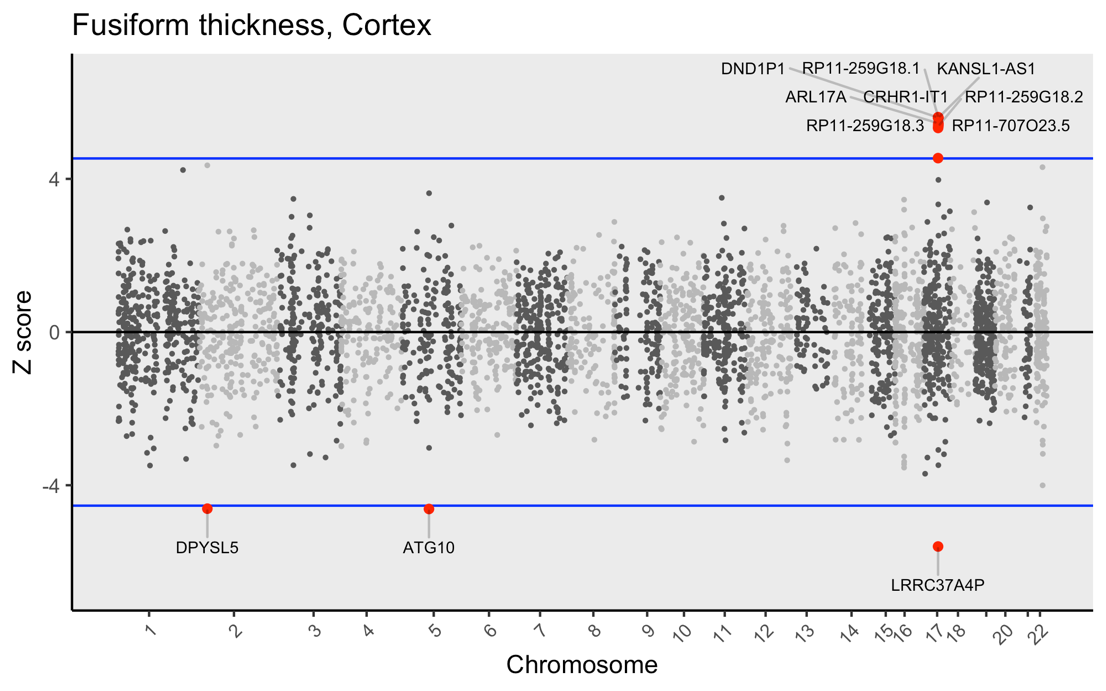

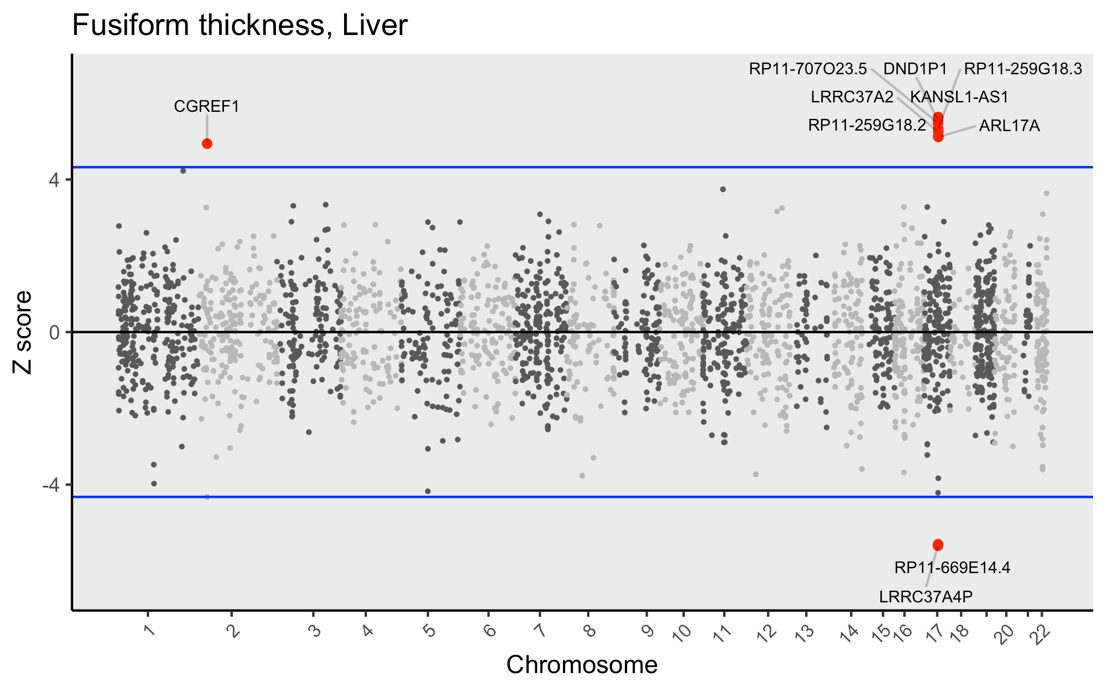

**Figure S3. Miami plots of TWAS results.** Transcriptome-wide imputation was conducted for biochemical and cortical trait GWAS which exhibited evidence for a causal relationship via latent causal variable and Mendelian randomization analyses (i.e. CRP, vitamin D, lingual thickness, lateral occipital thickness and temporal pole thickness). We additionally analysed traits for which *GCP* estimates were attenuated after correction for global cortical thickness (i.e. haematocrit percentage, RBC count, insula thickness and fusiform thickness) to explore transcriptomic similarities amongst these traits. Gene expression was estimated using whole blood, cortical and liver expression weights (GTEx v7), the latter of which was included as CRP is predominantly synthesised in the liver. Note the vast extent of significantly associated genes identified for biochemical traits, in contrast to the cortical traits. Blue lines correspond to *P_Bonferroni_* < 0.05. All cortical traits depicted above were not corrected for global thickness.

**a**

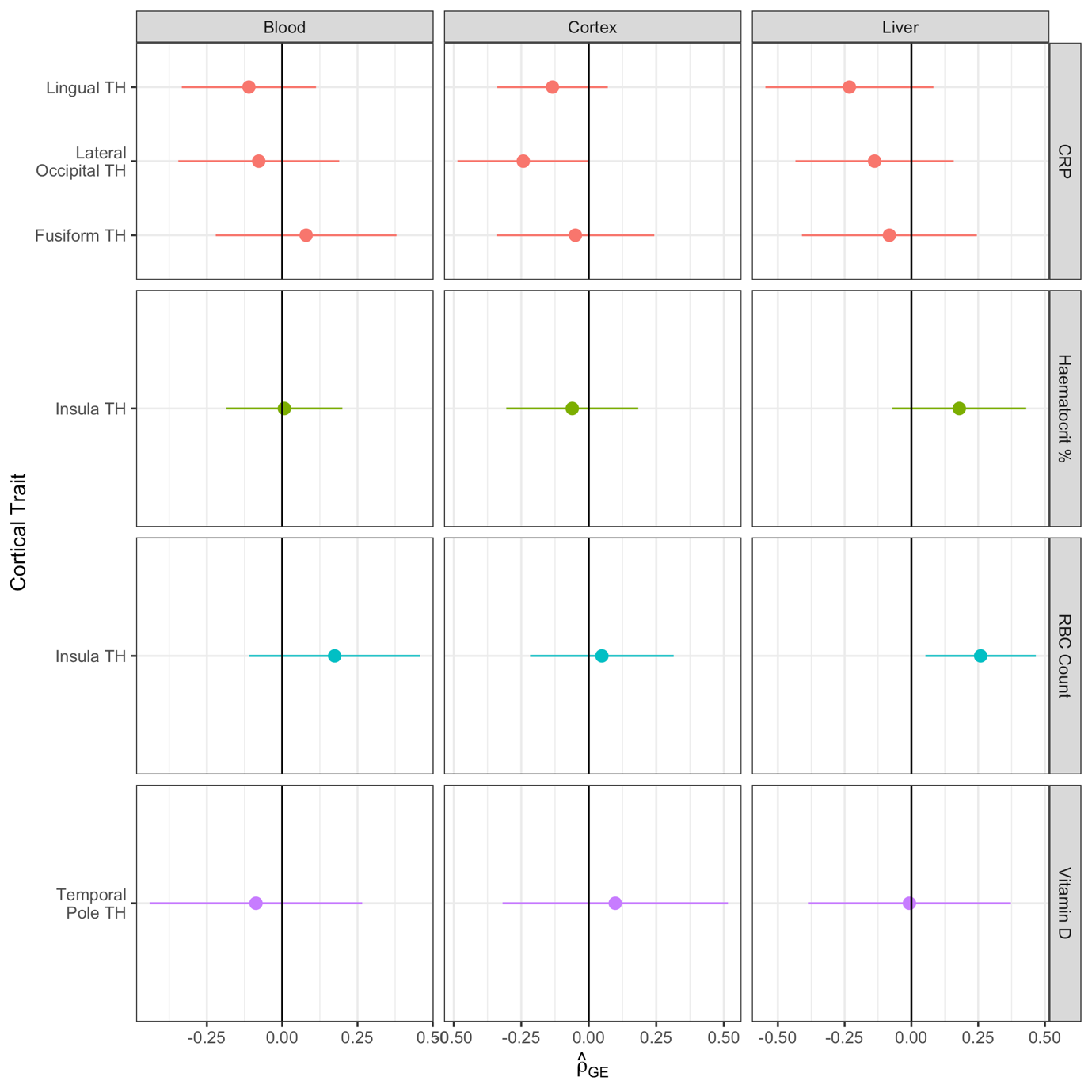

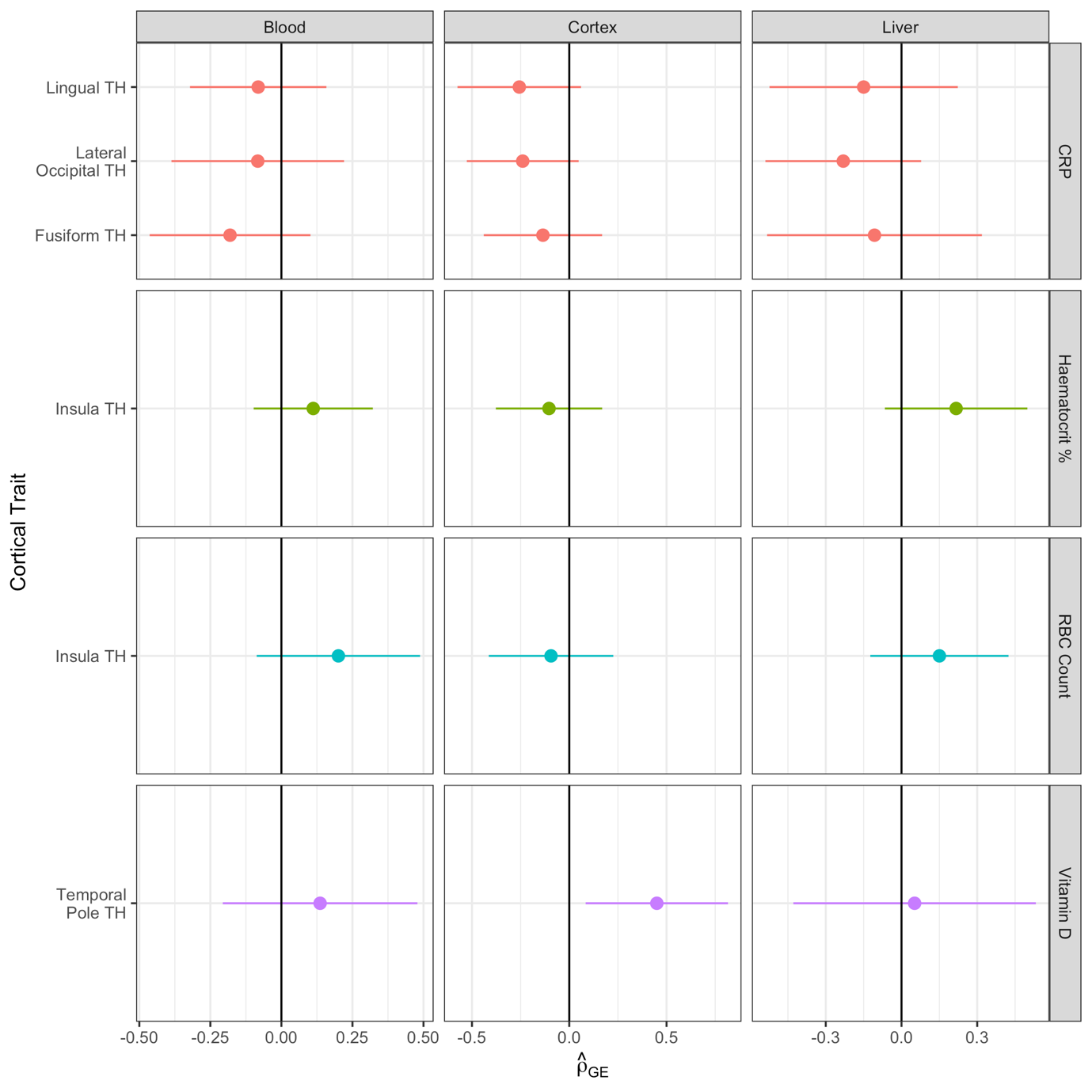

**b**

**Figure S4. Forest plots of all RHOGE results. (a)** Correlation of TWAS expression profiles amongst six trait pairings identified in Figure 3, using cortical GWAS without correction for global thickness. Note that no trait pairings were significantly correlated after correction for multiple testing (*P_Bonferroni_* < 0.05), however RBC count and insula thickness ($\hat{\rho}$*_GE_* = 0.26, *SE* = 0.11, *P* = 0.02) was nominally significant. **(b)** As in **(a)**, except utilising cortical GWAS with global correction applied. Data presented as $\hat{\rho}$*_GE_* (TWAS correlation) ± 95% CI. Similarly to **(a)**, no trait pairings survived correction for multiple testing, however the comparison of vitamin D and temporal pole thickness ($\hat{\rho}$*_GE_* = 0.45, *SE* = 0.19, *P* = 0.02) was nominally significant.
